# Effect mechanisms of different malaria chemoprevention regimens in pregnancy on infant growth outcomes: causal mediation analysis of a randomized controlled trial

**DOI:** 10.64898/2026.04.17.26351121

**Authors:** Anna T Nguyen, Joaniter I Nankabirwa, Abel Kakuru, Michelle E. Roh, Miriam Aguti, Harriet Adrama, Jimmy Kizza, Peter Olwoch, Moses R. Kamya, Grant Dorsey, Prasanna Jagannathan, Jade Benjamin-Chung

## Abstract

**Introduction:** Intermittent preventive treatment in pregnancy (IPTp) with sulfadoxine-pyrimethamine (SP) has become less effective at preventing malaria due to rising parasite resistance. IPTp with dihydroartemisinin-piperaquine (DP) alone or in combination with SP (DP+SP) dramatically lowers the risk of malaria in pregnancy compared to SP but is associated with lower birthweight and early life wasting. We estimated the effect of IPTp-DP, DP+SP, and SP on infant growth outcomes and assessed possible treatment mechanisms through a causal mediation analysis.

**Methods:** We used infant follow-up data (N=761) from a trial (NCT04336189) that randomized pregnant women to receive monthly IPTp-DP, SP, or DP+SP. We compared weight-for-length (WLZ) and length-for-age (LAZ) z-scores between treatment arms. We assessed possible mediation through pregnancy, birth, and infancy factors using interventional indirect effect models.

**Results:** Compared to IPTp-SP, IPTp-DP+SP decreased mean WLZ by 0.18 [95% confidence interval (CI) -0.03, 0.39] between 1-3 months and 0.28 (95% CI 0.07, 0.49) between 4-6 months, with the largest differences among primigravidae. Lower risk of active placental malaria in IPTp-DP+SP helped reduce differences in mean WLZ vs IPTp-SP (+0.06, 95% CI 0.02, 0.10). The IPTp-DP+SP arm had up to 0.28 lower mean LAZ between 7-13 months compared to IPTp-DP, particularly among children who were wasted between 0-6 months; low birthweight had a persistent, mediating effect on linear growth.

**Conclusion:** Adverse birth outcomes contributed to early growth faltering among children born to mothers receiving IPTp-DP+SP vs IPTp-SP, but the prevention of placental malaria partially counteracted the negative effects of IPTp-DP+SP on ponderal growth.

**Key Messages:**

1. Intermittent preventive treatment of malaria in pregnancy (IPTp) with a combination of dihydroartemisinin-piperaquine (DP) and sulfadoxine-pyrimethamine (SP) leads to lower birth weight compared to SP alone, but it is unclear whether effects persist through infancy and what mechanisms drive these differences.
2. DP+SP provided some improvements to ponderal growth over SP by preventing active placental malaria, but these benefits were not large enough to offset negative effects associated with other prenatal factors.
3. Infants born to mothers who received IPTp with DP+SP were at higher risk of growth faltering in the first year of life compared to DP or SP alone; while differences in weight-for-length subsided over time, some children developed chronic forms of malnutrition that may be difficult to recover from.

## Introduction

Annually, there are 121.9 million pregnancies in malaria endemic regions, with nearly 40% of these pregnancies occurring in Africa.^1^ Malaria during pregnancy carries substantial maternal and fetal risks, including severe maternal anemia, preterm birth, intrauterine growth restriction, low birthweight (LBW), and perinatal death.^2–5^ These conditions negatively impact on infant growth, increasing the risk of later health issues.^6^ Intermittent preventive treatment during pregnancy (IPTp) may improve infant growth by reducing malaria infections in pregnancy.^7–10^

In settings with moderate-to-high malaria transmission, the World Health Organization recommends IPTp with sulfadoxine-pyrimethamine (IPTp-SP), but widespread parasite resistance has made it less effective in Eastern and Southern Africa.^11,12^ IPTp with dihydroartemisinin-piperaquine (IPTp-DP) effectively prevents malaria in pregnancy, but is associated with lower birthweights and increased growth faltering in early infancy compared to IPTp-SP.^13^ SP’s broad-spectrum activity was thought to improve fetal growth relative to DP by preventing non-malarial infections.^14–17^

A recent trial in Uganda tested IPTp with a combination of DP and SP (IPTp-DP+SP) to assess whether their combined effect on reducing the risk of adverse birth outcomes would be greater than either drug alone (NCT04336189).^18^ IPTp-DP+SP did not improve malaria outcomes compared to IPTp-DP and, surprisingly, was associated with higher risks of small-for-gestational age (SGA) and LBW compared to IPTp-SP. Infants born to mothers in the IPTp-DP+SP arm faced higher wasting prevalence up to 6 months of age compared to IPTp-DP or IPTp-SP, but differences largely resolved by 12 months of age.^19^ While the trial concluded that IPTp-DP+SP was not a suitable replacement for IPTp-SP, it was unclear why infants in the IPTp-DP+SP arm had worse growth. Identifying specific pathways through which IPTp-DP, SP, and DP+SP impact infant growth could help explain why the combined therapy did not provide additional benefits.

In this study, we evaluated the causal pathways through which IPTp with DP, SP, or DP+SP affected infant growth in the first year of life using data from a randomized trial in Busia, Uganda.^18^ We considered the relative impacts of various maternal, birth, and infant factors, leveraging randomized interventional effect estimators to account for possible dependencies between mediators.

## Methods

We published a pre-analysis plan for this study at https://osf.io/rf4yp/ and listed deviations from this plan in Supplement 1.

### Study Population and Design

We analyzed follow-up data from a double-blinded randomized controlled trial (NCT04336189) conducted in Busia District, Uganda. The trial enrolled 2,757 HIV-negative pregnant women at 12-20 weeks’ gestation; additional details on recruitment and eligibility were previously reported.^18^ In brief, women were block randomized 1:1:1 to receive monthly DP, SP, or DP+SP starting in their second trimester. Routine visits were scheduled every four weeks, during which women received IPTp, underwent clinical examinations, and provided blood samples to test for malaria parasitemia by microscopy and polymerase chain reaction. Women were asked to visit the study clinic for all medical care. At delivery, birthweight and gestational age were recorded, and placental tissue was collected to assess placental malaria by histopathology.

Women who delivered a liveborn infant (N = 2,538) were invited to participate in a follow-up study to assess infant outcomes; enrollment was sequential and continued until at least 870 infants enrolled. Infants attended routine visits every four weeks through one year of age, and caretakers were asked to bring children to the study clinic for all medical care. Blood samples were taken by fingerprick every four weeks and any time children presented with an observed fever (≥38°C) or reported fever in the past 24 hours. Malaria parasitemia was assessed using blood smear by microscopy or polymerase chain reaction. Clinicians recorded a single height and weight measurement during each visit.

Positive malaria cases in pregnant women and infants were treated according to Ugandan Ministry of Health guidelines. Child malnutrition was treated according to WHO guidelines for pediatric hospital care, which includes health education and the use of therapeutic milk products (F-75, F-100) for severe acute malnutrition.

### Data

#### Growth Outcomes

We calculated weight-for-length z-scores (WLZ) and length-for-age z-scores (LAZ) from routine anthropometric measurements using the 2006 World Health Organization child growth standards.^20^ We excluded children with missing height or weight measurements or with z-scores below -6 or above +6 at any timepoint (N = 53) to minimize risks of measurement error when assessing age of incident stunting or wasting. To account for the potential influence of gestational age on estimated LAZs, we conducted a sensitivity analysis on treatment effects from 0-6 months using INTERGROWTH-21 standards based on children’s post-menstrual age (gestational age at birth plus postnatal age).^21^

Our primary outcomes were mean LAZ and WLZ at 1-3 months, 4-6 months, 7-9 months, and 10-12 months of age. Secondary outcomes included incidence of wasting (WLZ < - 2) and stunting (LAZ < -2) across the same age groups. Children were considered at risk of incident stunting if they had never been stunted, and at risk of incident wasting if they were not wasted in the prior 60 days.

#### Mediators

We considered mediators measured during pregnancy, delivery, and infancy. Pregnancy mediators included symptomatic malaria, parasitemia, placental malaria (any, active, past), mean weekly gestational weight gain, anemia, antibiotic use, and iron supplementation. Birth mediators included LBW, preterm birth, and SGA. Infant mediators included symptomatic malaria, parasitemia, anemia, and medication use, separately assessed at prior timepoints. Detailed mediator definitions are in Supplement 2.

#### Covariates

We adjusted all statistical models for potential confounders identified in our directed acyclic graph (Supplement 3), including child sex, gravidity, birth year, birth quarter, sickle cell trait, maternal age, maternal education, maternal height, maternal weight at enrollment, household wealth index, drinking water source, sanitation facility, and housing construction.

Additionally, we stratified analyses by infant sex and gravidity.

### Statistics

We modeled mean WLZs and LAZs by child age in each treatment arm using generalized additive models with cubic splines and child-specific random effects. We estimated 95% confidence intervals using a nonparametric bootstrap that resampled at the child level for 1,000 iterations. We approximated the empirical distribution of WLZs and LAZs using Gaussian kernel density estimation.

We estimated the total effect for DP+SP vs SP, DP+SP vs DP, and DP vs SP on growth outcomes using generalized linear models with a Gaussian family for continuous outcomes and log-binomial models with robust standard errors for binary outcomes.^22^ If log-binomial models failed to converge, we used modified Poisson regression models with robust standard errors.^23^ If a model had fewer than 20 total events or no events for a given level of the independent variable, we did not report an effect or proceed with mediation analyses.

We estimated treatment-mediator and mediator-outcome effects using the same modeling approach described above. For mediator-outcome effects, we only assessed mediators that preceded outcome measurement. We performed mediation analyses if both the treatment-mediator and mediator-outcome relationships had a p-value < 0.05 or large effect size (risk ratio < 0.9 or risk ratio > 1.1 for binary outcomes, absolute mean difference > 0.5 for continuous outcomes).

We estimated interventional indirect effects, as implemented by the “crumble” R package version 0.1.2.^24^ (Additional details in Supplement 4). We assessed the influence of individual candidate mediators, as well as the joint effect of all mediators that passed treatment-mediator and mediator-outcome screening. We adjusted for preceding mediators on the causal pathway that might introduce intermediate confounding (Supplement 3). For example, mediation analyses assessing child malaria as a potential mediator were adjusted for SGA, since IPTp affects SGA and SGA may influence child malaria risk. The interventional indirect effect would then represent the effect of IPTp on growth through the reduction of child malaria, independent from any effects through SGA.

## Results

There were 761 infants born between September 2021 and January 2024 with complete growth data in the first year of life included in the analyses. (Figure 1) The average age of enrollment was 5.2 weeks (SD = 0.9). Baseline characteristics and gravidity were similar across treatment groups, but there were differences in time of birth between the original trial and this follow-up study. (Table 1, Table S1). Additionally, children in the DP+SP arm who were included follow-up study had higher birth lengths than those in the original trial.

**Figure 1:**
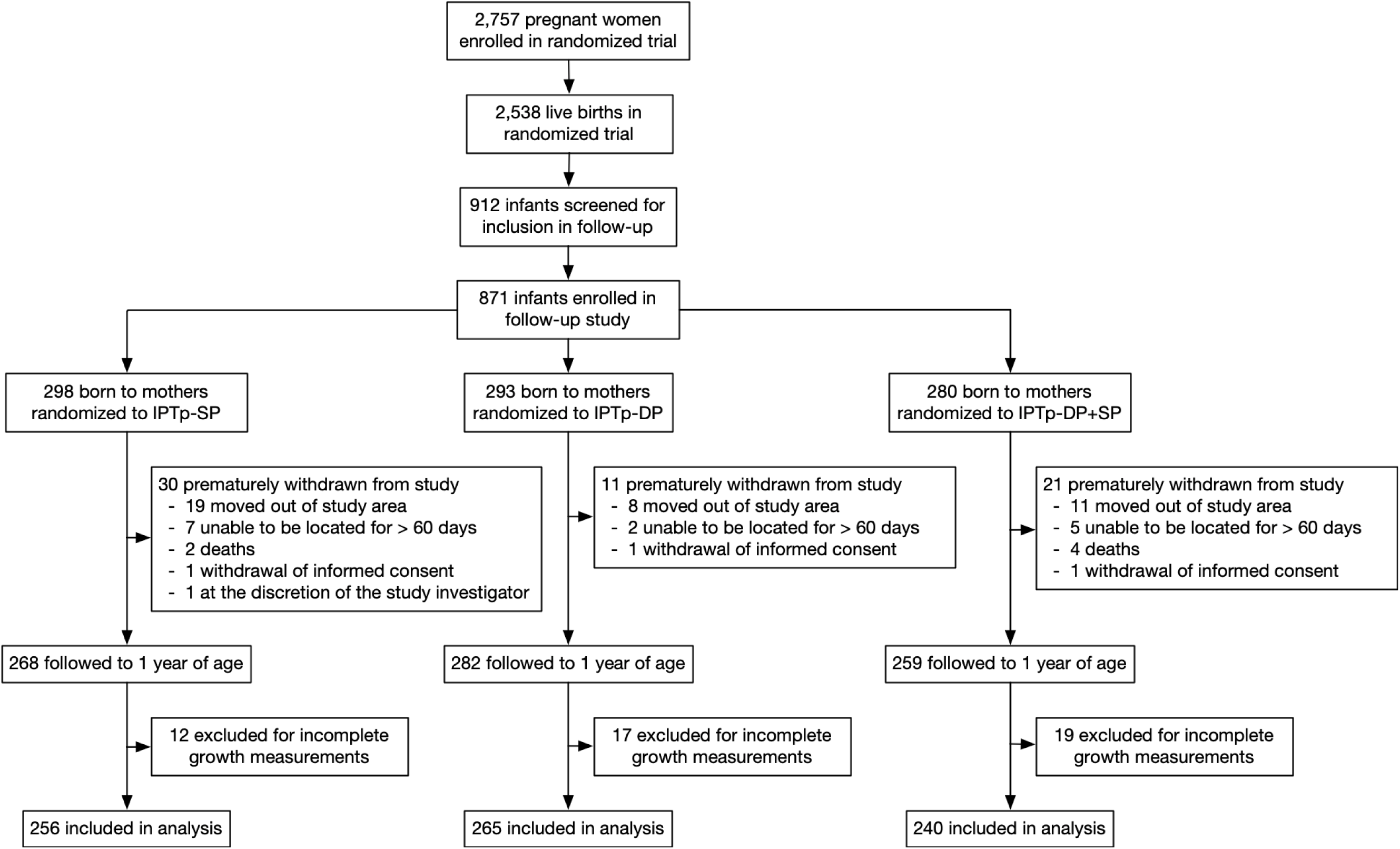
Consort Diagram of Study Cohort. Flowchart showing participants that were included in the current analysis. Children were included if their mothers were enrolled in a randomized trial of intermittent preventive treatment, selected for and enrolled in the infant follow-up study, and had complete monthly weight/height data through the first year of life.

**Table 1:** Baseline Characteristics of Study Participants.

| Variable |  | DP+SP | DP | SP |
| --- | --- | --- | --- | --- |
| *Infant Characteristics* |  |  |  |  |
| N |  | 240 | 265 | 256 |
| Age at Enrollment |  | 5.1 (0.7) | 5.1 (0.8) | 5.0 (0.7) |
| Child gender | Male | 115 (47.9%) | 143 (54.0%) | 138 (53.9%) |
|  | Female | 125 (52.1%) | 122 (46.0%) | 118 (46.1%) |
| Sickle cell trait | HbAA | 193 (80.4%) | 196 (74.0%) | 205 (80.1%) |
|  | HbAS | 47 (19.6%) | 66 (24.9%) | 50 (19.5%) |
|  | HbSS | 0 (0%) | 3 (1.1%) | 1 (0.4%) |
| Birth year | 2021 | 15 (6.2%) | 17 (6.4%) | 16 (6.2%) |
|  | 2022 | 48 (20.0%) | 72 (27.2%) | 69 (27.0%) |
|  | 2023 | 167 (69.6%) | 168 (63.4%) | 158 (61.7%) |
|  | 2024 | 10 (4.2%) | 8 (3.0%) | 13 (5.1%) |
| Birth season | Jan-Mar | 33 (13.8%) | 44 (16.6%) | 54 (21.1%) |
|  | Apr-Jun | 52 (21.7%) | 38 (14.3%) | 46 (18.0%) |
|  | Jul-Sep | 62 (25.8%) | 78 (29.4%) | 73 (28.5%) |
|  | Oct-Dec | 93 (38.8%) | 105 (39.6%) | 83 (32.4%) |
| Birth weight (kg) |  | 3.1 (0.5) | 3.1 (0.4) | 3.1 (0.5) |
| Birth length (cm) |  | 47.0 (3.2) | 46.9 (3.2) | 46.7 (3.1) |
| Birth length-for-age z-score |  | -1.3 (1.7) | -1.4 (1.7) | -1.5 (1.6) |
| Birth weight-for-length z-score |  | 0.6 (2.0) | 0.6 (1.9) | 0.8 (1.7) |
| Born preterm |  | 9 (3.8%) | 11 (4.2%) | 7 (2.7%) |
| Low birth weight |  | 15 (6.2%) | 17 (6.4%) | 9 (3.5%) |
| Small-for-gestational age |  | 58 (24.2%) | 56 (21.1%) | 52 (20.3%) |
| *Maternal Characteristics* |  |  |  |  |
| Gravidity | Primigravida | 52 (21.7%) | 60 (22.6%) | 57 (22.3%) |
|  | Multigravida | 188 (78.3%) | 205 (77.4%) | 199 (77.7%) |
| Maternal age (at birth) |  | 25.5 (6.0) | 26.4 (6.4) | 25.7 (6.3) |
| Maternal height at enrollment (cm) |  | 159.3 (6.3) | 158.1 (7.6) | 159.0 (5.9) |
| Maternal weight at enrollment (kg) |  | 56.8 (9.1) | 56.9 (8.4) | 56.6 (9.5) |
| Maternal gestational weight gain per week (kg/week) |  | 0.2 (0.1) | 0.2 (0.2) | 0.2 (0.1) |
| Maternal education level | None | 10 (4.2%) | 5 (1.9%) | 17 (6.6%) |
|  | Primary | 161 (67.1%) | 170 (64.2%) | 167 (65.2%) |
|  | Secondary | 68 (28.3%) | 84 (31.7%) | 69 (27.0%) |
|  | Higher | 1 (0.4%) | 6 (2.3%) | 3 (1.2%) |
| *Household Characteristics* |  |  |  |  |
| Household wealth | Poorest | 82 (34.2%) | 82 (30.9%) | 91 (35.5%) |
|  | Middle | 79 (32.9%) | 98 (37.0%) | 87 (34.0%) |
|  | Least Poor | 79 (32.9%) | 84 (31.7%) | 78 (30.5%) |
|  | Unknown | 0 (0%) | 1 (0.4%) | 0 (0%) |
| Household drinking water source | Unimproved | 9 (3.8%) | 11 (4.2%) | 24 (9.4%) |
|  | Improved | 231 (96.2%) | 253 (95.5%) | 232 (90.6%) |
|  | Unknown | 0 (0%) | 1 (0.4%) | 0 (0%) |
| Household sanitation facility | Uncovered Pit Latrine or No | 112 (46.7%) | 105 (39.6%) | 128 (50.0%) |
|  | Flush Toilet, Ventilated Improved Pit Latrine, or Covered Pit Latrine | 128 (53.3%) | 159 (60.0%) | 128 (50.0%) |
|  | Unknown | 0 (0%) | 1 (0.4%) | 0 (0%) |
| Housing construction | Traditional | 167 (69.6%) | 181 (68.3%) | 183 (71.5%) |
|  | Modern | 73 (30.4%) | 83 (31.3%) | 73 (28.5%) |
|  | Unknown | 0 (0%) | 1 (0.4%) | 0 (0%) |

Baseline characteristics of study participants. For categorical characteristics, the count and percent of relevant children are shown. For continuous characteristics, the mean value and standard deviation are shown.

### Total Effects of IPTp on Growth

Compared to IPTp-SP, infants in the IPTp-DP+SP and IPTp-DP arms had lower mean WLZs at birth.^19^ In the overall population, mean WLZ was 0.28 (95% CI 0.07, 0.49) lower in IPTp-DP+SP vs IPTp-SP between 4-6 months, but differences appeared to resolve by 7 months. (Figure 2, Table 2, Table S2). Children in the IPTp-DP+SP arm also had higher incidence of wasting (WLZ < 2 SD) at between 1-3 months compared to IPTp-SP (IRR = 1.55, 95% CI 0.94, 2.57) or IPTp-DP (IRR = 1.85, 95% CI 1.08, 3.16). (Figure S1, Table S3).

**Figure 2:**
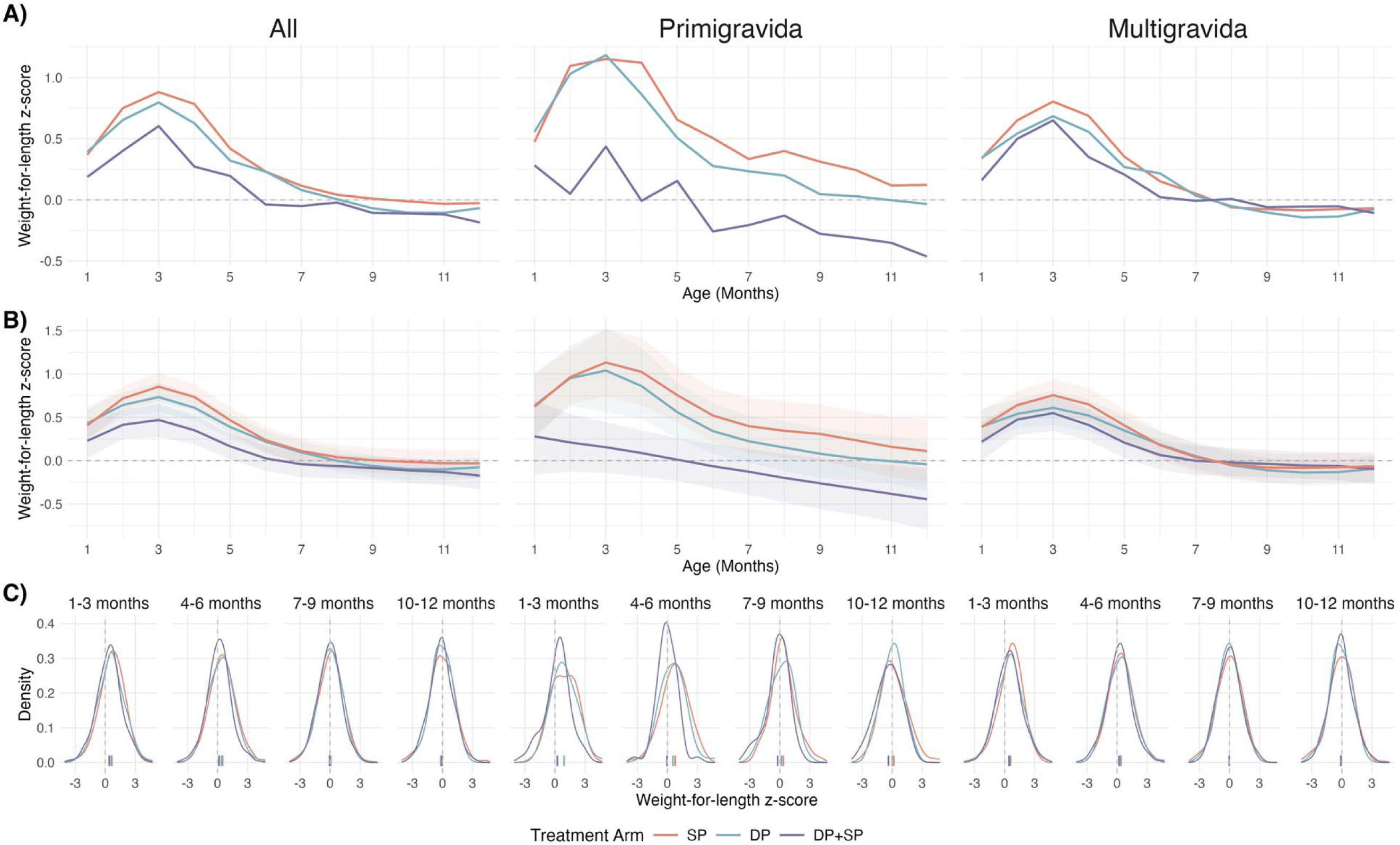
Differences in Weight-for-Length by Treatment Arm and Gravidity. A) Unadjusted mean weight-for-length z-scores (WLZ) by age in each IPTp-arm, stratified by gravidity. B) Mean WLZs, estimated using generalized additive models with cubic splines for age and child-specific random effects. 95% confidence intervals were estimated using a nonparametric bootstrap that resampled by child for 1,000 iterations. Models were adjusted for child sex, gravidity, birth year, birth quarter, maternal age, maternal education, maternal height, maternal weight at enrollment, household wealth index, drinking water source, and sanitation facility. C) Smoothed distributions of observed WLZs in each treatment arm, generated through Gaussian kernel density estimation. Empirical means for each distribution are displayed as ticks on the x-axis.

**Table 2:**
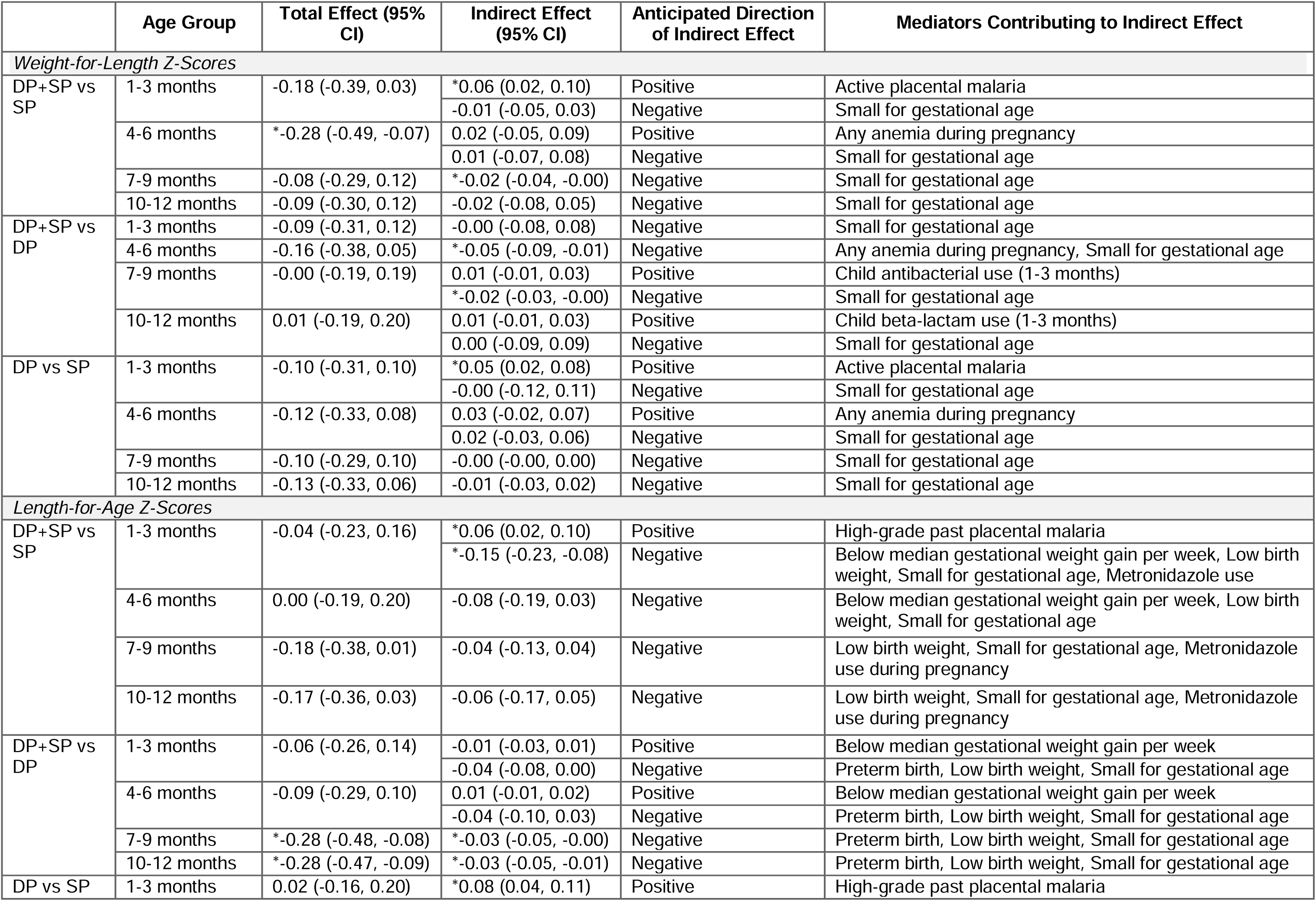
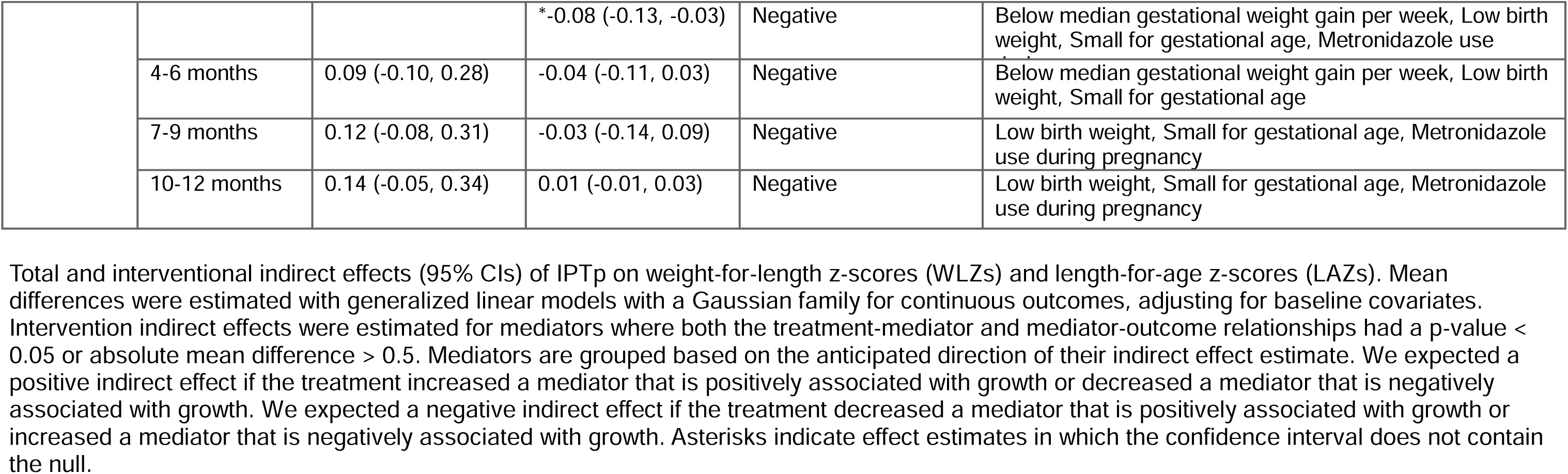
Total and Interventional Indirect Effects of intermittent Preventive Treatment in Pregnancy on Mean Weight-for-Length and Length-for-Age Z-Scores.

However, gravidity subgroup analyses showed that the negative effect of IPTp-DP+SP vs IPTp-SP persisted across all ages in children born to primigravid women, with the largest difference in WLZ between 4-6 months (-0.99, 95% CI -1.47, -0.51). This negative effect was not observed among multigravidae, for whom the largest difference in WLZ was at birth (-0.23, 95% CI -0.65, 0.20). Mean WLZs in IPTp-DP and IPTp-SP were similar from 1-13 months, but modestly favored IPTp-SP early in life. Trends in mean WLZs by age were similar by child sex. (Figure S2)

Mean LAZs were similar across arms between 1-6 months, but in later infancy, LAZs decreased in IPTp-DP+SP relative to IPTp-DP or IPTp-SP. (Figure 3, Table S4) At 7-9 months, mean LAZs in IPTp-DP+SP were 0.18 (95% CI -0.01, 0.38) lower than IPTp-SP and 0.28 (95% CI 0.08, 0.48) lower than IPTp-DP. At 10-12 months, mean LAZs in IPTp-DP+SP were 0.17 (95% CI -0.03, 0.36) lower than IPTp-SP and 0.28 (95% CI 0.09, 0.47) lower than IPTp-DP. We saw the largest differences in LAZs among children who were wasted in the first 6 months of life, male, or born to multigravida. (Figure S3-S4). Incidence of first-time stunting was higher in IPTp-DP vs IPTp-SP between 1-3 months (IRR 1.57, 95% CI 1.07, 2.30), but we found no other differences in stunting incidence by treatment arm at other ages. (Figure S5, Table S5).

**Figure 3:**
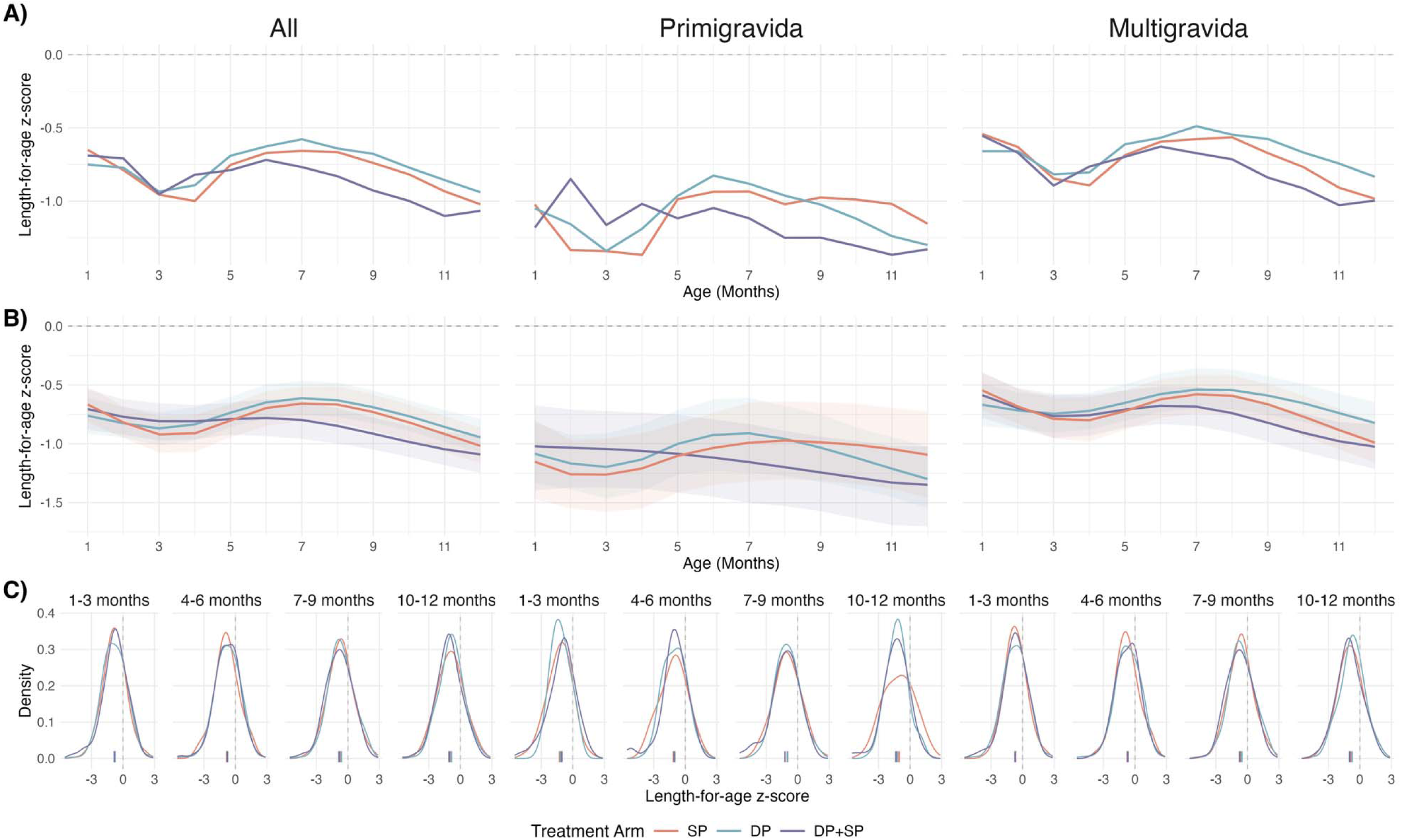
Differences in Length-For-Age Z-Scores by Treatment Arm and Gravidity. A) Unadjusted mean length-for-age z-scores (LAZ) by age in each IPTp-arm, stratified by gravidity. B) Mean LAZs estimated using generalized additive models with cubic splines for age and child-specific random effects. 95% confidence intervals were estimated using a nonparametric bootstrap that resampled by child for 1,000 iterations. Models were adjusted for child sex, gravidity, birth year, birth quarter, maternal age, maternal education, maternal height, maternal weight at enrollment, household wealth index, drinking water source, and sanitation facility. C) Smoothed distributions of observed LAZs, generated through Gaussian kernel density estimation. Empirical means for each distribution are displayed as ticks on the x-axis.

### Effect of IPTp on Candidate Mediators

Compared to IPTp-SP, IPTp-DP and IPTp-DP+SP reduced uncomplicated malaria in pregnancy, parasitemia in pregnancy, and placental malaria, but increased LBW and decreased gestational weight gain. (Figure 4A, Table S6). We also found that IPTp-DP+SP and IPTp-DP increased SGA compared to IPTp-SP, but only in multigravida women.

**Figure 4:**
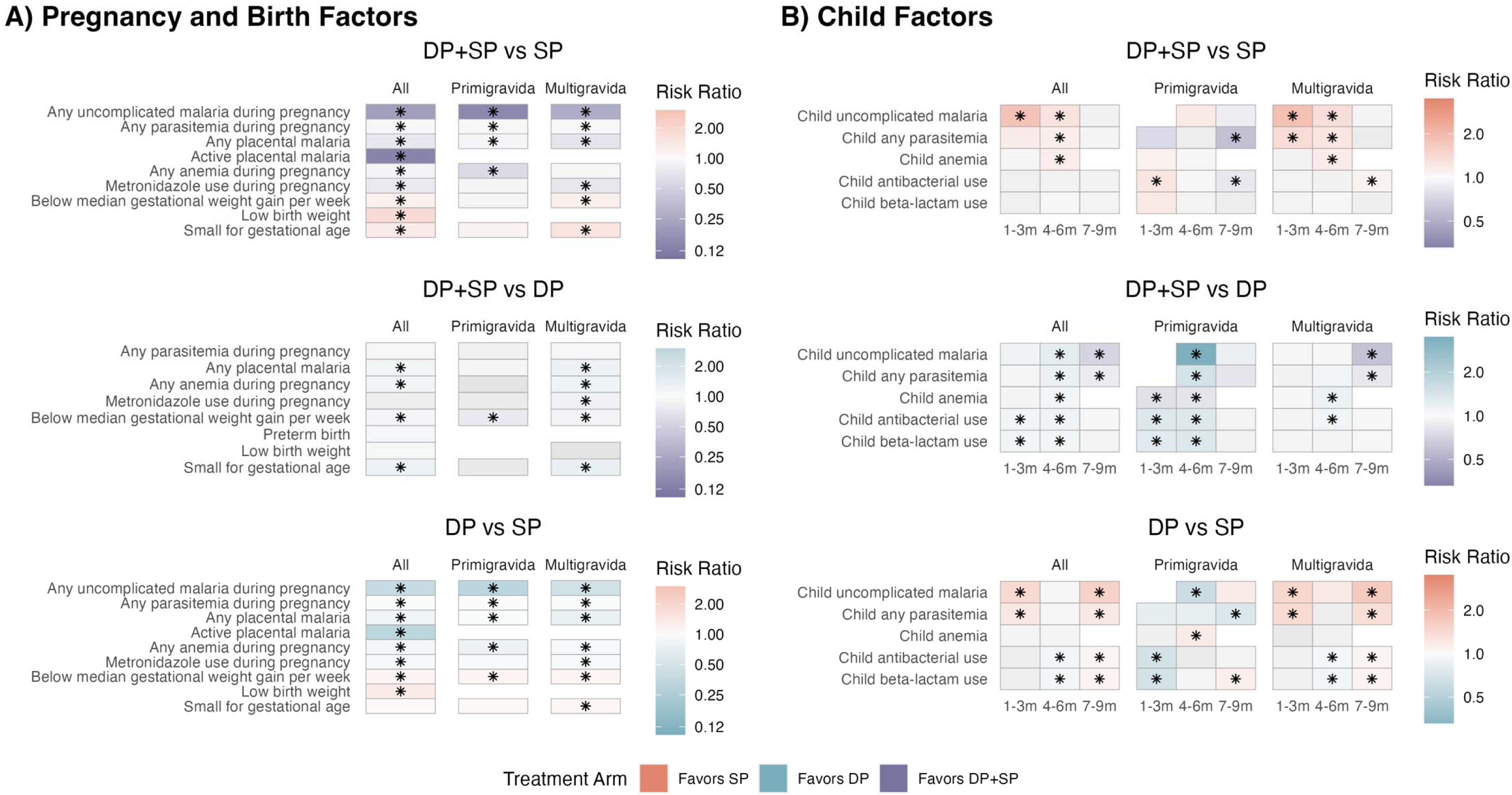
Effects of IPTp on Candidate Mediators Measured During Pregnancy, Birth, and Infancy. Heatmap of estimated prevalence ratios for IPTp regimens on (A) pregnancy and birth factors and (b) infant factors measured at different ages. Prevalence ratios were estimated with log-binomial models with robust standard errors, adjusting for baseline covariates. If log-binomial models failed to converge, we used modified Poisson regression models. Tiles are shaded in color if the prevalence ratio is large (PR > 1.1, PR < 0.9) or had a p-value < 0.05. Color is determined by the comparison arm with the lower risk of the mediator (the arm “favored” to provide protection against the mediator). Asterisks mark prevalence ratios that were statistically significant. Effects were not estimated if there were less than 20 observed cases, and are indicated by the absence of a tile.

IPTp-DP+SP and IPTp-DP increased infant malaria and parasitemia compared to IPTp-SP between 1-3 months, though prevalence was low in all arms. (Figure 4B, Figure S6). IPTp-DP+SP increased child antibacterial use compared to IPTp-DP between 1-6 months particularly among children born to primigravida women.

### Effect of Candidate Mediators on Outcomes

Active placental malaria at delivery was associated with lower mean WLZ at 1-3 months (-0.53, 95% CI -0.96, -0.09). SGA was associated with lower mean WLZ (range -0.21 to -0.33) in all age groups, but associations were largely concentrated among multigravida women (Figure S7, Table S7). We found similar mediator-outcome relationships by child sex. (Figure S8). Low LBW was associated with elevated wasting incidence between 1-3 months (PR = 2.83, 95% CI 1.64, 4.89). (Figure S9, Table S8)

Preterm birth, LBW, and SGA were associated with lower LAZ at all ages, and with increased stunting incidence at 1-3 months. (Table S9-10, Figure S10). We found stronger effects of SGA on LAZs in multigravida women, but no differences by child sex. (Figure S7, Figure S8)

### Mediated Effects of IPTp on Growth

In early infancy, mean WLZs were lower in IPTp-DP+SP and IPTp-DP compared to IPTp-SP. Compared to IPTp-SP, the IPTp-DP+SP and IPTp-DP arms increased WLZ between 1-3 months by 0.06 (95% CI 0.02, 0.10) and 0.05 (95% CI 0.02, 0.08), respectively, by preventing active placental malaria. (Table 2, Figure S11, Table S11) The indirect effect through active placental malaria was particularly strong in children who were male or born to primigravida women. (Figure S12-S13). The prevention of placental malaria also contributed to reduced wasting incidence, though confidence intervals contained the null (Table S12).

Increased SGA in IPTp-DP+SP contributed to decreased mean WLZs compared to IPTp-DP or IPTp-SP, but these indirect effects were relatively small.

Mean LAZs were similar across arms in early infancy, but after 7 months, mean LAZ decreased in IPTp-DP+SP compared to IPTp-DP or IPTp-SP. Mediators that measured or influenced fetal growth (gestational weight gain per week, LBW, SGA, preterm birth) and metronidazole use during pregnancy in IPTp-DP+SP played the largest mediating role, contributing to a 0.15 (95% CI 0.08, 0.23) decrease in LAZ compared to IPTp-SP and a 0.08 (95% CI 0.03, 0.13) decrease in LAZ compared to IPTp-SP between 1-3 months. (Figure S15, Table S13). These effects were persistent but waned with age, contributing to reductions in mean LAZs between 10-12 months of -0.06 (95% CI -0.17, 0.05) in IPTp-DP+SP vs IPTp-SP and -0.03 (95% CI -0.05, -0.01) in IPTp-DP+SP vs IPTp-DP. The indirect, negative effects of SGA in IPTp-DP+SP vs IPTp-SP were larger in female children and children born to multigravida women. (Figure S16-S17). SGA also led to modest increases in wasting incidence between 1-6 months in IPTp-DP+SP vs IPTp-SP. (Table S14). Reductions in high-grade placental malaria contributed to increased LAZs of 0.06 (95% CI 0.02, 0.10) in IPTp-DP+SP and 0.08 (0.04, 0.11) in IPTp-DP compared to SP, offsetting the negative indirect effects of fetal growth restriction in early infancy.

### Sensitivity Analysis

Differences in mean LAZs and stunting incidence based on postmenstrual age were consistent with estimates under WHO standards. The association of preterm birth, SGA, and LBW on LAZs and stunting incidence were weaker under INTERGROWTH-21^st^ standards. The indirect effects of these fetal growth factors on linear growth were attenuated but confidence intervals still do not contain the null. (Supplement 5).

## Discussion

Children born to mothers receiving IPTp-DP+SP had worse ponderal growth in early life compared to IPTp-SP or IPTp-DP, leading to impaired linear growth which may have been mediated by the higher risk of SGA and LBW associated with IPTp-DP+SP. The estimated interventional indirect effects on mean WLZ were substantially smaller than total treatment effects; this suggests that the negative effects of DP+SP vs SP on ponderal growth are driven by other, unmeasured pathways that outweigh benefits associated with malaria prevention during pregnancy. To our knowledge, this is the first study to investigate the intermediate role of infant factors on IPTp’s effect on infant growth outcomes, allowing us to quantify the relative impact of prenatal and postnatal factors.

IPTp-DP+SP resulted in up to 0.27 lower mean WLZs compared to IPTp-DP or IPTp-SP, with the largest differences in children born to primigravida. Children that were wasted in the first 6 months of life had lower mean LAZs between 7-12 months, suggesting that poor ponderal growth in early infancy developed into more chronic linear growth faltering. The prevention of placental malaria by IPTp regimens with DP increased mean WLZs and LAZs over IPTp-SP in early infancy, but these indirect effects were not large enough to offset other, persistent negative effects. This is consistent with studies that showed that placental malaria is linked to lower birthweight, and that IPTp might improve infant growth through the prevention of maternal malaria.^25–27^ Mothers receiving a regimen with DP were less likely to have active placental malaria at delivery, which contributed to a higher mean WLZs between 1-3 months despite overall negative effects of DP on ponderal growth. Infants born to primigravida women that lack immunity to placenta-adhesive parasites benefited the most from this anti-malaria effect. On the other hand, we found that the negative effects of fetal growth restriction on mean LAZs by IPTp regimens containing DP were larger among children born to multigravida women, who may experience weaker anti-malarial benefits. This is consistent with prior working showing that the effect of preventing placental malaria is weaker in subsequent pregnancies after immunity is established.^28–31^

The mediators we considered accounted for a very small portion of overall treatment effects for both WLZ and LAZ. Prior mediation analyses have highlighted the potential non-malarial benefits of SP over DP,^14,15^ and it is possible that we did not capture low-grade, chronic exposure to non-malarial pathogens that impact the relationship between IPTp and growth.

Alternatively, drug-drug interactions between DP and SP may have limited their combined effectiveness. A pharmacokinetic study nested in this trial found that DP+SP resulted in lower exposure to sulfadoxine and pyrimethamine compared to SP and lower exposure to piperaquine compared to DP.^33^ Here, we found that IPTp-DP+SP vs IPTp-SP had a smaller positive indirect effect on mean WLZ through the prevention of placental malaria compared to IPTp-DP vs IPTp-SP, reflecting the weaker anti-malarial effects of IPTp-DP+SP vs IPTp-DP. The relatively small indirect effects identified in this study appear to suggest there may be a negative effect of IPTp-DP on growth through an alternative, unidentified mechanism. Additional studies are needed to investigate the potential harm of DP on fetal and infant growth.

This study has several limitations. Not all children born in the randomized trial were included in the follow-up study, which may have limited statistical power and introduced selection bias due to differences in cohorts. Though we adjusted factors that may influence inclusion or other relationships between treatment, mediators, and outcomes, residual bias is possible. We redefined continuous mediators as binary or categorical variables, which may underestimate indirect effects for factors like gestational weight gain. We only assessed the first incidence of stunting, but there may be different effects on persistent or recurrent events, However, our study was not powered to assess these rare outcomes. Additionally, we had limited power to assess severe outcomes and estimate stratified mediated effects by gravidity and child sex. Finally, we did not correct for multiple comparisons and future trials are needed to validate these exploratory results.

## Conclusion

IPTp regimens that included DP resulted in fetal growth restriction, which had a persistent negative effect on ponderal and linear growth compared to IPTp-SP. Though IPTp regimens with DP reduced placental malarial, the benefits of preventing prenatal malaria were not large enough to offset DP’s negative effects on growth.

## Ethics approval

This study received ethical approval from Makerere University School of Biomedical Sciences Research and Ethics Committee (SBS-2021-052), the Uganda National Council of Science and Technology (HS 1819ES) and Stanford University (62131). Caregivers of participants provided written informed consent before enrolling the study.

## Acknowledgements

We appreciate the contributions of all study participants, as well as the staff who delivered the interventions and led data collection.

## Author Contributions

ATN, JBC, and PJ conceptualized the analysis. ATN and JBC designed the analytic strategy, which all authors reviewed and revised prior to the start of data analysis. JIN, AK, MA, HA, JK, and PO implemented the trial and led data curation. JIN, PJ, GD, PJR, and MRK acquired funding for the study. ATN performed statistical analyses and wrote a first draft of the manuscript, while JBC supervised. All authors reviewed, edited, and approved the final version of the manuscript.

## Data Availability

Data used in this analysis is publicly available in Stanford Research Data repository (https://doi.org/10.25740/rp609nq8772). All analyses were conducted using R version 4.4.1. Replication materials are publicly available at https://github.com/anna-nguyen/impact-growth-mediation-public.

## Use of Artificial Intelligence (AI) tools

AI tools were used to parallelize code and document functions for open-source materials, based on original analysis pipelines that were written by the authors. All AI output was manually checked and verified by ATN. No AI tools were used in the writing of the manuscript.

