## Supplemental Materials for "Effect mechanisms of different malaria chemoprevention regimens in pregnancy on infant growth outcomes: causal mediation analysis of a randomized controlled trial"

### Supplement 1: Deviations from Pre-Analysis Plan

1. Our pre-specified set of outcomes included severe stunting, severe wasting, and growth velocity. We could not assess severe stunting or wasting due to low incidence in this population. We did not include measures of growth velocity in this manuscript due to limited space.
2. During analysis, we found that some women had extreme fluctuations in their recorded weight during pregnancy. We believe these are attributed to data entry errors and used the following process to detect and correct extreme values:
   a. We flagged anomalies where there was an increase of >2kg/week followed by a decrease of >1 kg/week (or vice versa)
      i. These thresholds were based on the reccomended weight gain of 0.5 kg/week. We assume that a weight increase 4x higher than reccomended or decrease of 2x lower than reccomended was extreme.
   b. We imputed extreme values with a linear interpolation of the measurements taken immediately before and after.
3. We did not pre-specify small for gestational age as a mediator but included it to be comparable with other studies that report the impact of IPTp on birth outcomes.
4. We pre-specified confounder selection using LASSO regression. However, we found this process to be computationally prohibitive and difficult to interpret if each model adjusted for different confounder sets. We chose to adjust all models for the full confounders set, but also conducted a sensitivity analysis with a reduced confounders set to ensure that models were not being overfit. We also screened models to ensure that there were enough variables per event, and only reported effect estimates when there were 20 events observed for the dependent variable being modeled. Additionally, we did not report estimates when there were 0 events observed for any given level of the independent variable.
5. We stated that generalized additive models (GAMs) would be used to compare continuous outcomes. In the presented analysis, we used GAMs to model mean WLZs/LAZs by age in different treatment arms but used generalized linear models with a Gaussian family to estimate mean differences between arms.
6. We stated the 95% CIs for GAMS would be estimated using a parametric bootstrap. However, we implemented a nonparametric bootstrap that allowed us to marginalize across adjusted covariates.
7. We intended to include linear discriminant analysis (lda) and quadratic discriminant analysis (qda) in the SuperLearner used for the estimation of intervention indirect effects. However, we ran into issues with model convergence that led us to exclude them.
8. We estimated differences in LAZs between 7-13 months stratified by wasting status between 1-6 months, to support a post-hoc hypothesis that early differences in ponderal growth contributed to subsequent differences in linear growth.
9. We estimated the joint interventional indirect effects of groups of mediators that passed treatment-mediator or mediator-outcome screening. This allowed us to assess the indirect effect that was captured by any of the individual mediators.

### Supplement 2: Candidate Mediator Variable Definitions

| **Mediator** | **Type of measurement** | **Time of measurement(s), relative to growth at t=T** | **Measurement Method** | **Variable Definition** |
| --- | --- | --- | --- | --- |
| Placental malaria | Prevalence | Birth | Placental histopathology | 1. Active placental malaria (detected parasite via histopathology) [Yes/No]  2. Past placental malaria (detected pigment) [Yes/No]  3. High grade past infection (pigment in >30% of high powered field) [Yes/No]  4. Any placental malaria [Yes/No] |
| Peripheral malaria during pregnancy | Incidence | Duration of Pregnancy,  Second Trimester (13-28 weeks’ gestation), Third Trimester (29-40 weeks’ gestation) | Parasites detected in blood draws vs qPCR and/or microscopy  Symptomatic malaria = positive thick blood smear (given for fever in the past 24 hours or tympanic temperature ≥38·0°C) | 1. Parasitemia during pregnancy [Yes/No]  2. Symptomatic malaria (fever + microscopic parasitemia) during pregnancy [Yes/No]  3. Complicated malaria^†^ during pregnancy [Yes/No] |
| Antibiotic use during pregnancy | Prevalence | Routine Visits Throughout Pregnancy | Prescriptions | Any use of azithromycin, tetracycline, doxycycline, or metronidazole during pregnancy.^^^ [Yes/No] These antibiotics were selected due to their use in the treatment of STIs/RTIs, which may fall on the causal pathway between IPTp and child growth. |
| Maternal anemia | Prevalence | 20, 28, 36 weeks’ gestation | From blood draws | 1. Any anemia (Hb < 11 g/dL) [Yes/No]  2. Any severe anemia (Hb < 8 g/dL) [Yes/No] |
| Maternal Iron supplementation | Mean | Routine Visits Throughout Pregnancy | Prescriptions | Number of ferrous sulfate tablets taken by the mother during trial |
| Gestational weight gain | Mean | Birth | N/A | Gestational weight gain per week, estimated as the average weekly weight gain between 20 gestation weeks and the last antenatal care visit |
| Preterm birth | Prevalence | Delivery | Gestational age, based on ultrasounds | Delivery before 37 weeks’ gestation [Yes/No] |
| Low birthweight | Prevalence | Delivery | N/A | Birth weight ≤2500 g [Yes/No] |
| Small for Gestational Age | Prevalence | Delivery | N/A | Birthweight < 10th percentile for gestational age using INTERGROWTH-21st standards [Yes/No] |
| Child malaria | Incidence | t = 1-3 months, …, T - 1 | Parasites detected in blood draws vs qPCR and/or microscopy  Symptomatic malaria = positive thick blood smear (given for fever in the past 24 hours or tympanic temperature ≥38·0°C) | 1. Parasitemia during interval [Yes/No]  2. Symptomatic, uncomplicated malaria during interval [Yes/No]  3. Symptomatic, complicated malaria^†^ during interval [Yes/No] |
| Child anemia | Prevalence | 8, 24, and 52 weeks | From blood draws | 1. Any anemia (Hb < 11 g/dL) [Yes/No]  2. Any severe anemia (Hb < 8 g/dL) [Yes/No] |
| Child medication use | Prevalence | t = 1-3 months, …, T - 1 | Prescriptions of antimicrobials by class, as identified under the Anatomical Therapeutic Chemical classification system. | Any use of antimalarials, antiparasitics, antivirals, beta-lactams, fluoroquinolones, sulfonamides, or antibacterials^^^ in the interval [Yes/No] |

^†^ Complicated malaria cases are characterized by Parasitemia and one of the following criteria:

- Cerebral malaria (unarousable coma attributed to malaria)
- ≥ 3 convulsions over 24 hours
- Severe normocytic anemia (Hb < 5 gm/DL)
- Hypoglycemia
- Metabolic acidosis with respiratory distress
- Fluid and electrolyte disturbances
- Acute renal failure
- Acute pulmonary edema and adult respiratory distress syndrome
- Circulatory collapse, shock, septicemia
- Abnormal bleeding
- Jaundice
- Danger signs in children
  - Any convulsions in children
  - Inability to sit up or stand
  - Vomiting
  - Unable to breastfeed or drink
  - Lethargy

^^^Medication categories, assessed using prescription data:

- Azithromycin: Azithromycin
- Metronidazole: Metronidazole cream, Metronidazole injection, Metronidazole syrup, Metronidazole tablets
- Tetracycline: Tetracycline caps, Tetracycline eye ointment
- Doxycycline: Doxycycline
- Ferrous: Ferrous sulfate tablets, Ferrous sulfate/Folic acid
- Betalactams: Amoxicillin caps, Amoxicillin syrup, Ampicillin, Ampiclox caps, Ampiclox syrup, Augmentin syrup, Augmentin tablets, Benzathine penicillin injection, Benzyl Penicillin injecion, Cefixime, Ceftriaxone injection, Cefuroxime, Cephalexin syrup, Cephalexin tablets, Cloxacillin caps, Cloxacillin injection, Cloxacillin syrup
- Fluoroquinolones: Ciprofloxacin ear drops, Ciprofloxacin tablets, Ciprofloxacin, I.V.
- Sulfonamides: Co-trimoxazole, Co-trimoxazole injection, Co-trimoxazole syrup, Fansidar tablets (SP), Septrin, Sulfadoxine-pyrimethamine (SP), Trimethoprim-Sulfamethoxazole (TS)
- Other Antibacterial: Chloramphenicol caps, Chloramphenicol eye/ear drops, Chloramphenicol injection, Chloramphenicol syrup, Clindamycin caps, Erythromycin syrup, Erythromycin tablets, Gentamicin eye/ear drops, Gentamicin injection, Streptomycin injection
- Antimalarial: Artemether-lumefantrine, Artenum/Artemether, Artesunate, Chloroquine injection, Chloroquine tablets, Coartem (Artemether-lumefantrine), Fansidar tablets (SP), Quinine injection, Quinine tablets, Sulfadoxine-pyrimethamine (SP), Unknown antimalarial
- Antiparasitic: Mebendazole tablets
- Antiviral: Acyclovir cream, Acyclovir tablets

### Supplement 3: Directed Acyclic Graph and Covariate Variable Definitions

There are potential confounders that could bias treatment-outcome, treatment-mediator and mediator-outcome relationships. To minimize the risk of bias, we will adjust for the confounders (identified in a causal model) in all statistical models. Our confounder set includes factors that may inform selection into the study, and we assume that controlling for these variables is sufficient to block collider bias by selecting on survival and enrollment.

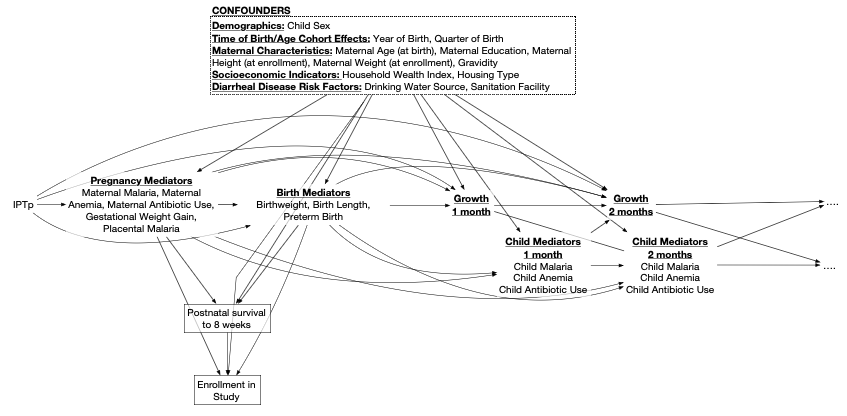

| **Risk Factor** | **Values** |
| --- | --- |
| Child Sex | Male, Female |
| Gravidity | Primigravida, Multigravida |
| Year of Birth | 2021, 2022, 2023, 2024 |
| Quarter of Birth | Jan-Mar, Apr-Jun, Jul-Sep, Oct-Dec |
| Maternal Age | Continuous (years) |
| Maternal Education | Primary or none, Secondary or higher |
| Maternal Height | Continuous |
| Maternal Weight at Enrollment | Continuous |
| Household Wealth Index | Poorest, Missing, Least Poor |
| Drinking Water Source | Unimproved, Improved |
| Sanitation Facility | Unimproved (uncovered pit latrine or no facility),  Improved (flush toilet, ventilated improved pit or covered pit latrine) |
| Housing Material | Traditional, Modern |

### Supplement 4: Estimation of Interventional Indirect Effects

We estimated interventional indirect effects, which compares counterfactual outcomes estimated under different mediator distributions. Unlike natural (in)direct effects, interventional indirect effects do not assume the absence of treatment-induced mediator-outcome confounders, which is a concern when working with multiple, sequential mediators. Instead of explicitly specifying a treatment-mediator model, we construct counterfactuals by simulating random draws of mediator values from distributions conditioned on treatment group. The interventional indirect effect then represents the effect of the treatment on the outcome through the mediator, independent of any upstream mediators. In other words, for A -> L -> M -> Y, the interventional indirect effect through M does not capture the downstream effects of L on M. This framework can be extended to assess the indirect effect through multiple mediators by simulating draws from their joint distributions.

We used the “crumble” R package (v0.1.2), which implements semi-parametric efficient estimators based on efficient influence functions (EIF). We used SuperLearner, an ensemble machine learning approach, to fit regression models of the EIF. We estimated the weights of the EIF using Riesz-learning, implemented with a multilayer perceptron. This method allowed us to construct weights for estimators without specifying the full functional form of the EIF. We crossfit both the SuperLearner models and perceptron to optimize model parameters.

The component models of SuperLearner are listed below, and were all implemented in the “mlr3superlearner” R package (v0.1.2) that is integrated into “crumble”.

For continuous outcomes, we included the following models:

- Mean
- Generalized linear models
- Generalized additive models
- Elastic nets (glmnet)
- Random forests (ranger)
- Neural nets (nnet)
- Extreme gradient boosting (xgboost)
- Support vector machines (svm)
- Bayesian Additive Regression Trees (bart)

For categorical outcomes, we included the following models:

- Mean
- Generalized linear models
- Elastic nets (glmnet)
- Random forests (ranger)
- Neural nets (nnet)
- Extreme gradient boosting (xgboost)
- Support vector machines (svm)
- Bayesian Additive Regression Trees (bart)

### Supplement 5: Sensitivity Analysis, LAZs under INTERGROWTH-21^st^ Standards

WHO growth standards use children’s chronological age to estimate length-for-age z-scores (LAZ). Children born earlier are often born smaller, but their postnatal growth is compared to the same standard as full-term children. Consequently, children with smaller gestational ages have systematically lower LAZs.

To assess the influence that gestational age might have on linear growth measurements, we conducted a sensitivity analysis estimating LAZs and stunting incidence under INTERGROWTH-21^st^ postnatal growth standards. These standards use post-menstrual age, which is a sum of gestational age and chronological age.

*Mean LAZ By Age and IPTp Arm, Stratified by Gravidity*

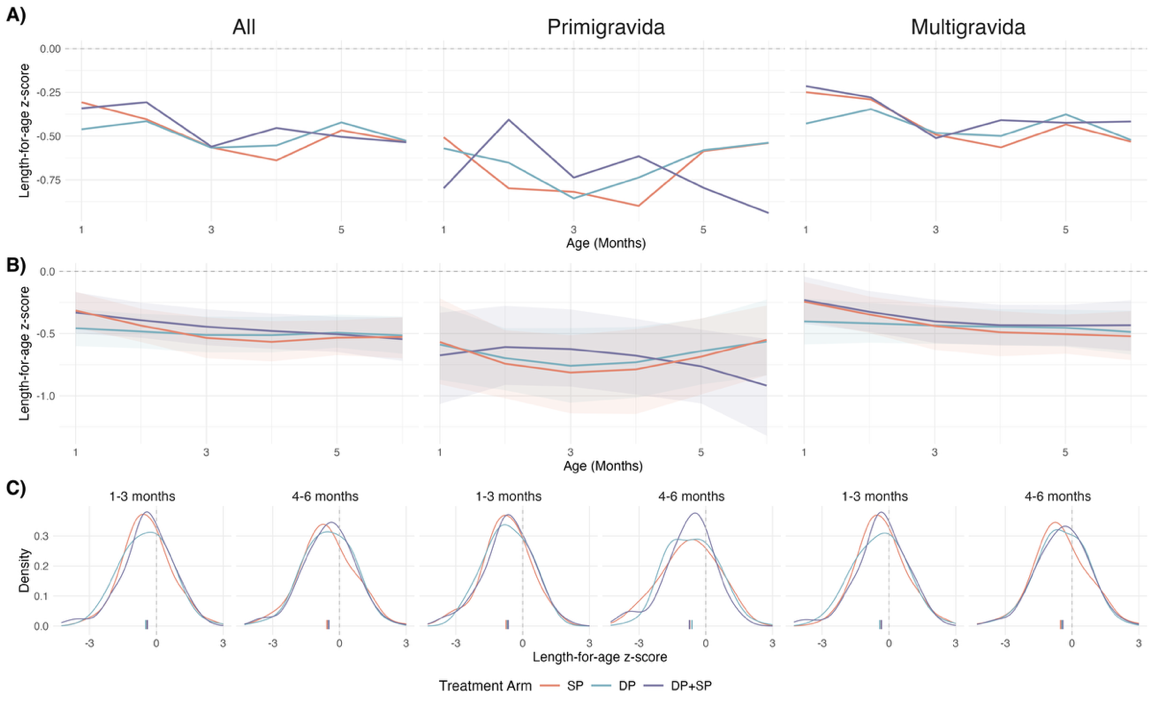

A) Unadjusted mean length-for-age z-scores (LAZ) under INTERGROWTH-21^st^ standards by age in each IPTp-arm, stratified by gravidity.

B) Mean LAZs, estimated using generalized additive models with cubic splines for age and child-specific random effects. 95% confidence intervals were estimated using a nonparametric bootstrap that resampled by child for 1,000 iterations. Models were adjusted for child sex, gravidity, birth year, birth quarter, maternal age, maternal education, maternal height, maternal weight at enrollment, household wealth index, drinking water source, sanitation facility.

C) Smoothed distributions of observed LAZs, generated through Gaussian kernel density estimation. Empirical means for each distribution are displayed as ticks on the x-axis.

*Mean LAZ By Age and IPTp Arm, Stratified by Child Sex*

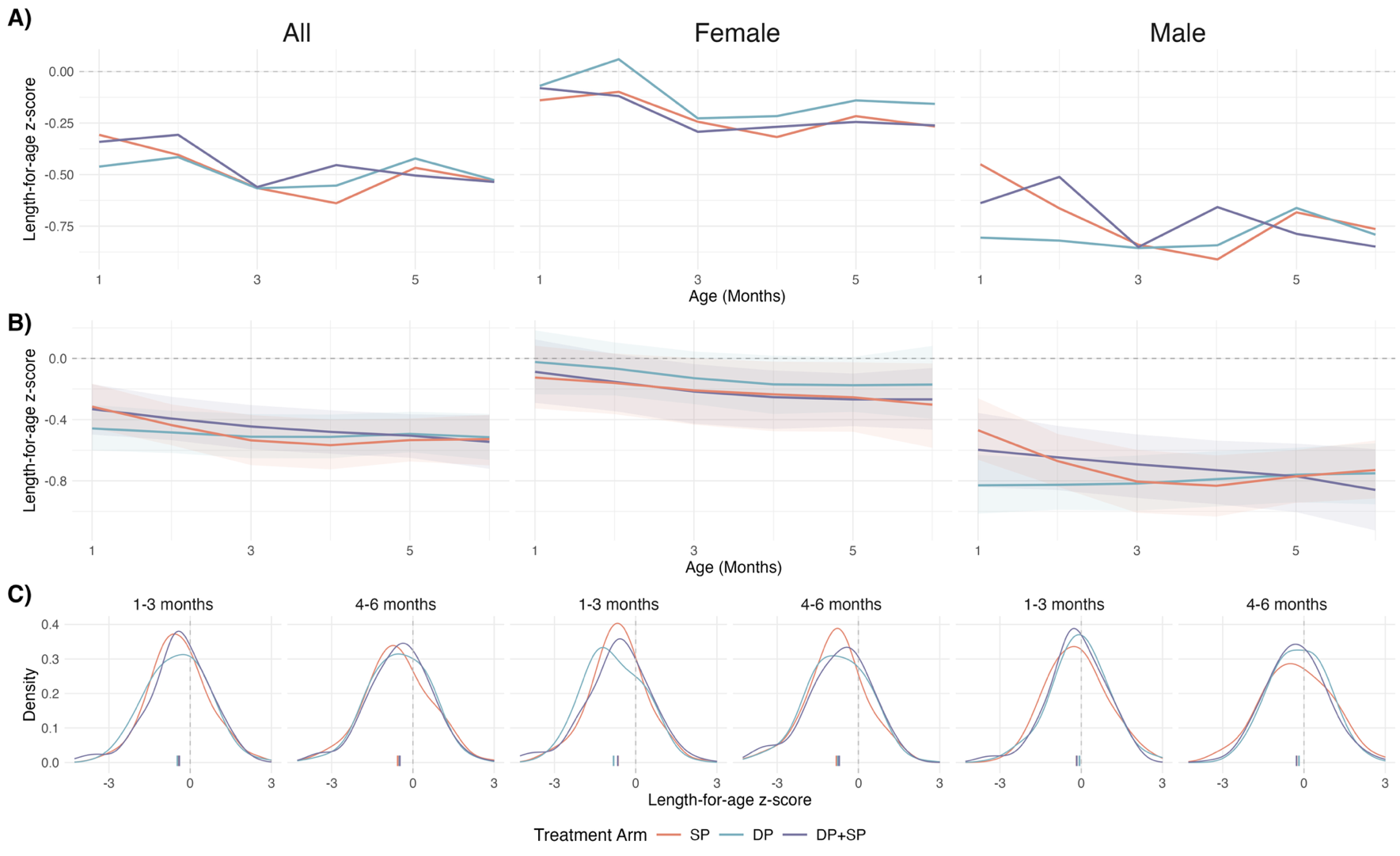

A) Unadjusted mean length-for-age z-scores (LAZ) under INTERGROWTH-21^st^ standards by age in each IPTp-arm, stratified by child sex.

B) Mean LAZs, estimated using generalized additive models with cubic splines for age and child-specific random effects. 95% confidence intervals were estimated using a nonparametric bootstrap that resampled by child for 1,000 iterations. Models were adjusted for child sex, gravidity, birth year, birth quarter, maternal age, maternal education, maternal height, maternal weight at enrollment, household wealth index, drinking water source, sanitation facility.

C) Smoothed distributions of observed LAZs, generated through Gaussian kernel density estimation. Empirical means for each distribution are displayed as ticks on the x-axis. Interventional indirect effects were only estimated if both the treatment-mediator and mediator-outcome relationships had p-values < 0.05 or large effect sizes (risk ratio < 0.9 or risk ratio > 1.1 for categorical outcomes, absolute mean difference > 0.5 for continuous outcomes).

*Total Effects of IPTp on LAZ*

|  | **Age Groups** | **Mean (SD)** | | | **Mean Difference (95% CI)** | | |
| --- | --- | --- | --- | --- | --- | --- | --- |
|  |  | DP+SP | DP | SP | DP+SP vs SP | DP+SP vs DP | DP vs SP |
| Overall | Birth | -1.34 (1.72) | -1.47 (1.82) | -1.51 (1.65) | 0.09 (-0.21, 0.40) | 0.09 (-0.22, 0.40) | 0.02 (-0.29, 0.33) |
|  | 1-3 months | -0.43 (1.12) | -0.52 (1.17) | -0.43 (1.09) | -0.08 (-0.26, 0.11) | -0.01 (-0.20, 0.18) | -0.05 (-0.24, 0.14) |
|  | 4-6 months | -0.50 (1.12) | -0.52 (1.14) | -0.55 (1.17) | 0.01 (-0.18, 0.21) | -0.05 (-0.24, 0.15) | 0.07 (-0.13, 0.26) |
| *Stratified by Gravidity* | | | | | | | |
| Primigravida | Birth | -1.61 (1.60) | -1.57 (1.70) | -1.50 (1.60) | -0.16 (-0.88, 0.55) | -0.06 (-0.70, 0.58) | 0.24 (-0.44, 0.92) |
|  | 1-3 months | -0.68 (1.08) | -0.80 (1.09) | -0.70 (1.09) | 0.24 (-0.24, 0.71) | 0.12 (-0.30, 0.55) | -0.05 (-0.47, 0.37) |
|  | 4-6 months | -0.74 (1.08) | -0.75 (1.20) | -0.67 (1.18) | 0.06 (-0.42, 0.53) | 0.02 (-0.42, 0.46) | 0.00 (-0.46, 0.47) |
| Multigravida | Birth | -1.27 (1.75) | -1.44 (1.85) | -1.51 (1.66) | 0.14 (-0.21, 0.49) | 0.09 (-0.28, 0.45) | 0.04 (-0.31, 0.39) |
|  | 1-3 months | -0.35 (1.12) | -0.43 (1.17) | -0.35 (1.08) | -0.13 (-0.34, 0.08) | -0.09 (-0.31, 0.13) | -0.07 (-0.29, 0.15) |
|  | 4-6 months | -0.43 (1.13) | -0.45 (1.11) | -0.51 (1.17) | -0.02 (-0.24, 0.20) | -0.10 (-0.32, 0.12) | 0.08 (-0.14, 0.30) |
| *Stratified by Child Sex* | | | | | | | |
| Female | Birth | -0.93 (1.59) | -1.21 (1.75) | -1.29 (1.52) | 0.26 (-0.15, 0.66) | 0.29 (-0.14, 0.72) | -0.03 (-0.47, 0.41) |
|  | 1-3 months | -0.19 (1.04) | -0.10 (1.10) | -0.18 (1.09) | -0.09 (-0.35, 0.17) | -0.11 (-0.37, 0.15) | 0.05 (-0.24, 0.33) |
|  | 4-6 months | -0.28 (1.06) | -0.19 (1.02) | -0.27 (1.19) | 0.01 (-0.26, 0.28) | -0.08 (-0.33, 0.18) | 0.05 (-0.23, 0.33) |
| Male | Birth | -1.79 (1.75) | -1.69 (1.85) | -1.70 (1.73) | -0.10 (-0.56, 0.36) | -0.07 (-0.54, 0.40) | 0.11 (-0.33, 0.56) |
|  | 1-3 months | -0.68 (1.15) | -0.86 (1.11) | -0.65 (1.04) | -0.07 (-0.34, 0.20) | 0.09 (-0.20, 0.38) | -0.12 (-0.38, 0.13) |
|  | 4-6 months | -0.74 (1.15) | -0.79 (1.16) | -0.79 (1.10) | 0.03 (-0.25, 0.31) | 0.01 (-0.29, 0.30) | 0.09 (-0.18, 0.36) |

Asterisks indicate effect estimates in which the confidence interval does not contain the null.

*Effects of Candidate Mediators on LAZ*

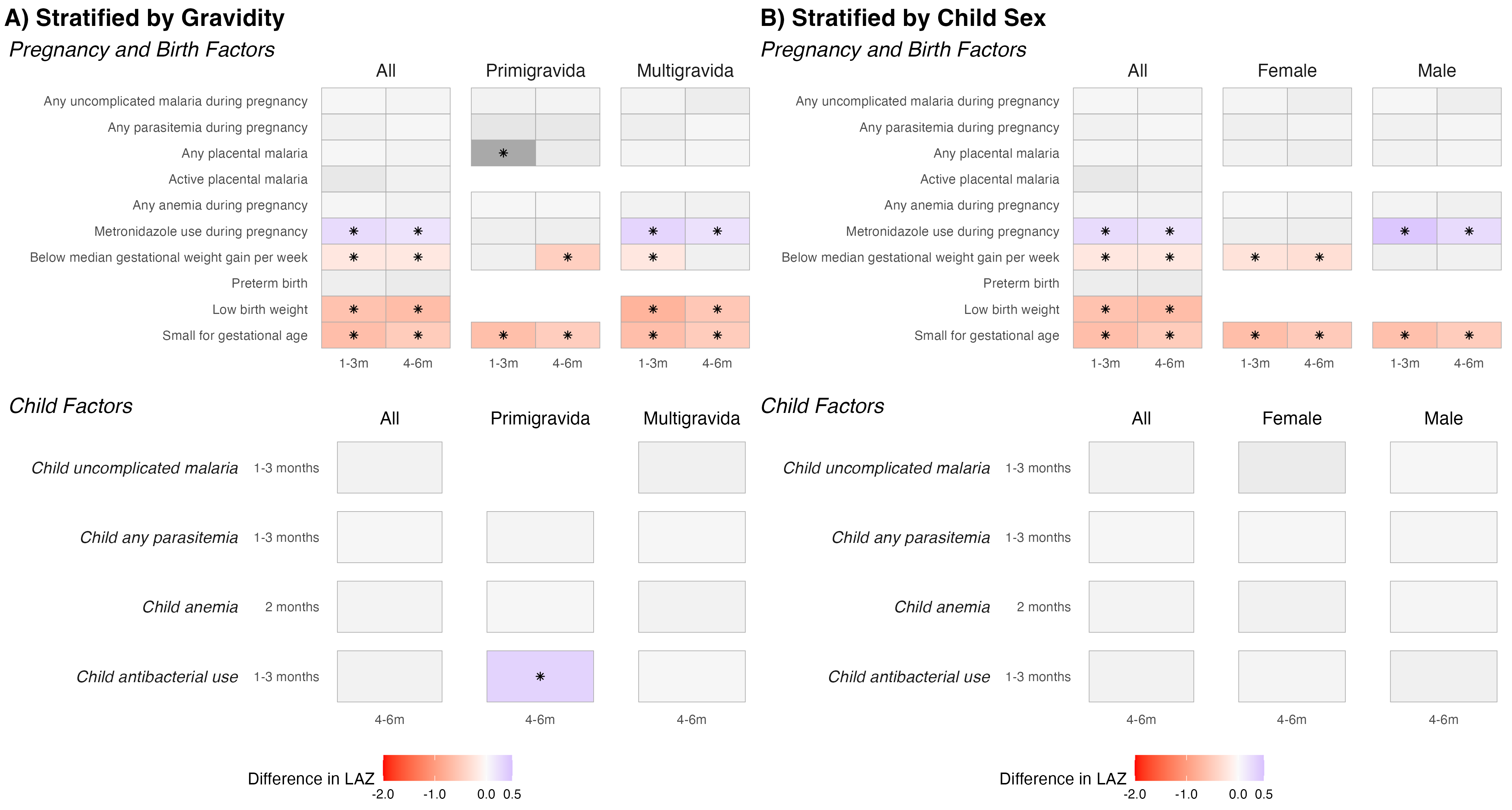

Heatmap of estimated mean differences by mediators on length-for-age z-scores (LAZs) estimated under INTERGROWTH-21^st^ standards, stratified by (A) gravidity and (B) child sex. Mean differences were estimated with generalized linear models with a Gaussian family for continuous outcomes, adjusting for baseline covariates. Tiles are shaded in color if the prevalence ratio was large (absolute mean difference > 0.5) or had p-values < 0.05. Estimates where confidence intervals did not cross the null are marked by asterisks. Effects were not estimated if there were less than 20 observed cases, or if the mediator preceded the outcome.

*Mediated Effects on LAZ*

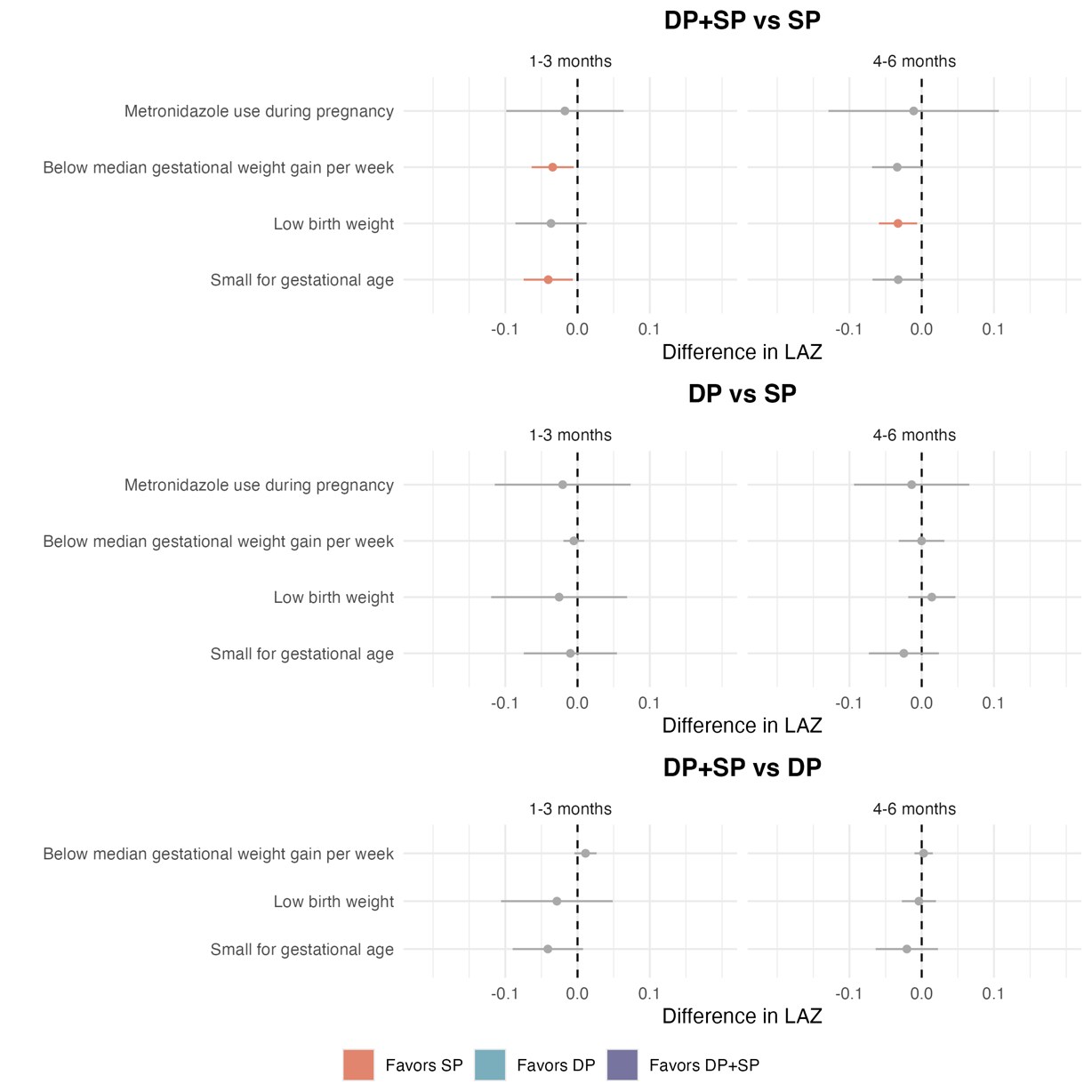

Interventional indirect effects and 95% confidence intervals of IPTp on length-for-age z-scores (LAZs) estimated under INTERGROWTH-21^st^ standards. Models were adjusted for baseline covariates, as well as intermediate confounders. Statistically significant effect estimates are displayed in color, non-significant estimates are displayed in gray. Color is determined by the comparison arm with higher LAZs (the arm “favored” to provide an increase in LAZ). Interventional indirect effects were only estimated if both the treatment-mediator and mediator-outcome relationships were large (absolute mean difference > 0.5) or had p-values below 0.05.

*Total Effects of IPTp on Incidence of First-Time Stunting*

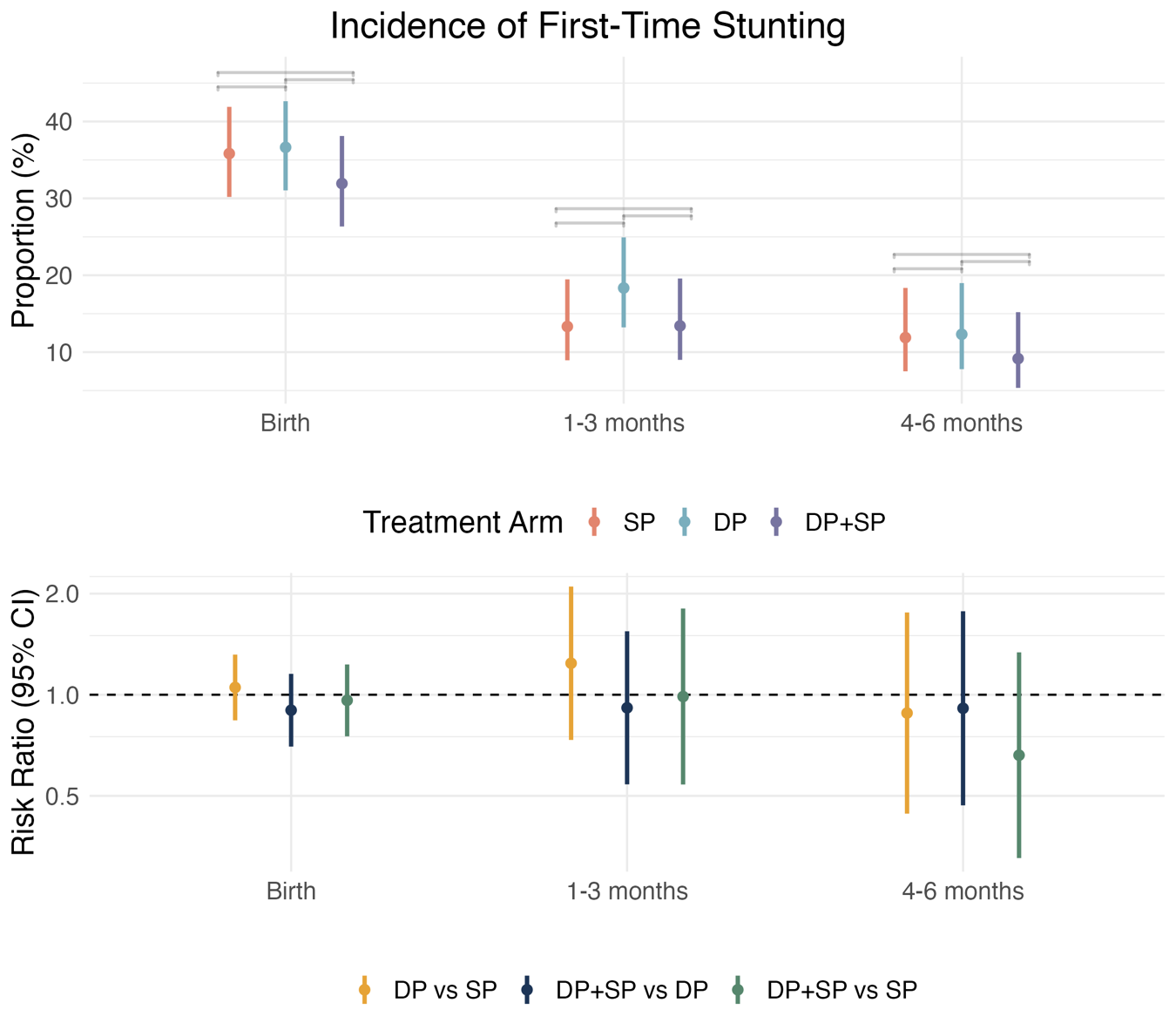

Incidence and incidence risk ratios (95% CIs) of stunting (LAZ < -2) estimated under INTERGROWTH-21^st^ standards, by age and treatment arm. Children were considered at risk of incident stunting if they had never been stunted at a prior time point. Incidence ratios were estimated with log-binomial models with robust standard errors, adjusting for baseline covariates.

*Effects of Candidate Mediators on Incidence of First-Time Stunting*

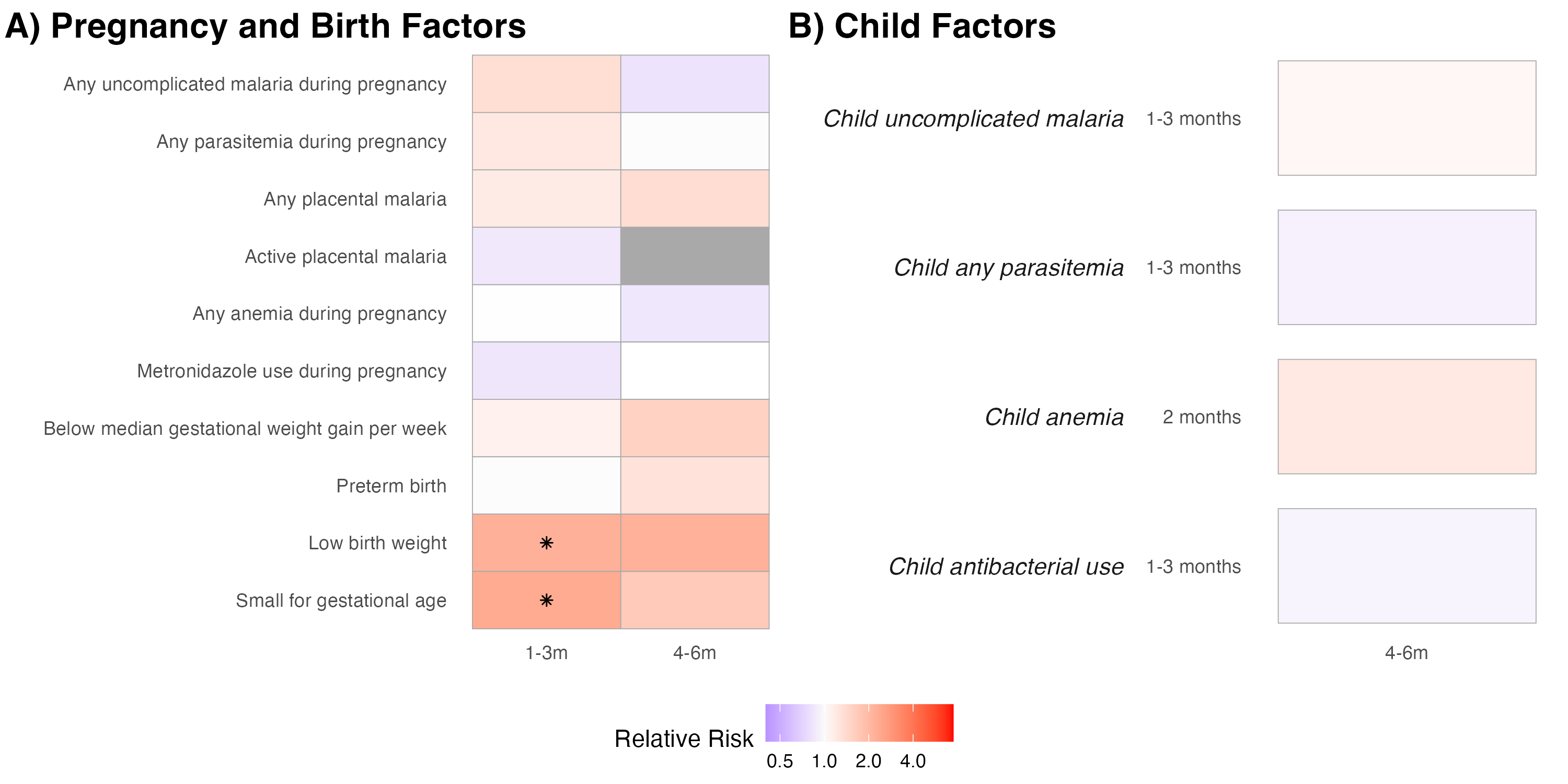

Heatmap of estimated incidence rate ratios for (A) pregnancy and birth factors and (b) infant factors measured at different ages on stunting incidence estimated under INTERGROWTH-21^st^ standards. Children were considered at risk of incident stunting if they had never been stunted at a prior time point. Incidence rate ratios were estimated with log-binomial models with robust standard errors, adjusting for baseline covariates. If log-binomial models failed to converge, we used modified Poisson regression models. Tiles are shaded in color if the incidence rate ratio is large (IRR > 1.1, IRR < 0.9) or had p-values < 0.05. Estimates where confidence intervals did not cross the null are marked by asterisks. Effects were not estimated if there were less than 20 observed cases, or if the mediator preceded the outcome.

*Mediated Effects on Incidence of First-Time Stunting*

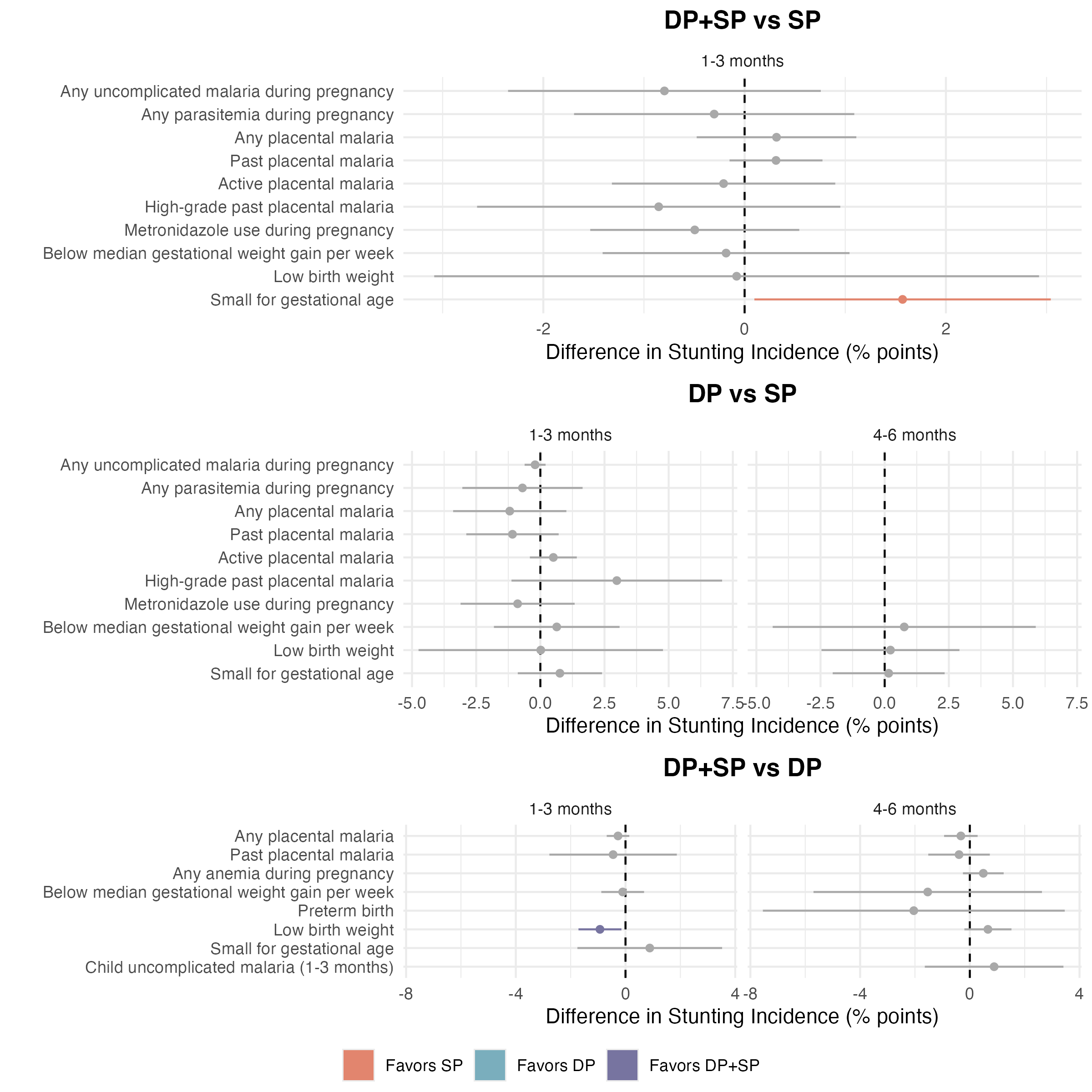

Interventional indirect effects and 95% confidence intervals of IPTp on stunting incidence (LAZ <-2) estimated under INTERGROWTH-21^st^ standards. Models were adjusted for baseline covariates, as well as intermediate confounders. Statistically significant effect estimates are displayed in color, non-significant estimates are displayed in gray. Color is determined by the comparison arm with lower risk of stunting (the arm “favored” to protect against stunting). Interventional indirect effects were only estimated if both the treatment-mediator and mediator-outcome relationships were large (risk ratio < 0.9 or risk ratio > 1.1) or had p-values below 0.05.

### Table S1: Baseline Characteristics in Original Trial vs Follow-Up Study

| **Variable** | | **SP** | | | **DP** | | | **DP+SP** | | |
| --- | --- | --- | --- | --- | --- | --- | --- | --- | --- | --- |
|  |  | Trial | Follow-Up | P-Value | Trial | Follow-Up | P-Value | Trial | Follow-Up | P-Value |
| N | | 815 | 256 | --- | 814 | 265 | --- | 830 | 240 | --- |
| Child gender | Male | 419 (51.4%) | 138 (53.9%) | 0.53 | 403 (49.5%) | 143 (54.0%) | 0.23 | 402 (48.4%) | 115 (47.9%) | 0.95 |
|  | Female | 396 (48.6%) | 118 (46.1%) |  | 411 (50.5%) | 122 (46.0%) |  | 428 (51.6%) | 125 (52.1%) |  |
| Birth year | 2021 | 104 (12.8%) | 16 (6.2%) | 0 | 106 (13.0%) | 17 (6.4%) | 0 | 109 (13.1%) | 15 (6.2%) | 0 |
|  | 2022 | 275 (33.7%) | 69 (27.0%) |  | 270 (33.2%) | 72 (27.2%) |  | 288 (34.7%) | 48 (20.0%) |  |
|  | 2023 | 288 (35.3%) | 158 (61.7%) |  | 295 (36.2%) | 168 (63.4%) |  | 290 (34.9%) | 167 (69.6%) |  |
|  | 2024 | 148 (18.2%) | 13 (5.1%) |  | 143 (17.6%) | 8 (3.0%) |  | 143 (17.2%) | 10 (4.2%) |  |
| Birth season | Jan-Mar | 229 (28.1%) | 54 (21.1%) | 0 | 217 (26.7%) | 44 (16.6%) | 0 | 214 (25.8%) | 33 (13.8%) | 0 |
|  | Apr-Jun | 199 (24.4%) | 46 (18.0%) |  | 200 (24.6%) | 38 (14.3%) |  | 208 (25.1%) | 52 (21.7%) |  |
|  | Jul-Sep | 199 (24.4%) | 73 (28.5%) |  | 210 (25.8%) | 78 (29.4%) |  | 209 (25.2%) | 62 (25.8%) |  |
|  | Oct-Dec | 188 (23.1%) | 83 (32.4%) |  | 187 (23.0%) | 105 (39.6%) |  | 199 (24.0%) | 93 (38.8%) |  |
| Birth weight (kg) | | 3.1 (0.5) | 3.1 (0.5) | 0.65 | 3.1 (0.5) | 3.1 (0.4) | 0.12 | 3.1 (0.5) | 3.1 (0.5) | 0.38 |
| Birth length (cm) | | 46.6 (3.3) | 46.7 (3.1) | 0.58 | 46.7 (3.1) | 46.9 (3.2) | 0.25 | 46.5 (3.5) | 47.0 (3.2) | 0.03 |
| Birth length-for-age z-score | | -1.6 (1.8) | -1.5 (1.6) | 0.39 | -1.5 (1.6) | -1.4 (1.7) | 0.21 | -1.6 (1.9) | -1.3 (1.7) | 0.01 |
| Birth weight-for-length z-score | | 0.9 (1.8) | 0.8 (1.7) | 0.79 | 0.6 (1.9) | 0.6 (1.9) | 0.61 | 0.7 (2.0) | 0.6 (2.0) | 0.44 |
| Born preterm | | 48 (5.9%) | 7 (2.7%) | 0.07 | 25 (3.1%) | 11 (4.2%) | 0.51 | 44 (5.3%) | 9 (3.8%) | 0.42 |
| Low birth weight | | 47 (5.8%) | 9 (3.5%) | 0.21 | 59 (7.2%) | 17 (6.4%) | 0.74 | 71 (8.6%) | 15 (6.2%) | 0.31 |
| Small-for-gestational age | | 152 (18.7%) | 52 (20.3%) | 0.62 | 206 (25.3%) | 56 (21.1%) | 0.19 | 194 (23.4%) | 58 (24.2%) | 0.87 |
| Gravidity | Primigravida | 192 (23.6%) | 57 (22.3%) | 0.73 | 203 (24.9%) | 60 (22.6%) | 0.5 | 228 (27.5%) | 52 (21.7%) | 0.09 |
|  | Multigravida | 623 (76.4%) | 199 (77.7%) |  | 611 (75.1%) | 205 (77.4%) |  | 602 (72.5%) | 188 (78.3%) |  |
| Maternal age (at birth) | | 25.7 (6.2) | 25.7 (6.3) | 0.99 | 25.9 (6.3) | 26.4 (6.4) | 0.26 | 25.3 (6.1) | 25.5 (6.0) | 0.78 |
| Maternal height at enrollment (cm) | | 159.1 (6.2) | 159.0 (5.9) | 0.73 | 159.0 (6.8) | 158.1 (7.6) | 0.09 | 159.3 (6.0) | 159.3 (6.3) | 0.95 |
| Maternal weight at enrollment (kg) | | 57.2 (9.5) | 56.6 (9.5) | 0.34 | 57.7 (9.9) | 56.9 (8.4) | 0.19 | 57.1 (8.9) | 56.8 (9.1) | 0.65 |
| Maternal gestational weight gain per week (kg/week) | | 0.2 (0.1) | 0.2 (0.1) | 0.78 | 0.2 (0.2) | 0.2 (0.2) | 0.34 | 0.2 (0.1) | 0.2 (0.1) | 0.95 |
| Maternal education level | None | 43 (5.3%) | 17 (6.6%) | 0.63 | 26 (3.2%) | 5 (1.9%) | 0.71 | 31 (3.7%) | 10 (4.2%) | 0.38 |
|  | Primary | 535 (65.6%) | 167 (65.2%) |  | 525 (64.5%) | 170 (64.2%) |  | 550 (66.3%) | 161 (67.1%) |  |
|  | Secondary | 219 (26.9%) | 69 (27.0%) |  | 244 (30.0%) | 84 (31.7%) |  | 232 (28.0%) | 68 (28.3%) |  |
|  | Higher | 18 (2.2%) | 3 (1.2%) |  | 19 (2.3%) | 6 (2.3%) |  | 17 (2.0%) | 1 (0.4%) |  |
| Household wealth | Poorest | 276 (33.9%) | 91 (35.5%) | 0.89 | 268 (32.9%) | 82 (30.9%) | 0.59 | 266 (32.0%) | 82 (34.2%) | 0.82 |
|  | Middle | 287 (35.2%) | 87 (34.0%) |  | 273 (33.5%) | 98 (37.0%) |  | 274 (33.0%) | 79 (32.9%) |  |
|  | Least Poor | 248 (30.4%) | 78 (30.5%) |  | 269 (33.0%) | 84 (31.7%) |  | 287 (34.6%) | 79 (32.9%) |  |
|  | Unknown | 4 (0.5%) | 0 (0.0%) |  | 4 (0.5%) | 1 (0.4%) |  | 3 (0.4%) |  |  |
| Household drinking water source | Unimproved | 57 (7.0%) | 24 (9.4%) | 0.27 | 43 (5.3%) | 11 (4.2%) | 0.57 | 40 (4.8%) | 9 (3.8%) | 0.59 |
|  | Improved | 754 (92.5%) | 232 (90.6%) |  | 767 (94.2%) | 253 (95.5%) |  | 787 (94.8%) | 231 (96.2%) |  |
|  | Unknown | 4 (0.5%) | 0 (0.0%) |  | 4 (0.5%) | 1 (0.4%) |  | 3 (0.4%) |  |  |
| Household sanitation facility | Uncovered Pit Latrine or No Facility | 356 (43.7%) | 128 (50.0%) | 0.1 | 333 (40.9%) | 105 (39.6%) | 0.75 | 331 (39.9%) | 112 (46.7%) | 0.08 |
|  | Flush Toliet, Ventilated Improved Pit Latrine, or Covered Pit Latrine | 455 (55.8%) | 128 (50.0%) |  | 477 (58.6%) | 159 (60.0%) |  | 496 (59.8%) | 128 (53.3%) |  |
|  | Unknown | 4 (0.5%) | 0 (0.0%) |  | 4 (0.5%) | 1 (0.4%) |  | 3 (0.4%) |  |  |
| Housing construction | Traditional | 545 (66.9%) | 183 (71.5%) | 0.23 | 552 (67.8%) | 181 (68.3%) | 0.96 | 554 (66.7%) | 167 (69.6%) | 0.5 |
|  | Modern | 266 (32.6%) | 73 (28.5%) |  | 258 (31.7%) | 83 (31.3%) |  | 273 (32.9%) | 73 (30.4%) |  |
|  | Unknown | 4 (0.5%) | 0 (0.0%) |  | 4 (0.5%) | 1 (0.4%) |  | 3 (0.4%) | 0 (0.0%) |  |

To assess potential differences between cohorts, we report p-values from t-tests for continuous variables and chi-square tests for categorical variables.

### Table S2: Effects of IPTp on Mean WLZ

|  | **Age Groups** | **Mean (SD)** | | | **Mean Difference (95% CI)** | | |
| --- | --- | --- | --- | --- | --- | --- | --- |
|  |  | DP+SP | DP | SP | DP+SP vs SP | DP+SP vs DP | DP vs SP |
| Overall | Birth | 0.57 (2.03) | 0.63 (1.93) | 0.84 (1.75) | -0.25 (-0.62, 0.11) | -0.06 (-0.43, 0.32) | -0.21 (-0.56, 0.14) |
|  | 1-3 months | 0.40 (1.23) | 0.61 (1.25) | 0.66 (1.18) | -0.18 (-0.39, 0.03) | -0.09 (-0.31, 0.12) | -0.10 (-0.31, 0.10) |
|  | 4-6 months | 0.15 (1.18) | 0.40 (1.25) | 0.49 (1.21) | *-0.28 (-0.49, -0.07) | -0.16 (-0.38, 0.05) | -0.12 (-0.33, 0.08) |
|  | 7-9 months | -0.06 (1.13) | 0.00 (1.12) | 0.06 (1.21) | -0.08 (-0.29, 0.12) | -0.00 (-0.19, 0.19) | -0.10 (-0.29, 0.10) |
|  | 10-12 months | -0.14 (1.15) | -0.09 (1.07) | -0.02 (1.25) | -0.09 (-0.30, 0.12) | 0.01 (-0.19, 0.20) | -0.13 (-0.33, 0.06) |
| *Stratified by Gravidity* | | | | | | | |
| Primigravida | Birth | 0.07 (1.75) | 0.31 (1.66) | 0.55 (1.79) | -0.18 (-1.11, 0.75) | -0.31 (-1.12, 0.49) | -0.26 (-0.99, 0.48) |
|  | 1-3 months | 0.25 (1.27) | 0.92 (1.23) | 0.91 (1.19) | *-0.98 (-1.51, -0.45) | -0.50 (-1.01, 0.00) | -0.12 (-0.59, 0.35) |
|  | 4-6 months | -0.04 (0.98) | 0.55 (1.28) | 0.78 (1.26) | *-0.99 (-1.47, -0.51) | -0.29 (-0.73, 0.16) | -0.48 (-0.97, 0.01) |
|  | 7-9 months | -0.21 (1.15) | 0.16 (1.15) | 0.35 (1.21) | *-0.76 (-1.27, -0.26) | -0.06 (-0.49, 0.37) | *-0.54 (-1.01, -0.06) |
|  | 10-12 months | -0.38 (1.22) | -0.00 (1.06) | 0.16 (1.28) | *-0.74 (-1.28, -0.21) | -0.07 (-0.49, 0.35) | -0.48 (-0.96, 0.01) |
| Multigravida | Birth | 0.69 (2.08) | 0.72 (2.00) | 0.93 (1.73) | -0.23 (-0.65, 0.20) | 0.00 (-0.44, 0.44) | -0.26 (-0.67, 0.15) |
|  | 1-3 months | 0.44 (1.22) | 0.52 (1.25) | 0.59 (1.17) | -0.04 (-0.28, 0.19) | 0.07 (-0.16, 0.31) | -0.13 (-0.35, 0.10) |
|  | 4-6 months | 0.20 (1.22) | 0.35 (1.24) | 0.40 (1.19) | -0.11 (-0.35, 0.13) | -0.08 (-0.32, 0.16) | -0.08 (-0.31, 0.15) |
|  | 7-9 months | -0.02 (1.12) | -0.04 (1.11) | -0.03 (1.19) | 0.08 (-0.14, 0.31) | 0.05 (-0.16, 0.27) | -0.04 (-0.25, 0.18) |
|  | 10-12 months | -0.07 (1.12) | -0.12 (1.08) | -0.08 (1.23) | 0.07 (-0.16, 0.30) | 0.07 (-0.15, 0.28) | -0.08 (-0.30, 0.14) |
| *Stratified by Child Sex* | | | | | | | |
| Female | Birth | 0.48 (2.02) | 0.68 (1.91) | 0.98 (1.61) | -0.36 (-0.90, 0.17) | -0.07 (-0.60, 0.46) | -0.30 (-0.82, 0.23) |
|  | 1-3 months | 0.31 (1.04) | 0.39 (1.28) | 0.63 (1.10) | -0.20 (-0.47, 0.06) | 0.01 (-0.28, 0.29) | -0.22 (-0.52, 0.07) |
|  | 4-6 months | 0.07 (1.07) | 0.24 (1.15) | 0.45 (1.10) | *-0.28 (-0.56, -0.01) | -0.10 (-0.39, 0.18) | -0.10 (-0.38, 0.18) |
|  | 7-9 months | -0.11 (1.05) | -0.01 (1.11) | 0.18 (0.99) | -0.15 (-0.41, 0.11) | -0.02 (-0.28, 0.24) | -0.10 (-0.36, 0.16) |
|  | 10-12 months | -0.12 (1.06) | -0.07 (1.07) | 0.13 (1.05) | -0.13 (-0.40, 0.13) | 0.02 (-0.25, 0.28) | -0.15 (-0.43, 0.12) |
| Male | Birth | 0.66 (2.05) | 0.59 (1.96) | 0.72 (1.86) | -0.29 (-0.81, 0.23) | -0.03 (-0.57, 0.52) | -0.22 (-0.71, 0.27) |
|  | 1-3 months | 0.50 (1.41) | 0.79 (1.20) | 0.69 (1.25) | -0.18 (-0.50, 0.14) | -0.21 (-0.53, 0.11) | 0.04 (-0.24, 0.31) |
|  | 4-6 months | 0.23 (1.28) | 0.53 (1.32) | 0.51 (1.30) | *-0.33 (-0.64, -0.02) | -0.25 (-0.58, 0.08) | -0.07 (-0.38, 0.23) |
|  | 7-9 months | -0.01 (1.21) | 0.01 (1.13) | -0.05 (1.35) | -0.03 (-0.35, 0.28) | 0.01 (-0.28, 0.30) | -0.02 (-0.31, 0.26) |
|  | 10-12 months | -0.16 (1.24) | -0.11 (1.07) | -0.15 (1.38) | -0.05 (-0.38, 0.27) | -0.03 (-0.31, 0.26) | -0.06 (-0.34, 0.23) |

Asterisks indicate effect estimates in which the confidence interval does not contain the null.

### Figure S1: Incidence and Incidence Risk Ratio of Wasting by Treatment Arm

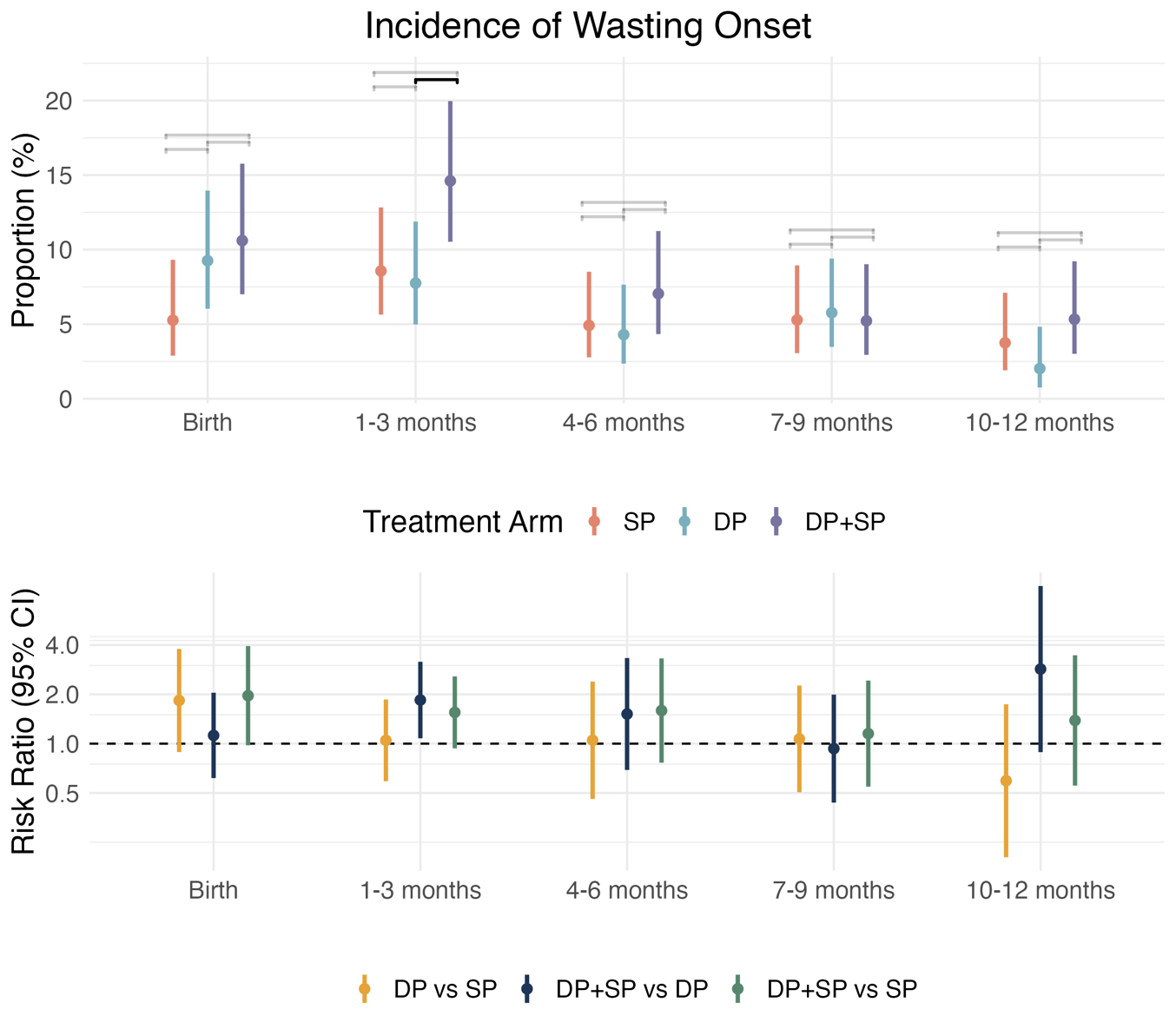

Incidence and incidence risk ratios (95% CIs) of wasting (WLZ < -2) by age and treatment arm. Children were considered at risk of incident wasting if they had not been wasted for the prior 60 days. Incidence ratios were estimated with log-binomial models with robust standard errors, adjusting for baseline covariates.

### Table S3: Effects of IPTp on Incidence of Wasting

|  | **Age Groups** | **Number of Cases (SD)** | | | **Risk Ratio (95% CI)** | | |
| --- | --- | --- | --- | --- | --- | --- | --- |
|  |  | DP+SP | DP | SP | DP+SP vs SP | DP+SP vs DP | DP vs SP |
| Overall | Birth | 21 (10.6%) | 20 (9.3%) | 11 (5.3%) | 1.96 (0.98, 3.94) | 1.12 (0.62, 2.05) | 1.83 (0.89, 3.79) |
|  | 1-3 months | 32 (14.6%) | 19 (7.8%) | 21 (8.6%) | 1.55 (0.94, 2.57) | *1.85 (1.08, 3.16) | 1.05 (0.59, 1.86) |
|  | 4-6 months | 16 (7.0%) | 11 (4.3%) | 12 (4.9%) | 1.59 (0.76, 3.31) | 1.52 (0.69, 3.33) | 1.05 (0.46, 2.39) |
|  | 7-9 months | 12 (5.2%) | 15 (5.8%) | 13 (5.3%) | 1.15 (0.55, 2.43) | 0.93 (0.44, 1.99) | 1.07 (0.50, 2.26) |
|  | 10-12 months | 12 (5.3%) | 5 (2.0%) | 9 (3.8%) | 1.39 (0.55, 3.46) | --- | --- |
| *Stratified by Gravidity* | | | | | | | |
| Primigravida | Birth | 4 (10.3%) | 4 (8.3%) | 4 (8.5%) | --- | --- | --- |
|  | 1-3 months | 8 (16.7%) | 3 (5.4%) | 4 (7.5%) | --- | --- | --- |
|  | 4-6 months | 3 (6.1%) | 1 (1.7%) | 4 (7.3%) | --- | --- | --- |
|  | 7-9 months | 3 (6.1%) | 3 (5.1%) | 2 (3.6%) | --- | --- | --- |
|  | 10-12 months | 3 (6.4%) | 2 (3.6%) | 2 (3.7%) | --- | --- | --- |
| Multigravida | Birth | 17 (10.7%) | 16 (9.5%) | 7 (4.3%) | 2.23 (0.96, 5.14) | 1.02 (0.53, 1.96) | *2.37 (1.00, 5.59) |
|  | 1-3 months | 24 (14.0%) | 16 (8.5%) | 17 (8.9%) | 1.39 (0.78, 2.45) | 1.53 (0.86, 2.72) | 1.09 (0.59, 2.02) |
|  | 4-6 months | 13 (7.3%) | 10 (5.1%) | 8 (4.2%) | 1.55 (0.68, 3.57) | 1.30 (0.58, 2.91) | --- |
|  | 7-9 months | 9 (5.0%) | 12 (6.0%) | 11 (5.8%) | 0.86 (0.38, 1.94) | 0.88 (0.39, 2.01) | 0.91 (0.41, 2.04) |
|  | 10-12 months | 9 (5.1%) | 3 (1.6%) | 7 (3.8%) | --- | --- | --- |
| *Stratified by Child Sex* | | | | | | | |
| Male | Birth | 11 (11.8%) | 12 (10.0%) | 9 (7.9%) | 1.83 (0.80, 4.21) | 1.42 (0.65, 3.14) | 1.19 (0.51, 2.79) |
|  | 1-3 months | 18 (17.3%) | 10 (7.6%) | 10 (7.8%) | 2.02 (0.92, 4.42) | 1.93 (0.94, 3.95) | 1.04 (0.45, 2.44) |
|  | 4-6 months | 9 (8.4%) | 8 (5.9%) | 7 (5.3%) | --- | --- | --- |
|  | 7-9 months | 7 (6.5%) | 10 (7.1%) | 11 (8.5%) | --- | --- | 0.75 (0.30, 1.83) |
|  | 10-12 months | 6 (5.7%) | 3 (2.3%) | 8 (6.5%) | --- | --- | --- |
| Female | Birth | 10 (9.5%) | 8 (8.3%) | 2 (2.1%) | --- | --- | --- |
|  | 1-3 months | 14 (12.2%) | 9 (7.9%) | 11 (9.5%) | 1.13 (0.54, 2.35) | 1.79 (0.82, 3.90) | 0.88 (0.38, 2.04) |
|  | 4-6 months | 7 (5.8%) | 3 (2.5%) | 5 (4.5%) | --- | --- | --- |
|  | 7-9 months | 5 (4.1%) | 5 (4.2%) | 2 (1.7%) | --- | --- | --- |
|  | 10-12 months | 6 (5.0%) | 2 (1.8%) | 1 (0.9%) | --- | --- | --- |

Asterisks indicate effect estimates in which the confidence interval does not contain the null. Effects were not estimated if there were less than 20 observed cases, as indicated with “---".

### Figure S2: Mean WLZ by Treatment Arm, Stratified by Child Sex

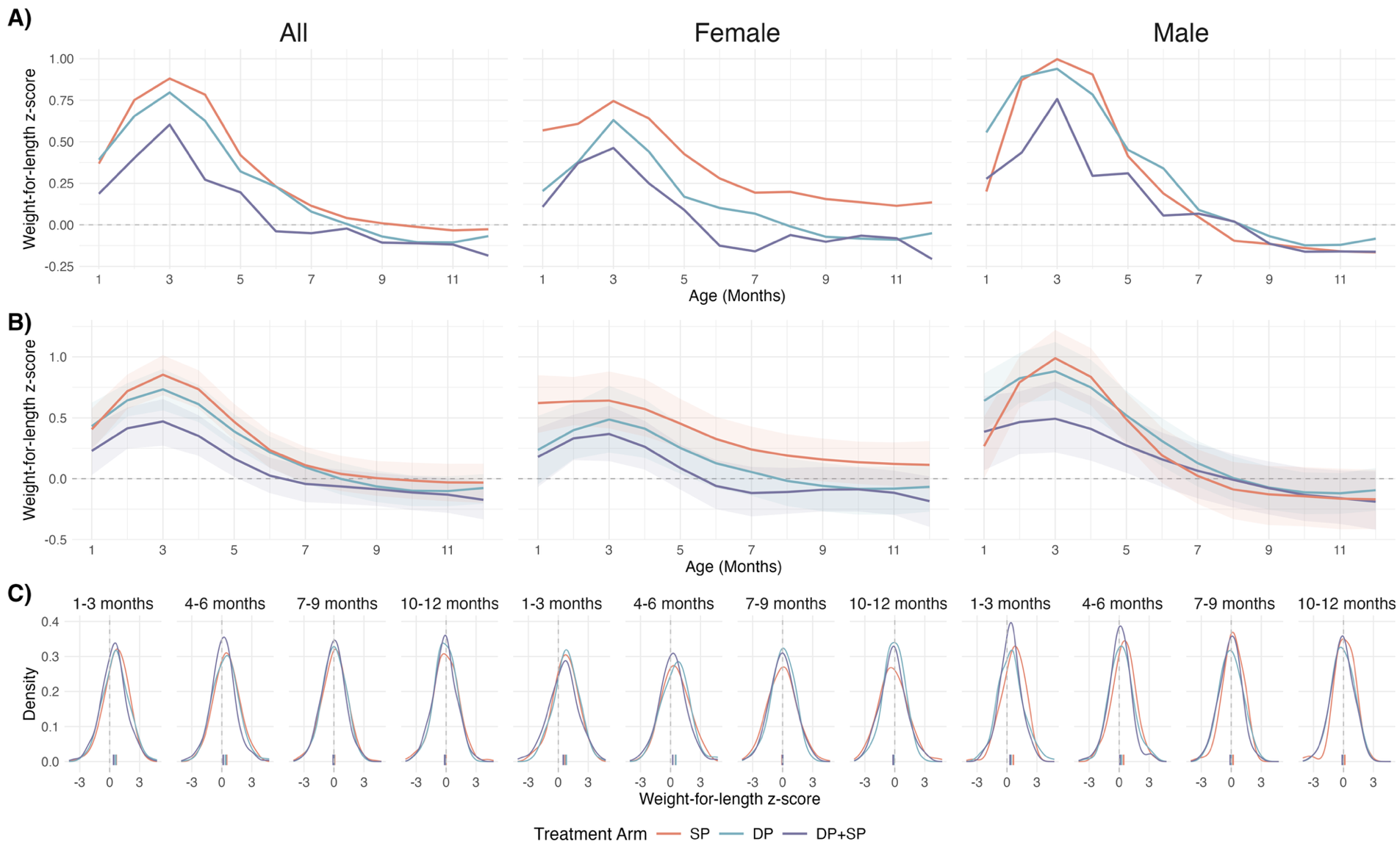

A) Unadjusted mean weight-for-length z-scores (WLZ) by age in each IPTp-arm, stratified by child sex.

B) Mean WLZs, estimated using generalized additive models with cubic splines for age and child-specific random effects. 95% confidence intervals were estimated using a nonparametric bootstrap that resampled by child for 1,000 iterations. Models were adjusted for child sex, gravidity, birth year, birth quarter, maternal age, maternal education, maternal height, maternal weight at enrollment, household wealth index, drinking water source, sanitation facility.

C) Smoothed distributions of observed WLZs, generated through Gaussian kernel density estimation. Empirical means for each distribution are displayed as ticks on the x-axis.

### Table S4: Effects of IPTp on Mean LAZ

|  | **Age Groups** | **Mean (SD)** | | | **Mean Difference (95% CI)** | | |
| --- | --- | --- | --- | --- | --- | --- | --- |
|  |  | DP+SP | DP | SP | DP+SP vs SP | DP+SP vs DP | DP vs SP |
| Overall | Birth | -1.27 (1.65) | -1.37 (1.68) | -1.45 (1.56) | 0.13 (-0.15, 0.42) | 0.05 (-0.24, 0.35) | 0.10 (-0.18, 0.38) |
|  | 1-3 months | -0.78 (1.20) | -0.81 (1.12) | -0.80 (1.10) | -0.04 (-0.23, 0.16) | -0.06 (-0.26, 0.14) | 0.02 (-0.16, 0.20) |
|  | 4-6 months | -0.77 (1.17) | -0.75 (1.15) | -0.82 (1.17) | 0.00 (-0.19, 0.20) | -0.09 (-0.29, 0.10) | 0.09 (-0.10, 0.28) |
|  | 7-9 months | -0.84 (1.22) | -0.63 (1.18) | -0.69 (1.20) | -0.18 (-0.38, 0.01) | *-0.28 (-0.48, -0.08) | 0.12 (-0.08, 0.31) |
|  | 10-12 months | -1.06 (1.19) | -0.85 (1.17) | -0.92 (1.25) | -0.17 (-0.36, 0.03) | *-0.28 (-0.47, -0.09) | 0.14 (-0.05, 0.34) |
| *Stratified by Gravidity* | | | | | | | |
| Primigravida | Birth | -1.40 (1.67) | -1.74 (1.43) | -1.80 (1.65) | 0.46 (-0.30, 1.21) | 0.49 (-0.13, 1.10) | 0.18 (-0.42, 0.79) |
|  | 1-3 months | -1.06 (1.27) | -1.18 (0.89) | -1.26 (1.13) | 0.41 (-0.12, 0.94) | 0.08 (-0.36, 0.51) | 0.08 (-0.31, 0.46) |
|  | 4-6 months | -1.06 (1.29) | -1.00 (1.06) | -1.11 (1.24) | 0.19 (-0.34, 0.72) | -0.11 (-0.55, 0.34) | 0.21 (-0.22, 0.64) |
|  | 7-9 months | -1.20 (1.36) | -0.96 (1.11) | -0.98 (1.26) | -0.04 (-0.59, 0.51) | -0.26 (-0.73, 0.21) | 0.28 (-0.14, 0.70) |
|  | 10-12 months | -1.35 (1.19) | -1.22 (1.01) | -1.05 (1.39) | -0.10 (-0.64, 0.44) | -0.15 (-0.57, 0.26) | 0.12 (-0.33, 0.57) |
| Multigravida | Birth | -1.24 (1.65) | -1.27 (1.73) | -1.35 (1.53) | 0.02 (-0.30, 0.34) | -0.08 (-0.42, 0.25) | 0.11 (-0.21, 0.43) |
|  | 1-3 months | -0.71 (1.17) | -0.71 (1.16) | -0.67 (1.06) | -0.15 (-0.37, 0.06) | -0.15 (-0.37, 0.08) | -0.02 (-0.24, 0.19) |
|  | 4-6 months | -0.69 (1.12) | -0.67 (1.17) | -0.73 (1.14) | -0.07 (-0.29, 0.14) | -0.13 (-0.35, 0.09) | 0.06 (-0.16, 0.28) |
|  | 7-9 months | -0.74 (1.17) | -0.54 (1.19) | -0.61 (1.17) | *-0.24 (-0.45, -0.02) | *-0.31 (-0.53, -0.09) | 0.09 (-0.13, 0.32) |
|  | 10-12 months | -0.98 (1.18) | -0.75 (1.19) | -0.89 (1.21) | -0.20 (-0.41, 0.02) | *-0.34 (-0.57, -0.12) | 0.16 (-0.06, 0.39) |
| *Stratified by Child Sex* | | | | | | | |
| Female | Birth | -1.11 (1.58) | -1.29 (1.64) | -1.41 (1.55) | 0.25 (-0.15, 0.64) | 0.19 (-0.22, 0.59) | 0.06 (-0.36, 0.47) |
|  | 1-3 months | -0.65 (1.11) | -0.50 (1.09) | -0.64 (1.12) | -0.05 (-0.32, 0.22) | -0.16 (-0.43, 0.11) | 0.12 (-0.16, 0.40) |
|  | 4-6 months | -0.55 (1.06) | -0.45 (0.98) | -0.54 (1.21) | 0.00 (-0.27, 0.27) | -0.09 (-0.33, 0.16) | 0.03 (-0.25, 0.31) |
|  | 7-9 months | -0.59 (1.09) | -0.44 (1.06) | -0.49 (1.08) | -0.13 (-0.38, 0.12) | -0.17 (-0.41, 0.07) | 0.04 (-0.23, 0.30) |
|  | 10-12 months | -0.83 (1.06) | -0.60 (1.04) | -0.72 (1.17) | -0.11 (-0.38, 0.16) | -0.24 (-0.48, 0.00) | 0.17 (-0.10, 0.44) |
| Male | Birth | -1.44 (1.72) | -1.45 (1.71) | -1.49 (1.58) | 0.04 (-0.38, 0.47) | -0.05 (-0.50, 0.39) | 0.19 (-0.21, 0.58) |
|  | 1-3 months | -0.93 (1.28) | -1.08 (1.07) | -0.94 (1.07) | -0.04 (-0.32, 0.25) | 0.06 (-0.24, 0.35) | -0.06 (-0.31, 0.19) |
|  | 4-6 months | -1.02 (1.23) | -1.00 (1.23) | -1.05 (1.09) | 0.01 (-0.27, 0.30) | -0.07 (-0.37, 0.24) | 0.14 (-0.13, 0.41) |
|  | 7-9 months | -1.12 (1.31) | -0.80 (1.26) | -0.86 (1.27) | -0.26 (-0.57, 0.05) | *-0.39 (-0.70, -0.07) | 0.17 (-0.12, 0.46) |
|  | 10-12 months | -1.31 (1.28) | -1.07 (1.23) | -1.10 (1.29) | -0.25 (-0.55, 0.05) | -0.31 (-0.62, 0.00) | 0.15 (-0.14, 0.45) |

Asterisks indicate effect estimates in which the confidence interval does not contain the null.

### Figure S3: Mean LAZ at 7-13 Months by Treatment Arm, Stratified by Wasting Status at 0-6 Months

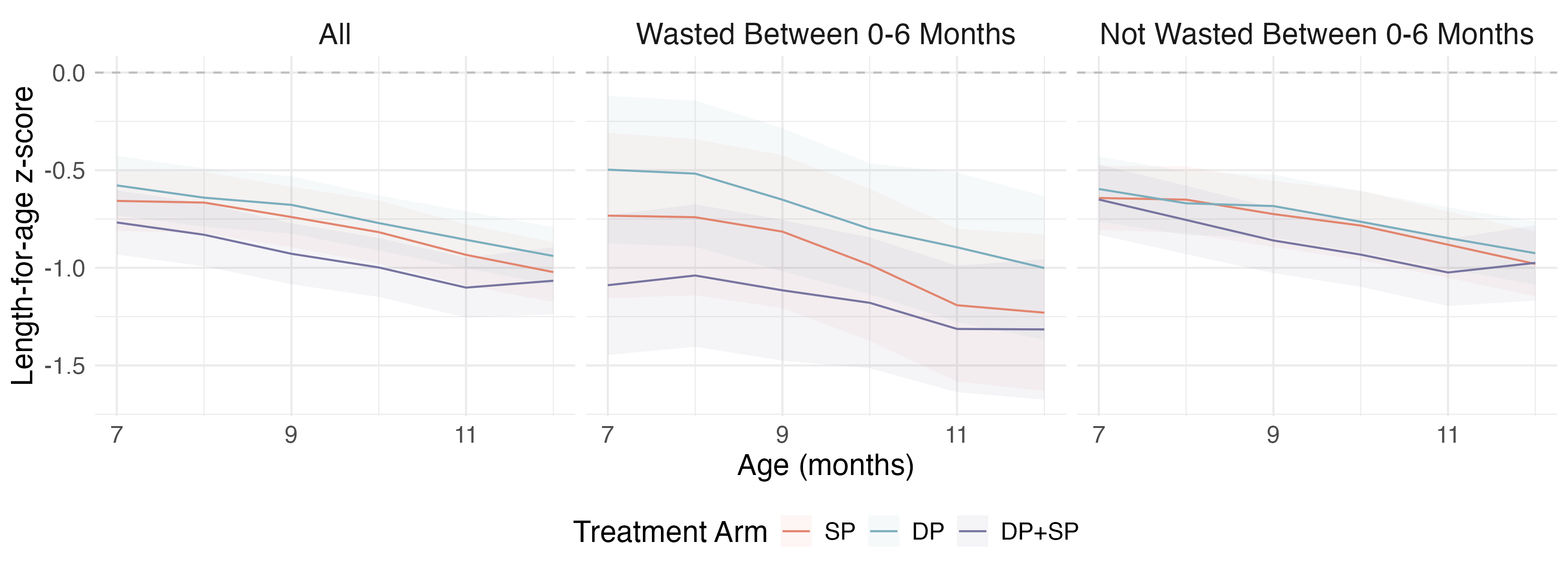

Unadjusted mean length-for-age z-scores (LAZ) by age between 7-13 months in each IPTp-arm, stratified by wasting status in the first 6 months of life. 95% CIs were estimated using standards errors under a normal distribution.

### Figure S4: Mean LAZ by Treatment Arm, Stratified by Child Sex

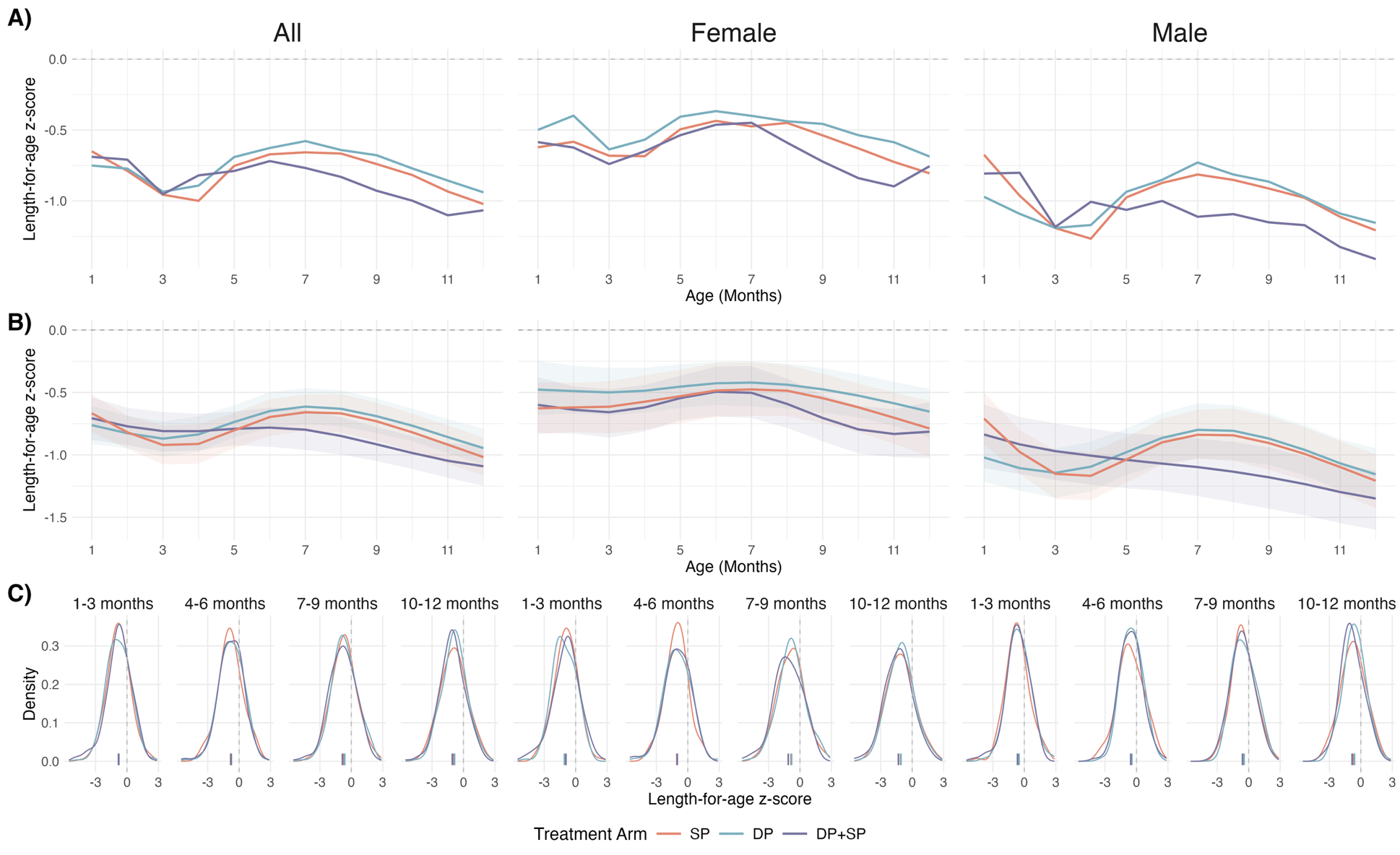

A) Unadjusted mean length-for-age z-scores (LAZ) by age in each IPTp-arm, stratified by child sex.

B) Mean LAZs, estimated using generalized additive models with cubic splines for age and child-specific random effects. 95% confidence intervals were estimated using a nonparametric bootstrap that resampled by child for 1,000 iterations. Models were adjusted for child sex, gravidity, birth year, birth quarter, maternal age, maternal education, maternal height, maternal weight at enrollment, household wealth index, drinking water source, sanitation facility.

C) Smoothed distributions of observed LAZ s, generated through Gaussian kernel density estimation. Empirical means for each distribution are displayed as ticks on the x-axis.

### Figure S5: Incidence and Incidence Risk Ratio for Stunting by Treatment Arm

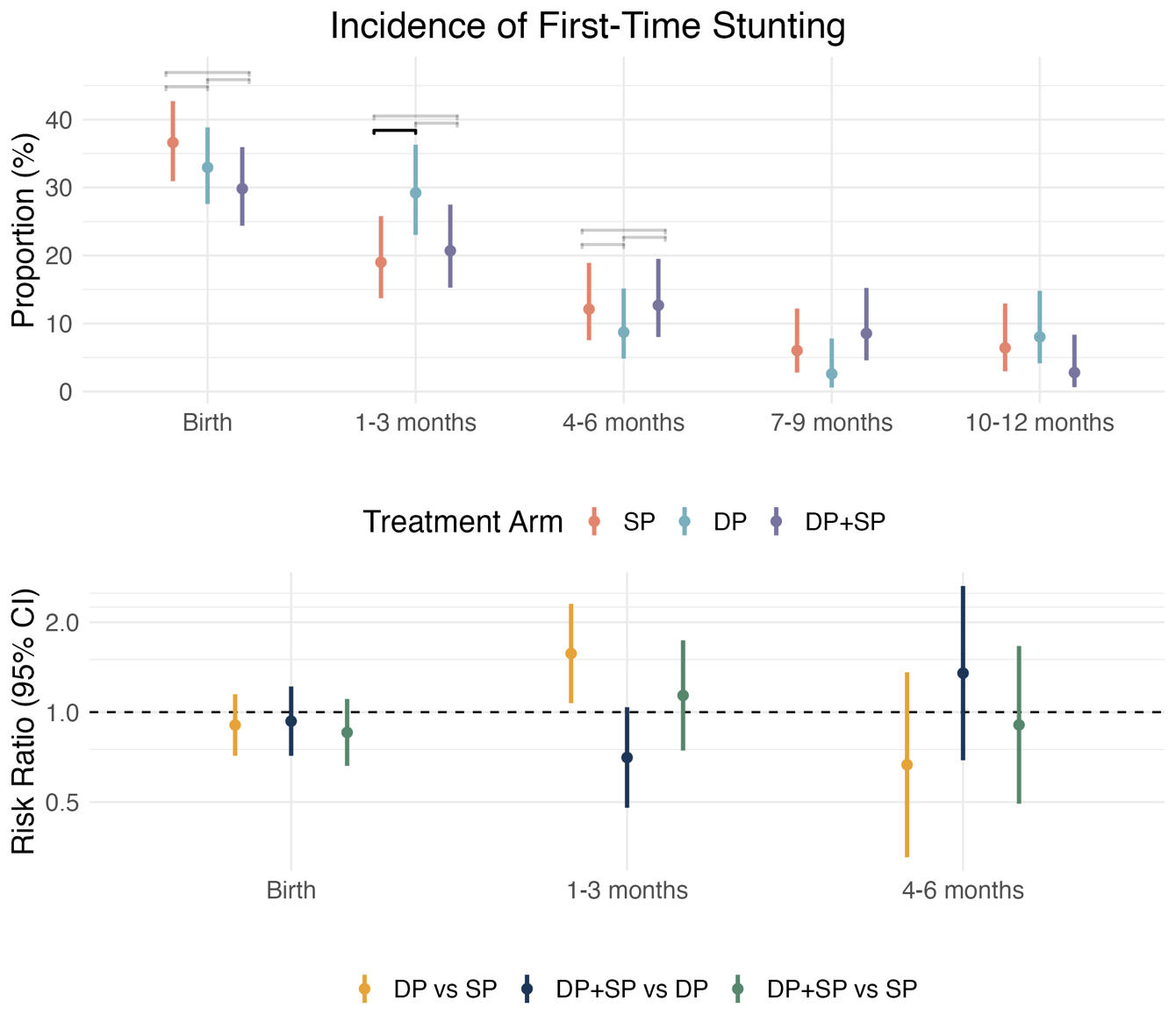

Incidence and incidence risk ratios (95% CIs) of stunting (LAZ < -2) by age and treatment arm. Children were considered at risk of incident stunting if they had never been stunted at a prior time point. Incidence ratios were estimated with log-binomial models with robust standard errors, adjusting for baseline covariates.

### Table S5: Effects of IPTp on Incidence of First-Time Stunting

|  | **Age Groups** | **Number of Cases (SD)** | | | **Risk Ratio (95% CI)** | | |
| --- | --- | --- | --- | --- | --- | --- | --- |
|  |  | DP+SP | DP | SP | DP+SP vs SP | DP+SP vs DP | DP vs SP |
| Overall | Birth | 71 (29.8%) | 87 (33.0%) | 93 (36.6%) | 0.86 (0.66, 1.11) | 0.93 (0.71, 1.22) | 0.91 (0.71, 1.15) |
|  | 1-3 months | 35 (20.7%) | 52 (29.2%) | 31 (19.0%) | 1.14 (0.74, 1.74) | 0.70 (0.48, 1.04) | *1.57 (1.07, 2.30) |
|  | 4-6 months | 17 (12.7%) | 11 (8.7%) | 16 (12.1%) | 0.91 (0.49, 1.67) | 1.35 (0.69, 2.64) | 0.67 (0.33, 1.36) |
|  | 7-9 months | 10 (8.5%) | 3 (2.6%) | 7 (6.0%) | --- | --- | --- |
|  | 10-12 months | 3 (2.8%) | 9 (8.0%) | 7 (6.4%) | --- | --- | --- |
| *Stratified by Gravidity* | | | | | | | |
| Primigravida | Birth | 17 (34.0%) | 25 (41.7%) | 24 (42.1%) | 1.14 (0.69, 1.88) | 0.82 (0.48, 1.41) | 1.15 (0.74, 1.79) |
|  | 1-3 months | 11 (31.4%) | 16 (45.7%) | 11 (33.3%) | 0.61 (0.24, 1.55) | 0.70 (0.31, 1.60) | 1.16 (0.64, 2.10) |
|  | 4-6 months | 3 (12.5%) | 1 (5.3%) | 3 (13.6%) | --- | --- | --- |
|  | 7-9 months | 3 (14.3%) | 1 (5.6%) | 2 (10.5%) | --- | --- | --- |
|  | 10-12 months | 0 (0.0%) | 2 (11.8%) | 1 (5.9%) | --- | --- | --- |
| Multigravida | Birth | 54 (28.7%) | 62 (30.4%) | 69 (35.0%) | 0.89 (0.66, 1.22) | 1.03 (0.75, 1.42) | 0.86 (0.65, 1.13) |
|  | 1-3 months | 24 (17.9%) | 36 (25.2%) | 20 (15.4%) | 1.44 (0.80, 2.60) | 0.90 (0.55, 1.47) | *1.66 (1.01, 2.73) |
|  | 4-6 months | 14 (12.7%) | 10 (9.3%) | 13 (11.8%) | 0.99 (0.49, 1.98) | 1.39 (0.65, 2.97) | 0.71 (0.33, 1.50) |
|  | 7-9 months | 7 (7.3%) | 2 (2.1%) | 5 (5.2%) | --- | --- | --- |
|  | 10-12 months | 3 (3.4%) | 7 (7.4%) | 6 (6.5%) | --- | --- | --- |
| *Stratified by Child Sex* | | | | | | | |
| Male | Birth | 40 (35.4%) | 56 (39.2%) | 50 (36.5%) | 0.98 (0.70, 1.38) | 0.85 (0.61, 1.18) | 0.98 (0.72, 1.33) |
|  | 1-3 months | 21 (28.0%) | 34 (39.1%) | 22 (25.0%) | 1.23 (0.70, 2.14) | 0.83 (0.51, 1.35) | 1.41 (0.91, 2.18) |
|  | 4-6 months | 8 (14.8%) | 7 (13.2%) | 12 (18.2%) | 0.77 (0.34, 1.71) | --- | --- |
|  | 7-9 months | 6 (13.0%) | 2 (4.3%) | 5 (9.3%) | --- | --- | --- |
|  | 10-12 months | 1 (2.5%) | 6 (13.6%) | 4 (8.2%) | --- | --- | --- |
| Female | Birth | 31 (24.8%) | 31 (25.6%) | 43 (36.8%) | 0.72 (0.48, 1.07) | 0.97 (0.62, 1.51) | 0.79 (0.53, 1.17) |
|  | 1-3 months | 14 (14.9%) | 18 (19.8%) | 9 (12.0%) | 1.20 (0.48, 2.97) | *0.54 (0.29, 1.00) | 1.60 (0.76, 3.38) |
|  | 4-6 months | 9 (11.2%) | 4 (5.5%) | 4 (6.1%) | --- | --- | --- |
|  | 7-9 months | 4 (5.6%) | 1 (1.4%) | 2 (3.2%) | --- | --- | --- |
|  | 10-12 months | 2 (3.0%) | 3 (4.4%) | 3 (5.0%) | --- | --- | --- |

Asterisks indicate effect estimates in which the confidence interval does not contain the null. Effects were not estimated if there were less than 20 observed cases, as indicated with “---".

### Table S6: Treatment-Mediator Effects

|  | **Mediator** | **Mediator Age** | **Number of Cases (SD)** | | | **Risk Ratio (95% CI)** | | |
| --- | --- | --- | --- | --- | --- | --- | --- | --- |
|  |  |  | DP+SP | DP | SP | DP+SP vs SP | DP+SP vs DP | DP vs SP |
| Overall | Any uncomplicated malaria during pregnancy | | 10 (4.2%) | 9 (3.4%) | 52 (20.3%) | *0.21 (0.16, 0.28) | --- | *0.17 (0.12, 0.23) |
|  | Any complicated malaria during pregnancy | | 0 (0.0%) | 0 (0.0%) | 0 (0.0%) | --- | --- | --- |
|  | Any parasitemia during pregnancy | | 208 (86.7%) | 221 (83.4%) | 231 (90.2%) | *0.95 (0.92, 0.97) | 1.02 (0.99, 1.06) | *0.93 (0.90, 0.96) |
|  | Any placental malaria | | 117 (51.1%) | 111 (42.9%) | 163 (67.1%) | *0.74 (0.69, 0.79) | *1.15 (1.07, 1.25) | *0.65 (0.60, 0.69) |
|  | Past placental malaria | | 116 (50.7%) | 110 (42.5%) | 162 (66.7%) | *0.74 (0.69, 0.79) | *1.16 (1.07, 1.25) | *0.64 (0.60, 0.69) |
|  | Active placental malaria | | 3 (1.3%) | 3 (1.2%) | 24 (9.9%) | *0.13 (0.08, 0.24) | --- | *0.12 (0.07, 0.21) |
|  | High-grade past placental malaria | | 0 (0.0%) | 1 (0.9%) | 24 (14.8%) | *0.00 (0.00, 0.00) | --- | *0.05 (0.02, 0.12) |
|  | Any anemia during pregnancy | | 139 (57.9%) | 129 (48.7%) | 168 (65.6%) | *0.89 (0.84, 0.95) | *1.16 (1.08, 1.26) | *0.76 (0.71, 0.82) |
|  | Any severe anemia during pregnancy | | 4 (1.7%) | 1 (0.4%) | 7 (2.7%) | --- | --- | --- |
|  | Azithromycin use during pregnancy | | 2 (0.8%) | 4 (1.5%) | 0 (0.0%) | --- | --- | --- |
|  | Tetracycline use during pregnancy | | 1 (0.4%) | 0 (0.0%) | 3 (1.2%) | --- | --- | --- |
|  | Doxycycline use during pregnancy | | 0 (0.0%) | 0 (0.0%) | 1 (0.4%) | --- | --- | --- |
|  | Metronidazole use during pregnancy | | 78 (32.5%) | 91 (34.3%) | 109 (42.6%) | *0.76 (0.69, 0.85) | 0.95 (0.85, 1.06) | *0.80 (0.73, 0.89) |
|  | Ferrous use during pregnancy | | 240 (100.0%) | 265 (100.0%) | 256 (100.0%) | 1.00 (1.00, 1.00) | 1.00 (1.00, 1.00) | 1.00 (1.00, 1.00) |
|  | Below median gestational weight gain per week | | 118 (50.9%) | 138 (52.9%) | 100 (40.0%) | *1.22 (1.12, 1.33) | *0.91 (0.85, 0.98) | *1.30 (1.20, 1.42) |
|  | Preterm birth | | 9 (3.8%) | 11 (4.2%) | 7 (2.7%) | --- | 1.11 (0.78, 1.60) | --- |
|  | Low birth weight | | 15 (6.2%) | 17 (6.4%) | 9 (3.5%) | *1.82 (1.20, 2.75) | 1.10 (0.78, 1.55) | *1.82 (1.25, 2.65) |
|  | Small for gestational age | | 58 (24.2%) | 56 (21.1%) | 52 (20.3%) | *1.33 (1.14, 1.55) | *1.26 (1.08, 1.46) | 1.13 (0.97, 1.32) |
|  | Child uncomplicated malaria | 1-3 months | 27 (11.2%) | 24 (9.1%) | 18 (7.0%) | *1.79 (1.38, 2.32) | 1.12 (0.88, 1.42) | *1.41 (1.06, 1.87) |
|  |  | 4-6 months | 47 (19.6%) | 44 (16.6%) | 45 (17.6%) | *1.28 (1.09, 1.49) | *1.26 (1.07, 1.48) | 1.00 (0.85, 1.17) |
|  |  | 7-9 months | 41 (17.1%) | 62 (23.4%) | 47 (18.4%) | 1.05 (0.89, 1.23) | *0.71 (0.61, 0.83) | *1.53 (1.32, 1.77) |
|  | Child complicated malaria | 1-3 months | 0 (0.0%) | 0 (0.0%) | 0 (0.0%) | --- | --- | --- |
|  |  | 4-6 months | 1 (0.4%) | 1 (0.4%) | 1 (0.4%) | --- | --- | --- |
|  |  | 7-9 months | 1 (0.4%) | 3 (1.1%) | 1 (0.4%) | --- | --- | --- |
|  | Child any parasitemia | 1-3 months | 49 (20.4%) | 55 (20.8%) | 46 (18.0%) | 1.17 (1.00, 1.38) | 0.96 (0.82, 1.13) | *1.22 (1.04, 1.43) |
|  |  | 4-6 months | 94 (39.2%) | 93 (35.1%) | 95 (37.1%) | *1.14 (1.03, 1.25) | *1.16 (1.05, 1.28) | 1.00 (0.90, 1.11) |
|  |  | 7-9 months | 81 (33.8%) | 112 (42.3%) | 96 (37.5%) | 0.98 (0.89, 1.08) | *0.88 (0.79, 0.97) | *1.20 (1.09, 1.32) |
|  | Child anemia | 2 months | 83 (60.1%) | 105 (61.8%) | 104 (61.5%) | 0.99 (0.91, 1.07) | 0.96 (0.88, 1.04) | 0.98 (0.91, 1.06) |
|  |  | 6 months | 98 (71.0%) | 116 (68.2%) | 111 (65.7%) | *1.13 (1.06, 1.21) | *1.08 (1.01, 1.15) | 1.02 (0.95, 1.09) |
|  |  | 13 months | 107 (77.5%) | 126 (74.1%) | 120 (71.0%) | *1.14 (1.08, 1.21) | 1.04 (0.99, 1.11) | 1.04 (0.98, 1.11) |
|  | Child severe anemia | 2 months | 0 (0.0%) | 6 (3.5%) | 1 (0.6%) | --- | --- | --- |
|  |  | 6 months | 9 (6.5%) | 10 (5.9%) | 5 (3.0%) | --- | --- | --- |
|  |  | 13 months | 3 (2.2%) | 12 (7.1%) | 5 (3.0%) | --- | --- | --- |
|  | Child antimalarial use | 1-3 months | 25 (10.6%) | 24 (9.3%) | 18 (7.1%) | *1.63 (1.25, 2.11) | 1.03 (0.81, 1.31) | *1.43 (1.08, 1.89) |
|  |  | 4-6 months | 49 (20.8%) | 45 (17.5%) | 48 (19.0%) | *1.25 (1.07, 1.45) | *1.31 (1.12, 1.54) | 0.95 (0.81, 1.11) |
|  |  | 7-9 months | 42 (17.8%) | 65 (25.3%) | 51 (20.2%) | 0.98 (0.84, 1.15) | *0.69 (0.59, 0.81) | *1.47 (1.28, 1.68) |
|  | Child antiparasitic use | 1-3 months | 0 (0.0%) | 0 (0.0%) | 0 (0.0%) | --- | --- | --- |
|  |  | 4-6 months | 2 (0.8%) | 0 (0.0%) | 1 (0.4%) | --- | --- | --- |
|  |  | 7-9 months | 0 (0.0%) | 1 (0.4%) | 1 (0.4%) | --- | --- | --- |
|  | Child antiviral use | 1-3 months | 0 (0.0%) | 1 (0.4%) | 0 (0.0%) | --- | --- | --- |
|  |  | 4-6 months | 0 (0.0%) | 0 (0.0%) | 2 (0.8%) | --- | --- | --- |
|  |  | 7-9 months | 0 (0.0%) | 1 (0.4%) | 0 (0.0%) | --- | --- | --- |
|  | Child antibacterial use | 1-3 months | 151 (64.0%) | 148 (57.6%) | 157 (62.1%) | 1.05 (0.99, 1.12) | *1.09 (1.02, 1.16) | 0.94 (0.88, 1.01) |
|  |  | 4-6 months | 185 (78.4%) | 186 (72.4%) | 194 (76.7%) | 1.04 (0.99, 1.08) | *1.11 (1.06, 1.17) | *0.93 (0.89, 0.98) |
|  |  | 7-9 months | 166 (70.3%) | 184 (71.6%) | 173 (68.4%) | 1.04 (0.99, 1.10) | 0.99 (0.94, 1.05) | *1.05 (1.00, 1.11) |
|  | Child beta-lactam use | 1-3 months | 143 (60.6%) | 141 (54.9%) | 148 (58.5%) | 1.06 (0.99, 1.13) | *1.08 (1.01, 1.16) | 0.95 (0.89, 1.02) |
|  |  | 4-6 months | 174 (73.7%) | 181 (70.4%) | 188 (74.3%) | 1.01 (0.96, 1.06) | *1.08 (1.03, 1.13) | *0.93 (0.89, 0.98) |
|  |  | 7-9 months | 154 (65.3%) | 177 (68.9%) | 162 (64.0%) | 1.03 (0.97, 1.09) | 0.96 (0.90, 1.01) | *1.08 (1.02, 1.14) |
|  | Child fluoroquinolone use | 1-3 months | 0 (0.0%) | 0 (0.0%) | 2 (0.8%) | --- | --- | --- |
|  |  | 4-6 months | 0 (0.0%) | 2 (0.8%) | 1 (0.4%) | --- | --- | --- |
|  |  | 7-9 months | 0 (0.0%) | 3 (1.2%) | 2 (0.8%) | --- | --- | --- |
|  | Child sulfonamide use | 1-3 months | 0 (0.0%) | 0 (0.0%) | 0 (0.0%) | --- | --- | --- |
|  |  | 4-6 months | 0 (0.0%) | 0 (0.0%) | 0 (0.0%) | --- | --- | --- |
|  |  | 7-9 months | 0 (0.0%) | 0 (0.0%) | 0 (0.0%) | --- | --- | --- |
|  | Child other antibacterial use | 1-3 months | 1 (0.4%) | 3 (1.2%) | 3 (1.2%) | --- | --- | --- |
|  |  | 4-6 months | 2 (0.8%) | 7 (2.7%) | 12 (4.7%) | --- | --- | --- |
|  |  | 7-9 months | 1 (0.4%) | 4 (1.6%) | 9 (3.6%) | --- | --- | --- |
| *Strratified by Gravidity* | | | | | | | | |
| Primigravida | Any uncomplicated malaria during pregnancy | | 4 (7.7%) | 4 (6.7%) | 28 (49.1%) | *0.14 (0.09, 0.22) | --- | *0.12 (0.08, 0.19) |
|  | Any complicated malaria during pregnancy | | 0 (0.0%) | 0 (0.0%) | 0 (0.0%) | --- | --- | --- |
|  | Any parasitemia during pregnancy | | 46 (88.5%) | 51 (85.0%) | 55 (96.5%) | *0.95 (0.90, 0.99) | 1.04 (0.99, 1.10) | *0.94 (0.90, 0.98) |
|  | Any placental malaria | | 39 (78.0%) | 48 (80.0%) | 47 (92.2%) | *0.92 (0.86, 0.98) | 1.02 (0.95, 1.11) | *0.93 (0.87, 1.00) |
|  | Past placental malaria | | 39 (78.0%) | 47 (78.3%) | 47 (92.2%) | *0.92 (0.86, 0.98) | 1.06 (0.98, 1.15) | *0.91 (0.84, 0.97) |
|  | Active placental malaria | | 2 (4.0%) | 2 (3.3%) | 11 (21.6%) | --- | --- | --- |
|  | High-grade past placental malaria | | 0 (0.0%) | 1 (2.1%) | 17 (36.2%) | --- | --- | --- |
|  | Any anemia during pregnancy | | 27 (51.9%) | 31 (51.7%) | 50 (87.7%) | *0.59 (0.52, 0.68) | 0.92 (0.78, 1.08) | *0.60 (0.53, 0.68) |
|  | Any severe anemia during pregnancy | | 1 (1.9%) | 0 (0.0%) | 4 (7.0%) | --- | --- | --- |
|  | Azithromycin use during pregnancy | | 1 (1.9%) | 2 (3.3%) | 0 (0.0%) | --- | --- | --- |
|  | Tetracycline use during pregnancy | | 1 (1.9%) | 0 (0.0%) | 0 (0.0%) | --- | --- | --- |
|  | Doxycycline use during pregnancy | | 0 (0.0%) | 0 (0.0%) | 0 (0.0%) | --- | --- | --- |
|  | Metronidazole use during pregnancy | | 24 (46.2%) | 23 (38.3%) | 29 (50.9%) | 0.94 (0.77, 1.15) | 1.06 (0.87, 1.29) | 0.83 (0.68, 1.01) |
|  | Ferrous use during pregnancy | | 52 (100.0%) | 60 (100.0%) | 57 (100.0%) | 1.00 (1.00, 1.00) | 1.00 (1.00, 1.00) | 1.00 (1.00, 1.00) |
|  | Below median gestational weight gain per week | | 27 (55.1%) | 34 (56.7%) | 25 (44.6%) | 1.04 (0.86, 1.26) | *0.76 (0.65, 0.88) | *1.30 (1.09, 1.55) |
|  | Preterm birth | | 3 (5.8%) | 2 (3.3%) | 4 (7.0%) | --- | --- | --- |
|  | Low birth weight | | 7 (13.5%) | 5 (8.3%) | 6 (10.5%) | --- | --- | --- |
|  | Small for gestational age | | 17 (32.7%) | 18 (30.0%) | 16 (28.1%) | 1.16 (0.85, 1.58) | 1.07 (0.81, 1.42) | 1.18 (0.90, 1.53) |
|  | Child uncomplicated malaria | 1-3 months | 5 (9.6%) | 3 (5.0%) | 4 (7.0%) | --- | --- | --- |
|  |  | 4-6 months | 14 (26.9%) | 9 (15.0%) | 14 (24.6%) | 1.18 (0.85, 1.66) | *3.41 (2.24, 5.19) | *0.63 (0.42, 0.94) |
|  |  | 7-9 months | 12 (23.1%) | 10 (16.7%) | 12 (21.1%) | 0.90 (0.66, 1.22) | 1.16 (0.79, 1.70) | 1.16 (0.82, 1.66) |
|  | Child complicated malaria | 1-3 months | 0 (0.0%) | 0 (0.0%) | 0 (0.0%) | --- | --- | --- |
|  |  | 4-6 months | 1 (1.9%) | 0 (0.0%) | 0 (0.0%) | --- | --- | --- |
|  |  | 7-9 months | 0 (0.0%) | 0 (0.0%) | 1 (1.8%) | --- | --- | --- |
|  | Child any parasitemia | 1-3 months | 8 (15.4%) | 9 (15.0%) | 13 (22.8%) | 0.75 (0.48, 1.16) | --- | 0.82 (0.55, 1.21) |
|  |  | 4-6 months | 21 (40.4%) | 20 (33.3%) | 26 (45.6%) | 0.93 (0.75, 1.16) | *1.47 (1.16, 1.87) | 0.84 (0.67, 1.06) |
|  |  | 7-9 months | 18 (34.6%) | 22 (36.7%) | 26 (45.6%) | *0.59 (0.48, 0.74) | 0.83 (0.65, 1.06) | *0.74 (0.58, 0.94) |
|  | Child anemia | 2 months | 18 (62.1%) | 27 (73.0%) | 22 (55.0%) | 1.10 (0.89, 1.37) | *0.79 (0.67, 0.94) | 1.04 (0.86, 1.26) |
|  |  | 6 months | 20 (69.0%) | 30 (81.1%) | 26 (65.0%) | 0.99 (0.84, 1.18) | *0.83 (0.72, 0.95) | *1.17 (1.02, 1.36) |
|  |  | 13 months | 22 (75.9%) | 30 (81.1%) | 30 (75.0%) | 0.93 (0.83, 1.05) | *0.81 (0.72, 0.92) | 1.05 (0.93, 1.18) |
|  | Child severe anemia | 2 months | 0 (0.0%) | 2 (5.4%) | 0 (0.0%) | --- | --- | --- |
|  |  | 6 months | 3 (10.3%) | 5 (13.5%) | 2 (5.0%) | --- | --- | --- |
|  |  | 13 months | 3 (10.3%) | 4 (10.8%) | 1 (2.5%) | --- | --- | --- |
|  | Child antimalarial use | 1-3 months | 4 (8.0%) | 3 (5.4%) | 5 (8.9%) | --- | --- | --- |
|  |  | 4-6 months | 14 (28.0%) | 8 (14.3%) | 16 (28.6%) | 0.87 (0.62, 1.22) | *3.89 (2.48, 6.09) | *0.44 (0.29, 0.66) |
|  |  | 7-9 months | 12 (24.0%) | 11 (19.6%) | 14 (25.0%) | 0.74 (0.53, 1.02) | 1.00 (0.69, 1.44) | 1.12 (0.80, 1.57) |
|  | Child antiparasitic use | 1-3 months | 0 (0.0%) | 0 (0.0%) | 0 (0.0%) | --- | --- | --- |
|  |  | 4-6 months | 0 (0.0%) | 0 (0.0%) | 0 (0.0%) | --- | --- | --- |
|  |  | 7-9 months | 0 (0.0%) | 0 (0.0%) | 0 (0.0%) | --- | --- | --- |
|  | Child antiviral use | 1-3 months | 0 (0.0%) | 0 (0.0%) | 0 (0.0%) | --- | --- | --- |
|  |  | 4-6 months | 0 (0.0%) | 0 (0.0%) | 0 (0.0%) | --- | --- | --- |
|  |  | 7-9 months | 0 (0.0%) | 0 (0.0%) | 0 (0.0%) | --- | --- | --- |
|  | Child antibacterial use | 1-3 months | 30 (60.0%) | 25 (44.6%) | 29 (51.8%) | *1.23 (1.04, 1.46) | *1.33 (1.12, 1.58) | *0.68 (0.55, 0.84) |
|  |  | 4-6 months | 37 (74.0%) | 38 (67.9%) | 40 (71.4%) | 1.00 (0.88, 1.12) | *1.33 (1.18, 1.50) | 0.92 (0.81, 1.05) |
|  |  | 7-9 months | 33 (66.0%) | 41 (73.2%) | 41 (73.2%) | *0.85 (0.74, 0.97) | 0.93 (0.83, 1.05) | 1.02 (0.93, 1.13) |
|  | Child beta-lactam use | 1-3 months | 28 (56.0%) | 24 (42.9%) | 27 (48.2%) | 1.20 (1.00, 1.44) | *1.29 (1.07, 1.54) | *0.68 (0.55, 0.85) |
|  |  | 4-6 months | 37 (74.0%) | 38 (67.9%) | 37 (66.1%) | 1.03 (0.91, 1.17) | *1.33 (1.18, 1.50) | 0.99 (0.87, 1.13) |
|  |  | 7-9 months | 31 (62.0%) | 40 (71.4%) | 37 (66.1%) | 0.93 (0.80, 1.07) | 0.92 (0.81, 1.04) | *1.13 (1.02, 1.26) |
|  | Child fluoroquinolone use | 1-3 months | 0 (0.0%) | 0 (0.0%) | 0 (0.0%) | --- | --- | --- |
|  |  | 4-6 months | 0 (0.0%) | 0 (0.0%) | 0 (0.0%) | --- | --- | --- |
|  |  | 7-9 months | 0 (0.0%) | 1 (1.8%) | 0 (0.0%) | --- | --- | --- |
|  | Child sulfonamide use | 1-3 months | 0 (0.0%) | 0 (0.0%) | 0 (0.0%) | --- | --- | --- |
|  |  | 4-6 months | 0 (0.0%) | 0 (0.0%) | 0 (0.0%) | --- | --- | --- |
|  |  | 7-9 months | 0 (0.0%) | 0 (0.0%) | 0 (0.0%) | --- | --- | --- |
|  | Child other antibacterial use | 1-3 months | 0 (0.0%) | 1 (1.8%) | 1 (1.8%) | --- | --- | --- |
|  |  | 4-6 months | 0 (0.0%) | 1 (1.8%) | 0 (0.0%) | --- | --- | --- |
|  |  | 7-9 months | 0 (0.0%) | 1 (1.8%) | 3 (5.4%) | --- | --- | --- |
| Multigravida | Any uncomplicated malaria during pregnancy | | 6 (3.2%) | 5 (2.4%) | 24 (12.1%) | *0.27 (0.18, 0.41) | --- | *0.24 (0.15, 0.37) |
|  | Any complicated malaria during pregnancy | | 0 (0.0%) | 0 (0.0%) | 0 (0.0%) | --- | --- | --- |
|  | Any parasitemia during pregnancy | | 162 (86.2%) | 170 (82.9%) | 176 (88.4%) | *0.96 (0.93, 0.99) | 1.02 (0.98, 1.06) | *0.94 (0.90, 0.97) |
|  | Any placental malaria | | 78 (43.6%) | 63 (31.7%) | 116 (60.4%) | *0.69 (0.64, 0.75) | *1.29 (1.15, 1.45) | *0.53 (0.48, 0.59) |
|  | Past placental malaria | | 77 (43.0%) | 63 (31.7%) | 115 (59.9%) | *0.69 (0.64, 0.76) | *1.28 (1.14, 1.43) | *0.54 (0.49, 0.60) |
|  | Active placental malaria | | 1 (0.6%) | 1 (0.5%) | 13 (6.8%) | --- | --- | --- |
|  | High-grade past placental malaria | | 0 (0.0%) | 0 (0.0%) | 7 (6.1%) | --- | --- | --- |
|  | Any anemia during pregnancy | | 112 (59.6%) | 98 (47.8%) | 118 (59.3%) | 1.01 (0.94, 1.09) | *1.24 (1.14, 1.36) | *0.82 (0.76, 0.89) |
|  | Any severe anemia during pregnancy | | 3 (1.6%) | 1 (0.5%) | 3 (1.5%) | --- | --- | --- |
|  | Azithromycin use during pregnancy | | 1 (0.5%) | 2 (1.0%) | 0 (0.0%) | --- | --- | --- |
|  | Tetracycline use during pregnancy | | 0 (0.0%) | 0 (0.0%) | 3 (1.5%) | --- | --- | --- |
|  | Doxycycline use during pregnancy | | 0 (0.0%) | 0 (0.0%) | 1 (0.5%) | --- | --- | --- |
|  | Metronidazole use during pregnancy | | 54 (28.7%) | 68 (33.2%) | 80 (40.2%) | *0.72 (0.64, 0.81) | *0.86 (0.75, 0.98) | *0.84 (0.75, 0.95) |
|  | Ferrous use during pregnancy | | 188 (100.0%) | 205 (100.0%) | 199 (100.0%) | 1.00 (1.00, 1.00) | 1.00 (1.00, 1.00) | 1.00 (1.00, 1.00) |
|  | Below median gestational weight gain per week | | 91 (49.7%) | 104 (51.7%) | 75 (38.7%) | *1.24 (1.12, 1.38) | *0.91 (0.83, 0.99) | *1.30 (1.17, 1.44) |
|  | Preterm birth | | 6 (3.2%) | 9 (4.4%) | 3 (1.5%) | --- | --- | --- |
|  | Low birth weight | | 8 (4.3%) | 12 (5.9%) | 3 (1.5%) | --- | 0.92 (0.58, 1.44) | --- |
|  | Small for gestational age | | 41 (21.8%) | 38 (18.5%) | 36 (18.1%) | *1.52 (1.26, 1.84) | *1.36 (1.13, 1.64) | *1.26 (1.04, 1.51) |
|  | Child uncomplicated malaria | 1-3 months | 22 (11.7%) | 21 (10.2%) | 14 (7.0%) | *1.86 (1.36, 2.54) | 0.99 (0.76, 1.28) | *1.46 (1.07, 1.98) |
|  |  | 4-6 months | 33 (17.6%) | 35 (17.1%) | 31 (15.6%) | *1.34 (1.12, 1.60) | 0.99 (0.82, 1.19) | 1.19 (0.98, 1.44) |
|  |  | 7-9 months | 29 (15.4%) | 52 (25.4%) | 35 (17.6%) | 1.02 (0.84, 1.24) | *0.61 (0.50, 0.73) | *1.66 (1.42, 1.95) |
|  | Child complicated malaria | 1-3 months | 0 (0.0%) | 0 (0.0%) | 0 (0.0%) | --- | --- | --- |
|  |  | 4-6 months | 0 (0.0%) | 1 (0.5%) | 1 (0.5%) | --- | --- | --- |
|  |  | 7-9 months | 1 (0.5%) | 3 (1.5%) | 0 (0.0%) | --- | --- | --- |
|  | Child any parasitemia | 1-3 months | 41 (21.8%) | 46 (22.4%) | 33 (16.6%) | *1.37 (1.14, 1.65) | 0.93 (0.78, 1.10) | *1.38 (1.15, 1.65) |
|  |  | 4-6 months | 73 (38.8%) | 73 (35.6%) | 69 (34.7%) | *1.24 (1.11, 1.39) | 1.11 (0.99, 1.23) | 1.06 (0.94, 1.19) |
|  |  | 7-9 months | 63 (33.5%) | 90 (43.9%) | 70 (35.2%) | 1.09 (0.97, 1.23) | *0.84 (0.75, 0.94) | *1.32 (1.19, 1.47) |
|  | Child anemia | 2 months | 65 (59.6%) | 78 (58.6%) | 82 (63.6%) | 0.96 (0.88, 1.05) | 1.01 (0.92, 1.12) | 0.92 (0.84, 1.00) |
|  |  | 6 months | 78 (71.6%) | 86 (64.7%) | 85 (65.9%) | *1.17 (1.09, 1.27) | *1.15 (1.06, 1.24) | 0.97 (0.89, 1.05) |
|  |  | 13 months | 85 (78.0%) | 96 (72.2%) | 90 (69.8%) | *1.19 (1.11, 1.27) | *1.10 (1.03, 1.18) | 1.03 (0.96, 1.11) |
|  | Child severe anemia | 2 months | 0 (0.0%) | 4 (3.0%) | 1 (0.8%) | --- | --- | --- |
|  |  | 6 months | 6 (5.5%) | 5 (3.8%) | 3 (2.3%) | --- | --- | --- |
|  |  | 13 months | 0 (0.0%) | 8 (6.0%) | 4 (3.1%) | --- | --- | --- |
|  | Child antimalarial use | 1-3 months | 21 (11.3%) | 21 (10.4%) | 13 (6.6%) | *1.81 (1.32, 2.48) | 0.96 (0.74, 1.25) | *1.56 (1.14, 2.14) |
|  |  | 4-6 months | 35 (18.8%) | 37 (18.4%) | 32 (16.2%) | *1.34 (1.13, 1.60) | 1.01 (0.85, 1.21) | 1.20 (0.99, 1.45) |
|  |  | 7-9 months | 30 (16.1%) | 54 (26.9%) | 37 (18.8%) | 0.98 (0.81, 1.19) | *0.60 (0.50, 0.72) | *1.61 (1.37, 1.88) |
|  | Child antiparasitic use | 1-3 months | 0 (0.0%) | 0 (0.0%) | 0 (0.0%) | --- | --- | --- |
|  |  | 4-6 months | 2 (1.1%) | 0 (0.0%) | 1 (0.5%) | --- | --- | --- |
|  |  | 7-9 months | 0 (0.0%) | 1 (0.5%) | 1 (0.5%) | --- | --- | --- |
|  | Child antiviral use | 1-3 months | 0 (0.0%) | 1 (0.5%) | 0 (0.0%) | --- | --- | --- |
|  |  | 4-6 months | 0 (0.0%) | 0 (0.0%) | 2 (1.0%) | --- | --- | --- |
|  |  | 7-9 months | 0 (0.0%) | 1 (0.5%) | 0 (0.0%) | --- | --- | --- |
|  | Child antibacterial use | 1-3 months | 121 (65.1%) | 123 (61.2%) | 128 (65.0%) | 1.03 (0.97, 1.11) | 1.04 (0.97, 1.12) | 0.96 (0.90, 1.03) |
|  |  | 4-6 months | 148 (79.6%) | 148 (73.6%) | 154 (78.2%) | 1.05 (1.00, 1.09) | *1.09 (1.04, 1.15) | *0.94 (0.89, 0.99) |
|  |  | 7-9 months | 133 (71.5%) | 143 (71.1%) | 132 (67.0%) | *1.08 (1.02, 1.15) | 1.02 (0.96, 1.08) | *1.06 (1.00, 1.13) |
|  | Child beta-lactam use | 1-3 months | 115 (61.8%) | 117 (58.2%) | 121 (61.4%) | 1.04 (0.96, 1.11) | 1.04 (0.96, 1.12) | 0.96 (0.90, 1.04) |
|  |  | 4-6 months | 137 (73.7%) | 143 (71.1%) | 151 (76.6%) | 0.99 (0.94, 1.04) | 1.05 (0.99, 1.11) | *0.92 (0.87, 0.97) |
|  |  | 7-9 months | 123 (66.1%) | 137 (68.2%) | 125 (63.5%) | 1.06 (0.99, 1.13) | 0.98 (0.92, 1.05) | *1.07 (1.01, 1.14) |
|  | Child fluoroquinolone use | 1-3 months | 0 (0.0%) | 0 (0.0%) | 2 (1.0%) | --- | --- | --- |
|  |  | 4-6 months | 0 (0.0%) | 2 (1.0%) | 1 (0.5%) | --- | --- | --- |
|  |  | 7-9 months | 0 (0.0%) | 2 (1.0%) | 2 (1.0%) | --- | --- | --- |
|  | Child sulfonamide use | 1-3 months | 0 (0.0%) | 0 (0.0%) | 0 (0.0%) | --- | --- | --- |
|  |  | 4-6 months | 0 (0.0%) | 0 (0.0%) | 0 (0.0%) | --- | --- | --- |
|  |  | 7-9 months | 0 (0.0%) | 0 (0.0%) | 0 (0.0%) | --- | --- | --- |
|  | Child other antibacterial use | 1-3 months | 1 (0.5%) | 2 (1.0%) | 2 (1.0%) | --- | --- | --- |
|  |  | 4-6 months | 2 (1.1%) | 6 (3.0%) | 12 (6.1%) | --- | --- | --- |
|  |  | 7-9 months | 1 (0.5%) | 3 (1.5%) | 6 (3.0%) | --- | --- | --- |
| *Stratified by Child Sex* | | | | | | | | |
| Female | Any uncomplicated malaria during pregnancy | | 5 (4.0%) | 3 (2.5%) | 18 (15.3%) | *0.19 (0.13, 0.28) | --- | *0.20 (0.12, 0.34) |
|  | Any complicated malaria during pregnancy | | 0 (0.0%) | 0 (0.0%) | 0 (0.0%) | --- | --- | --- |
|  | Any parasitemia during pregnancy | | 111 (88.8%) | 100 (82.0%) | 105 (89.0%) | 0.99 (0.95, 1.03) | *1.06 (1.01, 1.11) | *0.92 (0.88, 0.96) |
|  | Any placental malaria | | 60 (50.4%) | 49 (40.8%) | 67 (60.4%) | *0.74 (0.67, 0.82) | *1.14 (1.02, 1.27) | *0.64 (0.58, 0.71) |
|  | Past placental malaria | | 60 (50.4%) | 48 (40.0%) | 67 (60.4%) | *0.74 (0.67, 0.82) | *1.17 (1.05, 1.31) | *0.62 (0.56, 0.69) |
|  | Active placental malaria | | 0 (0.0%) | 1 (0.8%) | 9 (8.1%) | --- | --- | --- |
|  | High-grade past placental malaria | | 0 (0.0%) | 1 (2.1%) | 7 (10.4%) | --- | --- | --- |
|  | Any anemia during pregnancy | | 67 (53.6%) | 62 (50.8%) | 74 (62.7%) | *0.85 (0.77, 0.93) | 1.05 (0.94, 1.17) | *0.82 (0.75, 0.91) |
|  | Any severe anemia during pregnancy | | 0 (0.0%) | 0 (0.0%) | 5 (4.2%) | --- | --- | --- |
|  | Azithromycin use during pregnancy | | 0 (0.0%) | 2 (1.6%) | 0 (0.0%) | --- | --- | --- |
|  | Tetracycline use during pregnancy | | 0 (0.0%) | 0 (0.0%) | 1 (0.8%) | --- | --- | --- |
|  | Doxycycline use during pregnancy | | 0 (0.0%) | 0 (0.0%) | 0 (0.0%) | --- | --- | --- |
|  | Metronidazole use during pregnancy | | 46 (36.8%) | 40 (32.8%) | 55 (46.6%) | *0.79 (0.69, 0.90) | 1.14 (0.97, 1.34) | *0.66 (0.57, 0.77) |
|  | Ferrous use during pregnancy | | 125 (100.0%) | 122 (100.0%) | 118 (100.0%) | 1.00 (1.00, 1.00) | 1.00 (1.00, 1.00) | 1.00 (1.00, 1.00) |
|  | Below median gestational weight gain per week | | 53 (44.5%) | 62 (51.2%) | 41 (36.3%) | 1.12 (0.97, 1.29) | *0.83 (0.74, 0.93) | *1.37 (1.19, 1.57) |
|  | Preterm birth | | 7 (5.6%) | 4 (3.3%) | 3 (2.5%) | --- | --- | --- |
|  | Low birth weight | | 9 (7.2%) | 9 (7.4%) | 4 (3.4%) | --- | --- | --- |
|  | Small for gestational age | | 29 (23.2%) | 22 (18.0%) | 20 (16.9%) | *1.65 (1.27, 2.14) | *1.50 (1.20, 1.88) | 1.10 (0.85, 1.41) |
|  | Child uncomplicated malaria | 1-3 months | 11 (8.8%) | 10 (8.2%) | 7 (5.9%) | --- | 1.23 (0.87, 1.72) | --- |
|  |  | 4-6 months | 27 (21.6%) | 18 (14.8%) | 20 (16.9%) | *1.42 (1.14, 1.77) | *1.74 (1.38, 2.20) | 0.91 (0.72, 1.14) |
|  |  | 7-9 months | 21 (16.8%) | 31 (25.4%) | 23 (19.5%) | 1.02 (0.81, 1.28) | *0.64 (0.51, 0.79) | *1.80 (1.47, 2.20) |
|  | Child complicated malaria | 1-3 months | 0 (0.0%) | 0 (0.0%) | 0 (0.0%) | --- | --- | --- |
|  |  | 4-6 months | 0 (0.0%) | 0 (0.0%) | 0 (0.0%) | --- | --- | --- |
|  |  | 7-9 months | 0 (0.0%) | 3 (2.5%) | 0 (0.0%) | --- | --- | --- |
|  | Child any parasitemia | 1-3 months | 24 (19.2%) | 22 (18.0%) | 16 (13.6%) | *1.50 (1.14, 1.99) | 1.06 (0.84, 1.35) | *1.55 (1.18, 2.03) |
|  |  | 4-6 months | 49 (39.2%) | 46 (37.7%) | 43 (36.4%) | *1.23 (1.07, 1.41) | 1.10 (0.96, 1.25) | *1.19 (1.02, 1.38) |
|  |  | 7-9 months | 44 (35.2%) | 54 (44.3%) | 45 (38.1%) | 1.03 (0.90, 1.18) | *0.85 (0.74, 0.97) | *1.37 (1.19, 1.57) |
|  | Child anemia | 2 months | 42 (60.0%) | 42 (54.5%) | 46 (60.5%) | 0.92 (0.82, 1.03) | 1.08 (0.95, 1.23) | *0.77 (0.68, 0.88) |
|  |  | 6 months | 48 (68.6%) | 48 (62.3%) | 45 (59.2%) | *1.17 (1.05, 1.31) | 1.10 (0.99, 1.23) | 1.12 (0.99, 1.26) |
|  |  | 13 months | 54 (77.1%) | 51 (66.2%) | 48 (63.2%) | *1.23 (1.11, 1.35) | *1.13 (1.02, 1.24) | *1.13 (1.02, 1.26) |
|  | Child severe anemia | 2 months | 0 (0.0%) | 2 (2.6%) | 0 (0.0%) | --- | --- | --- |
|  |  | 6 months | 5 (7.1%) | 3 (3.9%) | 5 (6.6%) | --- | --- | --- |
|  |  | 13 months | 1 (1.4%) | 5 (6.5%) | 1 (1.3%) | --- | --- | --- |
|  | Child antimalarial use | 1-3 months | 10 (8.1%) | 9 (7.6%) | 6 (5.2%) | --- | --- | --- |
|  |  | 4-6 months | 28 (22.8%) | 17 (14.4%) | 23 (20.0%) | *1.28 (1.04, 1.57) | *1.94 (1.52, 2.48) | *0.69 (0.55, 0.87) |
|  |  | 7-9 months | 21 (17.1%) | 33 (28.0%) | 26 (22.6%) | 0.91 (0.73, 1.13) | *0.61 (0.49, 0.76) | *1.61 (1.33, 1.95) |
|  | Child antiparasitic use | 1-3 months | 0 (0.0%) | 0 (0.0%) | 0 (0.0%) | --- | --- | --- |
|  |  | 4-6 months | 2 (1.6%) | 0 (0.0%) | 0 (0.0%) | --- | --- | --- |
|  |  | 7-9 months | 0 (0.0%) | 1 (0.8%) | 1 (0.9%) | --- | --- | --- |
|  | Child antiviral use | 1-3 months | 0 (0.0%) | 1 (0.8%) | 0 (0.0%) | --- | --- | --- |
|  |  | 4-6 months | 0 (0.0%) | 0 (0.0%) | 1 (0.9%) | --- | --- | --- |
|  |  | 7-9 months | 0 (0.0%) | 1 (0.8%) | 0 (0.0%) | --- | --- | --- |
|  | Child antibacterial use | 1-3 months | 79 (64.2%) | 72 (61.0%) | 74 (64.3%) | 1.05 (0.96, 1.14) | 1.08 (0.98, 1.18) | 0.99 (0.90, 1.08) |
|  |  | 4-6 months | 97 (78.9%) | 80 (67.8%) | 88 (76.5%) | 1.04 (0.98, 1.11) | *1.21 (1.13, 1.30) | *0.83 (0.77, 0.89) |
|  |  | 7-9 months | 85 (69.1%) | 87 (73.7%) | 76 (66.1%) | 1.04 (0.96, 1.13) | 0.97 (0.90, 1.04) | *1.10 (1.02, 1.19) |
|  | Child beta-lactam use | 1-3 months | 74 (60.2%) | 69 (58.5%) | 68 (59.1%) | 1.07 (0.97, 1.17) | 1.05 (0.95, 1.16) | 1.03 (0.93, 1.14) |
|  |  | 4-6 months | 92 (74.8%) | 77 (65.3%) | 85 (73.9%) | 1.04 (0.97, 1.11) | *1.19 (1.10, 1.29) | *0.82 (0.76, 0.89) |
|  |  | 7-9 months | 79 (64.2%) | 86 (72.9%) | 71 (61.7%) | 1.04 (0.95, 1.13) | *0.91 (0.85, 0.99) | *1.17 (1.08, 1.28) |
|  | Child fluoroquinolone use | 1-3 months | 0 (0.0%) | 0 (0.0%) | 0 (0.0%) | --- | --- | --- |
|  |  | 4-6 months | 0 (0.0%) | 1 (0.8%) | 1 (0.9%) | --- | --- | --- |
|  |  | 7-9 months | 0 (0.0%) | 3 (2.5%) | 1 (0.9%) | --- | --- | --- |
|  | Child sulfonamide use | 1-3 months | 0 (0.0%) | 0 (0.0%) | 0 (0.0%) | --- | --- | --- |
|  |  | 4-6 months | 0 (0.0%) | 0 (0.0%) | 0 (0.0%) | --- | --- | --- |
|  |  | 7-9 months | 0 (0.0%) | 0 (0.0%) | 0 (0.0%) | --- | --- | --- |
|  | Child other antibacterial use | 1-3 months | 0 (0.0%) | 2 (1.7%) | 2 (1.7%) | --- | --- | --- |
|  |  | 4-6 months | 1 (0.8%) | 1 (0.8%) | 2 (1.7%) | --- | --- | --- |
|  |  | 7-9 months | 0 (0.0%) | 0 (0.0%) | 3 (2.6%) | --- | --- | --- |
| Male | Any uncomplicated malaria during pregnancy | | 5 (4.3%) | 6 (4.2%) | 34 (24.6%) | *0.17 (0.11, 0.26) | --- | *0.15 (0.10, 0.21) |
|  | Any complicated malaria during pregnancy | | 0 (0.0%) | 0 (0.0%) | 0 (0.0%) | --- | --- | --- |
|  | Any parasitemia during pregnancy | | 97 (84.3%) | 121 (84.6%) | 126 (91.3%) | *0.90 (0.86, 0.94) | 0.98 (0.93, 1.03) | *0.93 (0.89, 0.96) |
|  | Any placental malaria | | 57 (51.8%) | 62 (44.6%) | 96 (72.7%) | *0.72 (0.66, 0.78) | *1.19 (1.07, 1.33) | *0.63 (0.57, 0.69) |
|  | Past placental malaria | | 56 (50.9%) | 62 (44.6%) | 95 (72.0%) | *0.72 (0.66, 0.78) | *1.18 (1.06, 1.32) | *0.64 (0.58, 0.70) |
|  | Active placental malaria | | 3 (2.7%) | 2 (1.4%) | 15 (11.4%) | --- | --- | --- |
|  | High-grade past placental malaria | | 0 (0.0%) | 0 (0.0%) | 17 (17.9%) | --- | --- | --- |
|  | Any anemia during pregnancy | | 72 (62.6%) | 67 (46.9%) | 94 (68.1%) | 0.93 (0.86, 1.00) | *1.30 (1.17, 1.44) | *0.71 (0.65, 0.78) |
|  | Any severe anemia during pregnancy | | 4 (3.5%) | 1 (0.7%) | 2 (1.4%) | --- | --- | --- |
|  | Azithromycin use during pregnancy | | 2 (1.7%) | 2 (1.4%) | 0 (0.0%) | --- | --- | --- |
|  | Tetracycline use during pregnancy | | 1 (0.9%) | 0 (0.0%) | 2 (1.4%) | --- | --- | --- |
|  | Doxycycline use during pregnancy | | 0 (0.0%) | 0 (0.0%) | 1 (0.7%) | --- | --- | --- |
|  | Metronidazole use during pregnancy | | 32 (27.8%) | 51 (35.7%) | 54 (39.1%) | *0.72 (0.62, 0.85) | *0.80 (0.67, 0.95) | 0.96 (0.83, 1.11) |
|  | Ferrous use during pregnancy | | 115 (100.0%) | 143 (100.0%) | 138 (100.0%) | 1.00 (1.00, 1.00) | 1.00 (1.00, 1.00) | 1.00 (1.00, 1.00) |
|  | Below median gestational weight gain per week | | 65 (57.5%) | 76 (54.3%) | 59 (43.1%) | *1.25 (1.12, 1.40) | 0.99 (0.90, 1.09) | *1.22 (1.09, 1.37) |
|  | Preterm birth | | 2 (1.7%) | 7 (4.9%) | 4 (2.9%) | --- | --- | --- |
|  | Low birth weight | | 6 (5.2%) | 8 (5.6%) | 5 (3.6%) | --- | --- | --- |
|  | Small for gestational age | | 29 (25.2%) | 34 (23.8%) | 32 (23.2%) | *1.22 (1.00, 1.49) | 1.17 (0.95, 1.44) | 1.13 (0.93, 1.38) |
|  | Child uncomplicated malaria | 1-3 months | 16 (13.9%) | 14 (9.8%) | 11 (8.0%) | *1.51 (1.10, 2.06) | 1.04 (0.76, 1.44) | 1.31 (0.91, 1.90) |
|  |  | 4-6 months | 20 (17.4%) | 26 (18.2%) | 25 (18.1%) | 0.92 (0.73, 1.16) | 0.97 (0.76, 1.24) | 0.99 (0.80, 1.23) |
|  |  | 7-9 months | 20 (17.4%) | 31 (21.7%) | 24 (17.4%) | 1.11 (0.88, 1.40) | 0.83 (0.65, 1.04) | 1.20 (0.97, 1.47) |
|  | Child complicated malaria | 1-3 months | 0 (0.0%) | 0 (0.0%) | 0 (0.0%) | --- | --- | --- |
|  |  | 4-6 months | 1 (0.9%) | 1 (0.7%) | 1 (0.7%) | --- | --- | --- |
|  |  | 7-9 months | 1 (0.9%) | 0 (0.0%) | 1 (0.7%) | --- | --- | --- |
|  | Child any parasitemia | 1-3 months | 25 (21.7%) | 33 (23.1%) | 30 (21.7%) | 0.91 (0.74, 1.12) | 0.90 (0.73, 1.12) | 1.05 (0.87, 1.28) |
|  |  | 4-6 months | 45 (39.1%) | 47 (32.9%) | 52 (37.7%) | 0.99 (0.87, 1.13) | *1.24 (1.07, 1.43) | *0.87 (0.75, 1.00) |
|  |  | 7-9 months | 37 (32.2%) | 58 (40.6%) | 51 (37.0%) | 0.90 (0.78, 1.04) | 0.89 (0.77, 1.03) | 1.02 (0.89, 1.16) |
|  | Child anemia | 2 months | 41 (60.3%) | 63 (67.7%) | 58 (62.4%) | 1.03 (0.92, 1.15) | *0.88 (0.79, 0.98) | *1.11 (1.01, 1.23) |
|  |  | 6 months | 50 (73.5%) | 68 (73.1%) | 66 (71.0%) | *1.09 (1.00, 1.19) | 1.05 (0.97, 1.14) | 1.01 (0.93, 1.10) |
|  |  | 13 months | 53 (77.9%) | 75 (80.6%) | 72 (77.4%) | 1.06 (0.99, 1.15) | 1.00 (0.93, 1.07) | 1.02 (0.96, 1.09) |
|  | Child severe anemia | 2 months | 0 (0.0%) | 4 (4.3%) | 1 (1.1%) | --- | --- | --- |
|  |  | 6 months | 4 (5.9%) | 7 (7.5%) | 0 (0.0%) | --- | --- | --- |
|  |  | 13 months | 2 (2.9%) | 7 (7.5%) | 4 (4.3%) | --- | --- | --- |
|  | Child antimalarial use | 1-3 months | 15 (13.3%) | 15 (10.8%) | 12 (8.7%) | 1.25 (0.93, 1.69) | 0.91 (0.65, 1.25) | 1.34 (0.94, 1.92) |
|  |  | 4-6 months | 21 (18.6%) | 28 (20.1%) | 25 (18.1%) | 0.98 (0.78, 1.23) | 0.95 (0.75, 1.20) | 1.10 (0.89, 1.36) |
|  |  | 7-9 months | 21 (18.6%) | 32 (23.0%) | 25 (18.1%) | 1.17 (0.93, 1.47) | 0.83 (0.66, 1.04) | 1.22 (1.00, 1.50) |
|  | Child antiparasitic use | 1-3 months | 0 (0.0%) | 0 (0.0%) | 0 (0.0%) | --- | --- | --- |
|  |  | 4-6 months | 0 (0.0%) | 0 (0.0%) | 1 (0.7%) | --- | --- | --- |
|  |  | 7-9 months | 0 (0.0%) | 0 (0.0%) | 0 (0.0%) | --- | --- | --- |
|  | Child antiviral use | 1-3 months | 0 (0.0%) | 0 (0.0%) | 0 (0.0%) | --- | --- | --- |
|  |  | 4-6 months | 0 (0.0%) | 0 (0.0%) | 1 (0.7%) | --- | --- | --- |
|  |  | 7-9 months | 0 (0.0%) | 0 (0.0%) | 0 (0.0%) | --- | --- | --- |
|  | Child antibacterial use | 1-3 months | 72 (63.7%) | 76 (54.7%) | 83 (60.1%) | 1.06 (0.97, 1.15) | *1.13 (1.03, 1.24) | 0.92 (0.83, 1.01) |
|  |  | 4-6 months | 88 (77.9%) | 106 (76.3%) | 106 (76.8%) | 1.02 (0.96, 1.09) | 1.04 (0.97, 1.11) | 1.01 (0.95, 1.07) |
|  |  | 7-9 months | 81 (71.7%) | 97 (69.8%) | 97 (70.3%) | 1.02 (0.95, 1.10) | 1.02 (0.95, 1.10) | 1.00 (0.93, 1.07) |
|  | Child beta-lactam use | 1-3 months | 69 (61.1%) | 72 (51.8%) | 80 (58.0%) | 1.05 (0.95, 1.15) | *1.13 (1.02, 1.24) | *0.90 (0.82, 0.99) |
|  |  | 4-6 months | 82 (72.6%) | 104 (74.8%) | 103 (74.6%) | 0.98 (0.92, 1.05) | 1.00 (0.93, 1.07) | 1.01 (0.95, 1.08) |
|  |  | 7-9 months | 75 (66.4%) | 91 (65.5%) | 91 (65.9%) | 1.01 (0.94, 1.10) | 1.02 (0.94, 1.11) | 0.99 (0.92, 1.07) |
|  | Child fluoroquinolone use | 1-3 months | 0 (0.0%) | 0 (0.0%) | 2 (1.4%) | --- | --- | --- |
|  |  | 4-6 months | 0 (0.0%) | 1 (0.7%) | 0 (0.0%) | --- | --- | --- |
|  |  | 7-9 months | 0 (0.0%) | 0 (0.0%) | 1 (0.7%) | --- | --- | --- |
|  | Child sulfonamide use | 1-3 months | 0 (0.0%) | 0 (0.0%) | 0 (0.0%) | --- | --- | --- |
|  |  | 4-6 months | 0 (0.0%) | 0 (0.0%) | 0 (0.0%) | --- | --- | --- |
|  |  | 7-9 months | 0 (0.0%) | 0 (0.0%) | 0 (0.0%) | --- | --- | --- |
|  | Child other antibacterial use | 1-3 months | 1 (0.9%) | 1 (0.7%) | 1 (0.7%) | --- | --- | --- |
|  |  | 4-6 months | 1 (0.9%) | 6 (4.3%) | 10 (7.2%) | --- | --- | --- |
|  |  | 7-9 months | 1 (0.9%) | 4 (2.9%) | 6 (4.3%) | --- | --- | --- |

Asterisks indicate effect estimates in which the confidence interval does not contain the null. Effects were not estimated if there were less than 20 observed cases, as indicated with “---".

### Figure S6: Treatment-Mediator Effects, Stratified by Child Sex

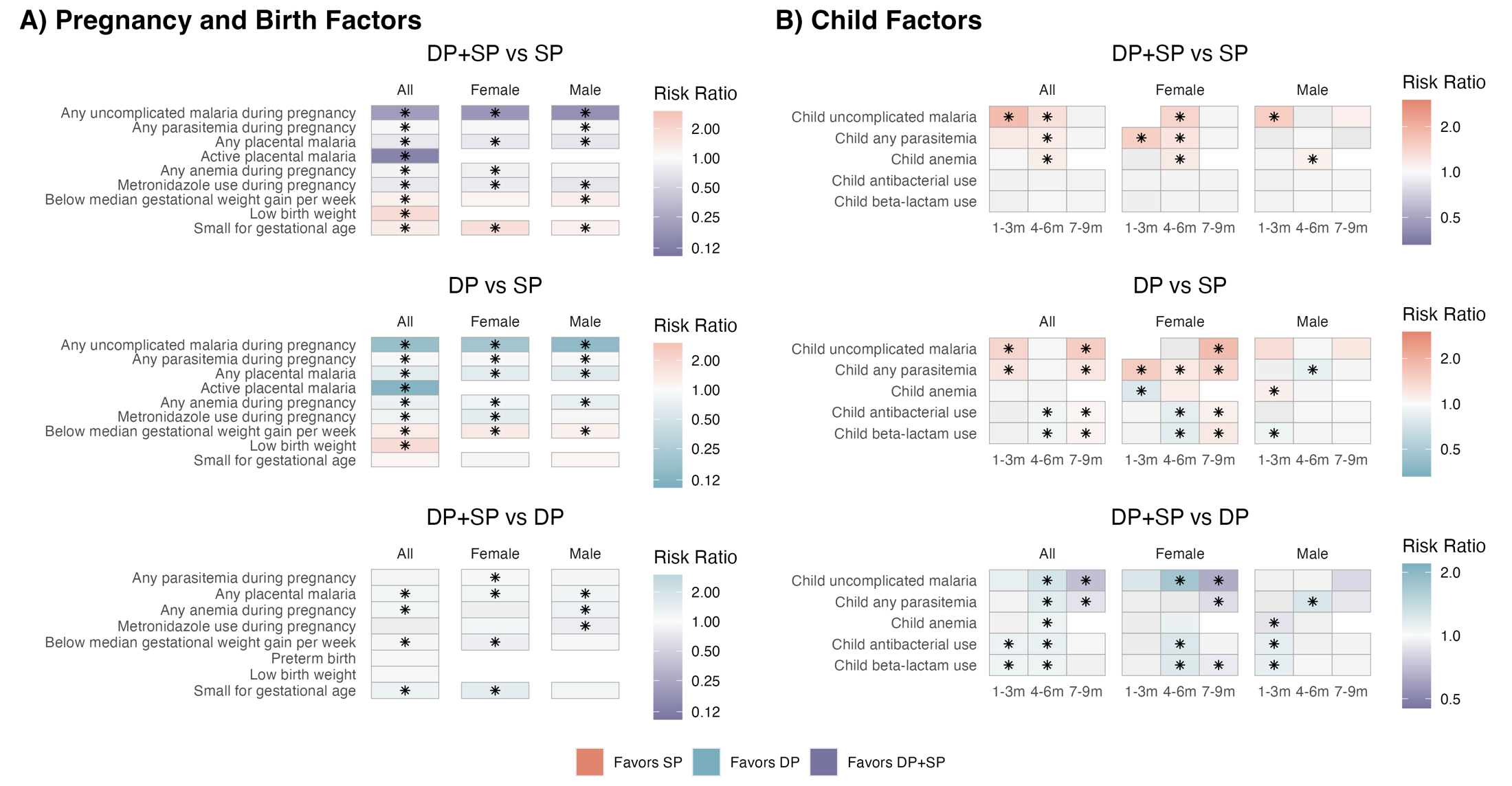

Heatmap of estimated prevalence ratios for IPTp regimens on (A) pregnancy and birth factors and (b) infant factors measured at different ages. Prevalence ratios were estimated with log-binomial models with robust standard errors, adjusting for baseline covariates. If log-binomial models failed to converge, we used modified Poisson regression models. Tiles are shaded in color if the prevalence ratio is large (PR > 1.1, PR < 0.9) or or had p-values < 0.05. Estimates where confidence intervals did not cross the null are marked by asterisks. Color is determined by the comparison arm with the lower risk of the mediator (the arm “favored” to provide protection against the mediator). Effects were not estimated if there were less than 20 observed cases, and are indicated by the absence of a tile.

### Figure S7: Mediator-Outcome Effects, Stratified by Gravidity

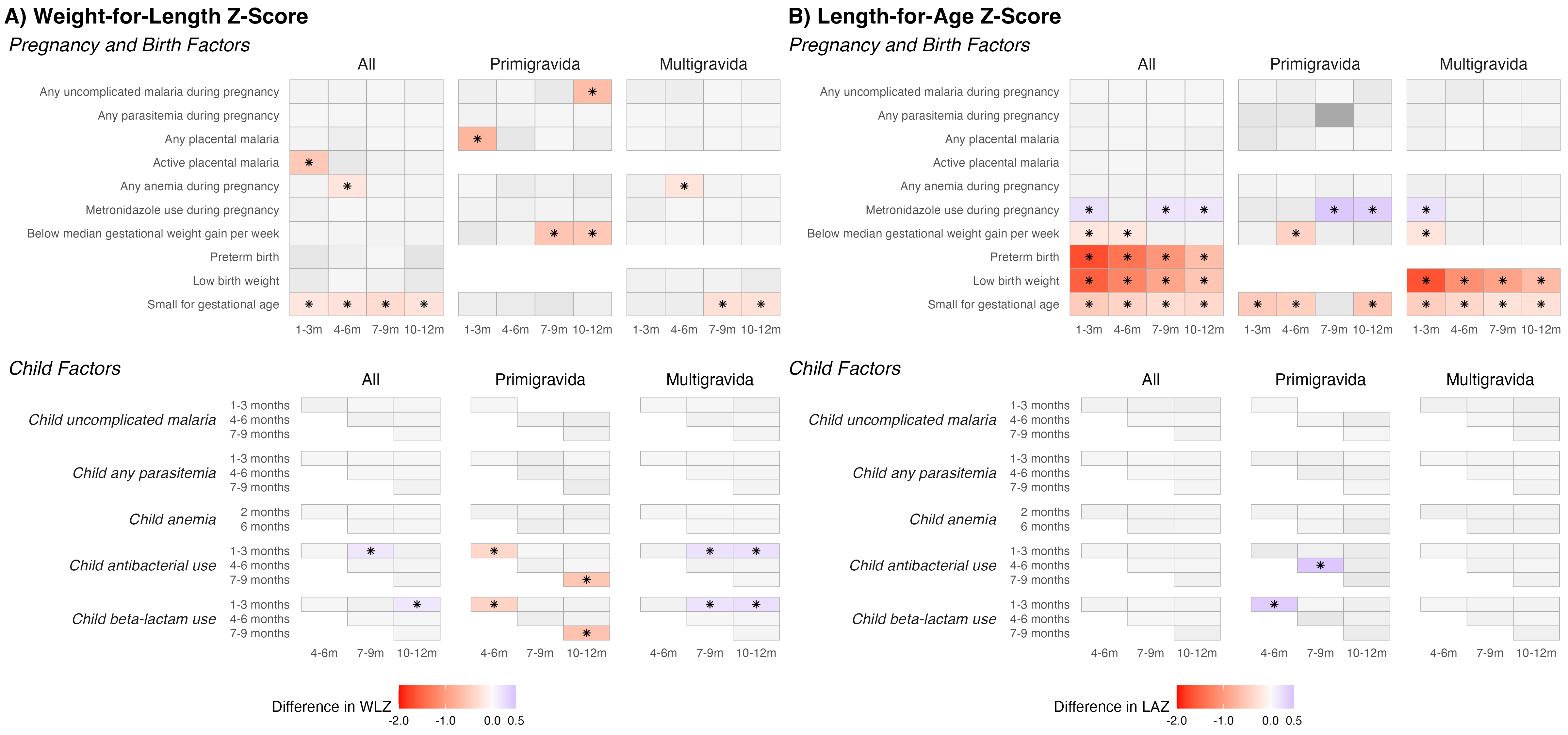

Heatmap of estimated mean differences by mediators on (A) weight-for-length z-scores (WLZs) and (b) length-for-age z-scores (LAZs). Mean differences were estimated with generalized linear models with a Gaussian family for continuous outcomes, adjusting for baseline covariates. Tiles are shaded in color if the prevalence ratio is large (absolute mean difference > 0.5) or had p-values < 0.05. Estimates where confidence intervals did not cross the null are marked by asterisks. Effects were not estimated if there were less than 20 observed cases, or if the mediator did not precede the outcome – these cases are indicated by the absence of a tile.

### Table S7: Effects of Mediators on WLZs

|  | **Mediator** | **Mediator Age** | **Mean difference in WLZ (95% CI)** | | | |
| --- | --- | --- | --- | --- | --- | --- |
|  |  |  | 1-3 months | 4-6 months | 7-9 months | 10-12 months |
| Overall | Any uncomplicated malaria during pregnancy | | -0.09 (-0.40, 0.21) | -0.09 (-0.40, 0.22) | -0.05 (-0.34, 0.25) | -0.11 (-0.41, 0.18) |
|  | Any parasitemia during pregnancy | | 0.00 (-0.25, 0.25) | -0.05 (-0.30, 0.21) | -0.02 (-0.25, 0.22) | -0.01 (-0.25, 0.23) |
|  | Any placental malaria | | -0.07 (-0.26, 0.12) | -0.18 (-0.38, 0.01) | -0.02 (-0.21, 0.16) | 0.01 (-0.17, 0.20) |
|  | Past placental malaria | | -0.07 (-0.26, 0.12) | -0.17 (-0.36, 0.03) | -0.02 (-0.20, 0.16) | 0.00 (-0.18, 0.19) |
|  | Active placental malaria | | *-0.53 (-0.96, -0.09) | -0.32 (-0.77, 0.12) | -0.15 (-0.56, 0.27) | 0.09 (-0.33, 0.52) |
|  | High-grade past placental malaria | | -0.39 (-0.92, 0.14) | -0.10 (-0.63, 0.44) | -0.12 (-0.65, 0.41) | 0.04 (-0.50, 0.58) |
|  | Any anemia during pregnancy | | -0.09 (-0.27, 0.08) | *-0.23 (-0.40, -0.05) | -0.07 (-0.23, 0.09) | -0.06 (-0.22, 0.11) |
|  | Metronidazole use during pregnancy | | -0.14 (-0.31, 0.04) | 0.00 (-0.17, 0.18) | 0.05 (-0.12, 0.21) | 0.08 (-0.09, 0.24) |
|  | Below median gestational weight gain per week | | -0.05 (-0.22, 0.12) | -0.06 (-0.23, 0.12) | -0.10 (-0.27, 0.06) | -0.09 (-0.25, 0.08) |
|  | Preterm birth | | -0.39 (-0.84, 0.06) | 0.16 (-0.29, 0.61) | -0.03 (-0.45, 0.40) | -0.42 (-0.85, 0.01) |
|  | Low birth weight | | -0.29 (-0.67, 0.08) | 0.02 (-0.35, 0.40) | -0.12 (-0.47, 0.23) | -0.32 (-0.67, 0.04) |
|  | Small for gestational age | | *-0.21 (-0.42, -0.01) | *-0.26 (-0.47, -0.05) | *-0.33 (-0.53, -0.14) | *-0.26 (-0.46, -0.06) |
|  | Child uncomplicated malaria | 1-3 months | --- | -0.13 (-0.42, 0.17) | -0.01 (-0.29, 0.27) | 0.03 (-0.25, 0.32) |
|  |  | 4-6 months | --- | --- | 0.13 (-0.09, 0.35) | 0.05 (-0.17, 0.27) |
|  |  | 7-9 months | --- | --- | --- | -0.03 (-0.24, 0.18) |
|  | Child any parasitemia | 1-3 months | --- | -0.01 (-0.23, 0.20) | -0.00 (-0.20, 0.20) | 0.02 (-0.18, 0.23) |
|  |  | 4-6 months | --- | --- | 0.00 (-0.17, 0.18) | -0.02 (-0.20, 0.15) |
|  |  | 7-9 months | --- | --- | --- | 0.11 (-0.07, 0.29) |
|  | Child anemia | 2 months | --- | 0.04 (-0.18, 0.26) | -0.00 (-0.21, 0.20) | 0.01 (-0.20, 0.22) |
|  |  | 6 months | --- | --- | 0.06 (-0.16, 0.27) | 0.05 (-0.18, 0.27) |
|  | Child antimalarial use | 1-3 months | --- | -0.20 (-0.50, 0.10) | -0.06 (-0.34, 0.23) | -0.01 (-0.29, 0.28) |
|  |  | 4-6 months | --- | --- | 0.13 (-0.09, 0.34) | 0.07 (-0.15, 0.28) |
|  |  | 7-9 months | --- | --- | --- | 0.01 (-0.20, 0.22) |
|  | Child antibacterial use | 1-3 months | --- | 0.01 (-0.16, 0.19) | *0.17 (0.01, 0.33) | 0.16 (-0.01, 0.32) |
|  |  | 4-6 months | --- | --- | 0.07 (-0.12, 0.25) | 0.06 (-0.13, 0.25) |
|  |  | 7-9 months | --- | --- | --- | -0.08 (-0.26, 0.09) |
|  | Child beta-lactam use | 1-3 months | --- | 0.01 (-0.16, 0.18) | 0.15 (-0.01, 0.32) | *0.17 (0.01, 0.33) |
|  |  | 4-6 months | --- | --- | -0.02 (-0.19, 0.16) | -0.03 (-0.21, 0.15) |
|  |  | 7-9 months | --- | --- | --- | -0.03 (-0.19, 0.14) |
| *Stratified by Gravidity* | | | | | | |
| Primigravida | Any uncomplicated malaria during pregnancy | | -0.18 (-0.72, 0.36) | -0.05 (-0.56, 0.46) | -0.30 (-0.80, 0.20) | *-0.67 (-1.17, -0.17) |
|  | Any parasitemia during pregnancy | | -0.11 (-0.79, 0.56) | -0.07 (-0.71, 0.57) | -0.03 (-0.66, 0.61) | -0.03 (-0.67, 0.62) |
|  | Any placental malaria | | *-0.73 (-1.25, -0.21) | -0.36 (-0.89, 0.16) | -0.02 (-0.55, 0.50) | -0.23 (-0.75, 0.30) |
|  | Past placental malaria | | *-0.77 (-1.28, -0.25) | -0.39 (-0.91, 0.13) | -0.08 (-0.60, 0.44) | -0.26 (-0.79, 0.26) |
|  | Any anemia during pregnancy | | -0.02 (-0.46, 0.42) | -0.24 (-0.65, 0.17) | -0.14 (-0.55, 0.27) | -0.27 (-0.68, 0.15) |
|  | Metronidazole use during pregnancy | | 0.12 (-0.27, 0.50) | 0.15 (-0.22, 0.51) | 0.08 (-0.28, 0.44) | 0.14 (-0.23, 0.50) |
|  | Below median gestational weight gain per week | | -0.33 (-0.75, 0.09) | -0.12 (-0.53, 0.28) | *-0.57 (-0.96, -0.18) | *-0.54 (-0.93, -0.14) |
|  | Small for gestational age | | -0.20 (-0.61, 0.21) | -0.27 (-0.66, 0.11) | -0.38 (-0.75, 0.00) | -0.17 (-0.56, 0.21) |
|  | Child uncomplicated malaria | 4-6 months | --- | --- | 0.12 (-0.33, 0.57) | 0.22 (-0.23, 0.67) |
|  |  | 7-9 months | --- | --- | --- | 0.16 (-0.31, 0.62) |
|  | Child any parasitemia | 1-3 months | --- | -0.03 (-0.50, 0.45) | 0.19 (-0.28, 0.66) | -0.04 (-0.52, 0.43) |
|  |  | 4-6 months | --- | --- | 0.20 (-0.18, 0.57) | 0.13 (-0.25, 0.51) |
|  |  | 7-9 months | --- | --- | --- | 0.25 (-0.17, 0.67) |
|  | Child anemia | 2 months | --- | -0.12 (-0.60, 0.36) | -0.11 (-0.62, 0.39) | -0.18 (-0.69, 0.33) |
|  |  | 6 months | --- | --- | 0.19 (-0.34, 0.72) | -0.09 (-0.62, 0.45) |
|  | Child antimalarial use | 4-6 months | --- | --- | 0.07 (-0.38, 0.53) | 0.24 (-0.22, 0.70) |
|  |  | 7-9 months | --- | --- | --- | 0.13 (-0.33, 0.59) |
|  | Child antibacterial use | 1-3 months | --- | *-0.37 (-0.74, -0.01) | -0.02 (-0.39, 0.36) | -0.10 (-0.47, 0.28) |
|  |  | 4-6 months | --- | --- | -0.12 (-0.54, 0.30) | -0.02 (-0.45, 0.40) |
|  |  | 7-9 months | --- | --- | --- | *-0.56 (-0.98, -0.14) |
|  | Child beta-lactam use | 1-3 months | --- | *-0.42 (-0.79, -0.06) | -0.08 (-0.45, 0.29) | -0.11 (-0.49, 0.26) |
|  |  | 4-6 months | --- | --- | -0.19 (-0.60, 0.22) | -0.10 (-0.52, 0.32) |
|  |  | 7-9 months | --- | --- | --- | *-0.60 (-1.01, -0.18) |
| Multigravida | Any uncomplicated malaria during pregnancy | | -0.17 (-0.58, 0.23) | -0.22 (-0.63, 0.20) | -0.02 (-0.41, 0.37) | 0.07 (-0.32, 0.46) |
|  | Any parasitemia during pregnancy | | 0.01 (-0.25, 0.28) | -0.07 (-0.34, 0.21) | -0.03 (-0.29, 0.22) | -0.00 (-0.26, 0.26) |
|  | Any placental malaria | | 0.02 (-0.19, 0.23) | -0.15 (-0.36, 0.06) | -0.03 (-0.22, 0.17) | 0.06 (-0.14, 0.26) |
|  | Past placental malaria | | 0.03 (-0.18, 0.24) | -0.13 (-0.34, 0.08) | -0.02 (-0.22, 0.18) | 0.05 (-0.15, 0.25) |
|  | Any anemia during pregnancy | | -0.15 (-0.34, 0.04) | *-0.24 (-0.43, -0.04) | -0.08 (-0.26, 0.10) | -0.03 (-0.21, 0.15) |
|  | Metronidazole use during pregnancy | | -0.18 (-0.38, 0.02) | -0.03 (-0.23, 0.18) | 0.04 (-0.15, 0.23) | 0.06 (-0.13, 0.25) |
|  | Below median gestational weight gain per week | | 0.04 (-0.15, 0.24) | 0.02 (-0.18, 0.22) | 0.03 (-0.16, 0.21) | 0.04 (-0.15, 0.22) |
|  | Low birth weight | | -0.15 (-0.63, 0.34) | -0.11 (-0.61, 0.39) | -0.10 (-0.56, 0.36) | -0.27 (-0.74, 0.20) |
|  | Small for gestational age | | -0.18 (-0.43, 0.06) | -0.23 (-0.48, 0.02) | *-0.28 (-0.51, -0.05) | *-0.27 (-0.50, -0.03) |
|  | Child uncomplicated malaria | 1-3 months | --- | -0.03 (-0.36, 0.29) | 0.00 (-0.30, 0.31) | 0.14 (-0.16, 0.45) |
|  |  | 4-6 months | --- | --- | 0.12 (-0.13, 0.38) | 0.00 (-0.25, 0.26) |
|  |  | 7-9 months | --- | --- | --- | -0.07 (-0.31, 0.17) |
|  | Child any parasitemia | 1-3 months | --- | -0.02 (-0.26, 0.22) | -0.04 (-0.27, 0.18) | 0.04 (-0.18, 0.27) |
|  |  | 4-6 months | --- | --- | -0.05 (-0.24, 0.15) | -0.05 (-0.24, 0.15) |
|  |  | 7-9 months | --- | --- | --- | 0.07 (-0.13, 0.28) |
|  | Child anemia | 2 months | --- | 0.03 (-0.21, 0.27) | -0.02 (-0.24, 0.21) | -0.00 (-0.24, 0.23) |
|  |  | 6 months | --- | --- | -0.01 (-0.25, 0.23) | 0.04 (-0.21, 0.28) |
|  | Child antimalarial use | 1-3 months | --- | -0.11 (-0.44, 0.22) | -0.04 (-0.34, 0.27) | 0.08 (-0.23, 0.38) |
|  |  | 4-6 months | --- | --- | 0.11 (-0.14, 0.36) | -0.01 (-0.26, 0.24) |
|  |  | 7-9 months | --- | --- | --- | -0.01 (-0.24, 0.22) |
|  | Child antibacterial use | 1-3 months | --- | 0.12 (-0.08, 0.32) | *0.21 (0.02, 0.39) | *0.22 (0.04, 0.41) |
|  |  | 4-6 months | --- | --- | 0.10 (-0.11, 0.31) | 0.06 (-0.16, 0.27) |
|  |  | 7-9 months | --- | --- | --- | -0.02 (-0.22, 0.17) |
|  | Child beta-lactam use | 1-3 months | --- | 0.12 (-0.08, 0.31) | *0.20 (0.02, 0.38) | *0.24 (0.06, 0.42) |
|  |  | 4-6 months | --- | --- | 0.02 (-0.18, 0.22) | -0.03 (-0.23, 0.17) |
|  |  | 7-9 months | --- | --- | --- | 0.06 (-0.13, 0.24) |
| *Stratified by Gender* | | | | | | |
| Female | Any uncomplicated malaria during pregnancy | | -0.38 (-0.85, 0.09) | -0.23 (-0.69, 0.24) | -0.23 (-0.66, 0.20) | -0.28 (-0.72, 0.17) |
|  | Any parasitemia during pregnancy | | 0.06 (-0.29, 0.40) | -0.05 (-0.39, 0.29) | 0.08 (-0.24, 0.39) | 0.02 (-0.31, 0.34) |
|  | Any placental malaria | | 0.03 (-0.23, 0.30) | 0.07 (-0.19, 0.33) | 0.20 (-0.04, 0.44) | 0.23 (-0.01, 0.48) |
|  | Past placental malaria | | 0.02 (-0.24, 0.28) | 0.06 (-0.20, 0.33) | 0.19 (-0.05, 0.43) | 0.23 (-0.02, 0.48) |
|  | Any anemia during pregnancy | | 0.07 (-0.16, 0.31) | -0.22 (-0.45, 0.00) | -0.08 (-0.29, 0.14) | -0.06 (-0.27, 0.16) |
|  | Metronidazole use during pregnancy | | 0.02 (-0.22, 0.26) | -0.07 (-0.30, 0.17) | -0.05 (-0.27, 0.17) | -0.07 (-0.29, 0.16) |
|  | Below median gestational weight gain per week | | -0.04 (-0.29, 0.20) | 0.05 (-0.19, 0.29) | -0.06 (-0.28, 0.17) | -0.02 (-0.25, 0.21) |
|  | Small for gestational age | | *-0.34 (-0.64, -0.05) | -0.27 (-0.56, 0.02) | *-0.28 (-0.55, -0.01) | *-0.31 (-0.59, -0.03) |
|  | Child uncomplicated malaria | 1-3 months | --- | 0.14 (-0.29, 0.57) | 0.11 (-0.29, 0.51) | 0.16 (-0.25, 0.57) |
|  |  | 4-6 months | --- | --- | 0.21 (-0.09, 0.50) | 0.25 (-0.06, 0.55) |
|  |  | 7-9 months | --- | --- | --- | 0.22 (-0.06, 0.50) |
|  | Child any parasitemia | 1-3 months | --- | 0.04 (-0.26, 0.34) | 0.10 (-0.18, 0.37) | 0.13 (-0.16, 0.41) |
|  |  | 4-6 months | --- | --- | 0.04 (-0.19, 0.27) | 0.08 (-0.16, 0.31) |
|  |  | 7-9 months | --- | --- | --- | 0.23 (-0.01, 0.47) |
|  | Child anemia | 2 months | --- | 0.09 (-0.20, 0.38) | -0.01 (-0.29, 0.27) | 0.09 (-0.20, 0.38) |
|  |  | 6 months | --- | --- | 0.10 (-0.18, 0.39) | 0.16 (-0.13, 0.46) |
|  | Child antimalarial use | 4-6 months | --- | --- | 0.24 (-0.04, 0.52) | 0.28 (-0.01, 0.56) |
|  |  | 7-9 months | --- | --- | --- | *0.29 (0.03, 0.55) |
|  | Child antibacterial use | 1-3 months | --- | 0.15 (-0.08, 0.38) | *0.24 (0.03, 0.46) | *0.26 (0.04, 0.47) |
|  |  | 4-6 months | --- | --- | 0.09 (-0.15, 0.33) | 0.08 (-0.16, 0.32) |
|  |  | 7-9 months | --- | --- | --- | -0.01 (-0.24, 0.22) |
|  | Child beta-lactam use | 1-3 months | --- | 0.11 (-0.12, 0.33) | 0.18 (-0.03, 0.40) | *0.22 (0.01, 0.43) |
|  |  | 4-6 months | --- | --- | 0.06 (-0.18, 0.29) | 0.02 (-0.21, 0.26) |
|  |  | 7-9 months | --- | --- | --- | 0.03 (-0.20, 0.25) |
| Male | Any uncomplicated malaria during pregnancy | | 0.12 (-0.30, 0.54) | 0.02 (-0.41, 0.45) | 0.08 (-0.32, 0.49) | -0.04 (-0.45, 0.37) |
|  | Any parasitemia during pregnancy | | -0.01 (-0.38, 0.36) | -0.05 (-0.43, 0.33) | -0.09 (-0.45, 0.27) | -0.03 (-0.39, 0.34) |
|  | Any placental malaria | | -0.11 (-0.39, 0.17) | *-0.38 (-0.67, -0.08) | -0.19 (-0.47, 0.08) | -0.18 (-0.46, 0.10) |
|  | Past placental malaria | | -0.09 (-0.37, 0.19) | *-0.33 (-0.62, -0.03) | -0.17 (-0.45, 0.11) | -0.18 (-0.46, 0.10) |
|  | Any anemia during pregnancy | | -0.23 (-0.49, 0.02) | -0.20 (-0.47, 0.06) | -0.03 (-0.28, 0.22) | -0.03 (-0.28, 0.22) |
|  | Metronidazole use during pregnancy | | *-0.32 (-0.58, -0.06) | 0.07 (-0.20, 0.34) | 0.17 (-0.08, 0.42) | *0.26 (0.00, 0.51) |
|  | Below median gestational weight gain per week | | -0.04 (-0.29, 0.21) | -0.12 (-0.38, 0.14) | -0.13 (-0.37, 0.12) | -0.12 (-0.36, 0.13) |
|  | Small for gestational age | | -0.13 (-0.42, 0.17) | -0.22 (-0.52, 0.08) | *-0.34 (-0.63, -0.06) | -0.19 (-0.48, 0.09) |
|  | Child uncomplicated malaria | 1-3 months | --- | -0.42 (-0.83, 0.00) | -0.20 (-0.60, 0.19) | -0.13 (-0.53, 0.27) |
|  |  | 4-6 months | --- | --- | 0.08 (-0.25, 0.41) | -0.09 (-0.42, 0.24) |
|  |  | 7-9 months | --- | --- | --- | -0.23 (-0.55, 0.09) |
|  | Child any parasitemia | 1-3 months | --- | -0.11 (-0.41, 0.20) | -0.11 (-0.40, 0.18) | -0.08 (-0.37, 0.21) |
|  |  | 4-6 months | --- | --- | -0.04 (-0.30, 0.22) | -0.12 (-0.38, 0.15) |
|  |  | 7-9 months | --- | --- | --- | -0.00 (-0.28, 0.28) |
|  | Child anemia | 2 months | --- | -0.10 (-0.44, 0.23) | -0.07 (-0.39, 0.24) | -0.10 (-0.41, 0.22) |
|  |  | 6 months | --- | --- | -0.09 (-0.43, 0.25) | -0.14 (-0.48, 0.21) |
|  | Child antimalarial use | 1-3 months | --- | *-0.48 (-0.89, -0.06) | -0.26 (-0.65, 0.14) | -0.16 (-0.55, 0.24) |
|  |  | 4-6 months | --- | --- | 0.04 (-0.28, 0.37) | -0.11 (-0.43, 0.22) |
|  |  | 7-9 months | --- | --- | --- | -0.23 (-0.55, 0.09) |
|  | Child antibacterial use | 1-3 months | --- | -0.12 (-0.38, 0.14) | 0.10 (-0.14, 0.35) | 0.07 (-0.18, 0.32) |
|  |  | 4-6 months | --- | --- | 0.05 (-0.23, 0.33) | 0.05 (-0.23, 0.34) |
|  |  | 7-9 months | --- | --- | --- | -0.16 (-0.42, 0.11) |
|  | Child beta-lactam use | 1-3 months | --- | -0.10 (-0.36, 0.16) | 0.12 (-0.12, 0.37) | 0.12 (-0.13, 0.37) |
|  |  | 4-6 months | --- | --- | -0.09 (-0.37, 0.18) | -0.09 (-0.36, 0.19) |
|  |  | 7-9 months | --- | --- | --- | -0.07 (-0.33, 0.19) |

Asterisks indicate effect estimates in which the confidence interval does not contain the null. Effects were not estimated if there were less than 20 observed cases, or if the mediator preceded the outcome, as indicated with “---”

### Figure S8: Mediator-Outcome Effects, Stratified by Child Sex

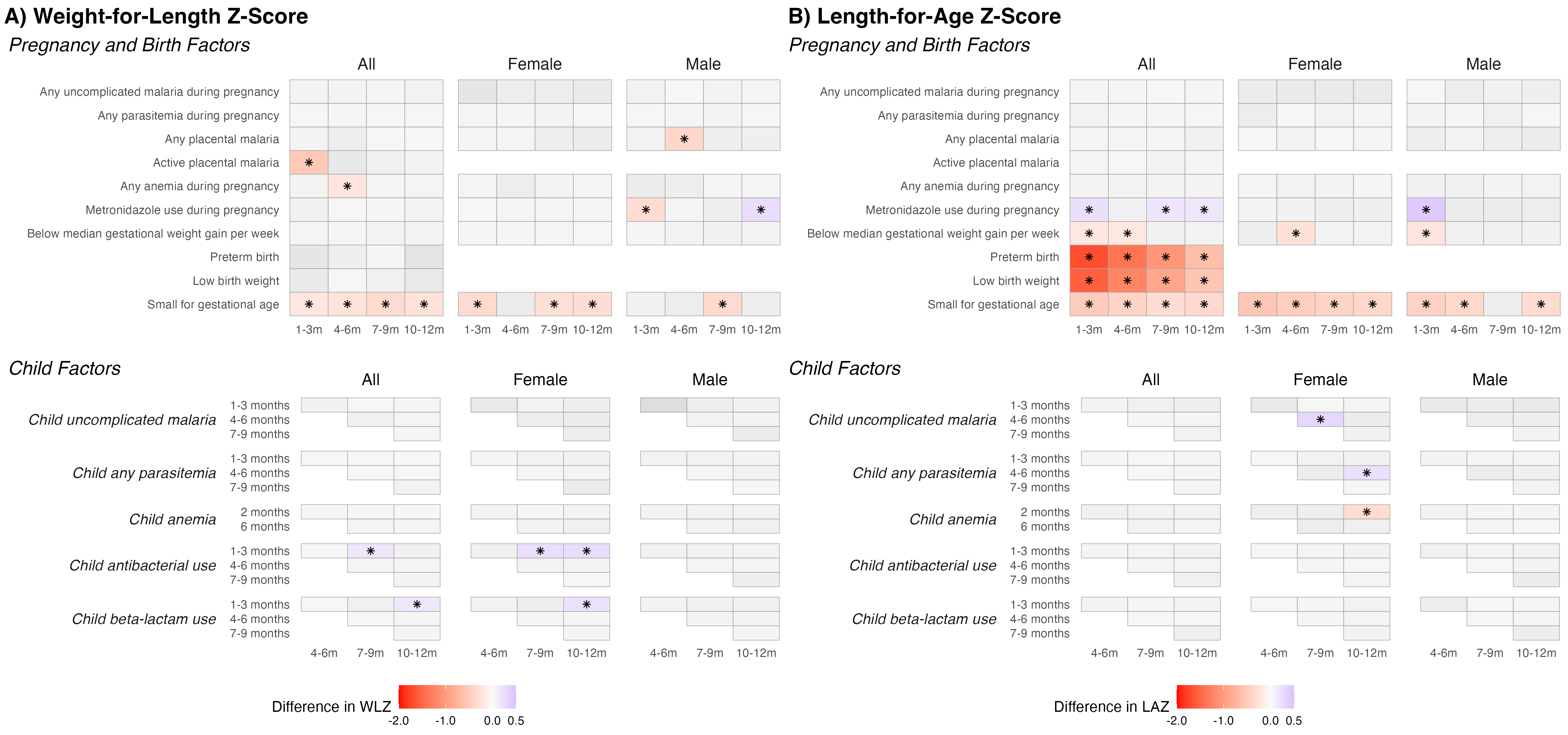

Heatmap of estimated mean differences by mediators on (A) weight-for-length z-scores (WLZs) and (b) length-for-age z-scores (LAZs). Mean differences were estimated with generalized linear models with a Gaussian family for continuous outcomes, adjusting for baseline covariates. Tiles are shaded in color if the effect size was large (absolute mean difference > 0.5) or had p-values < 0.05. Estimates where confidence intervals did not cross the null are marked by asterisks. Effects were not estimated if there were less than 20 observed cases, or if the mediator did not precede the outcome – these cases are indicated by the absence of a tile.

### Figure S9: Mediator-Outcome Effects, Incidence of Wasting

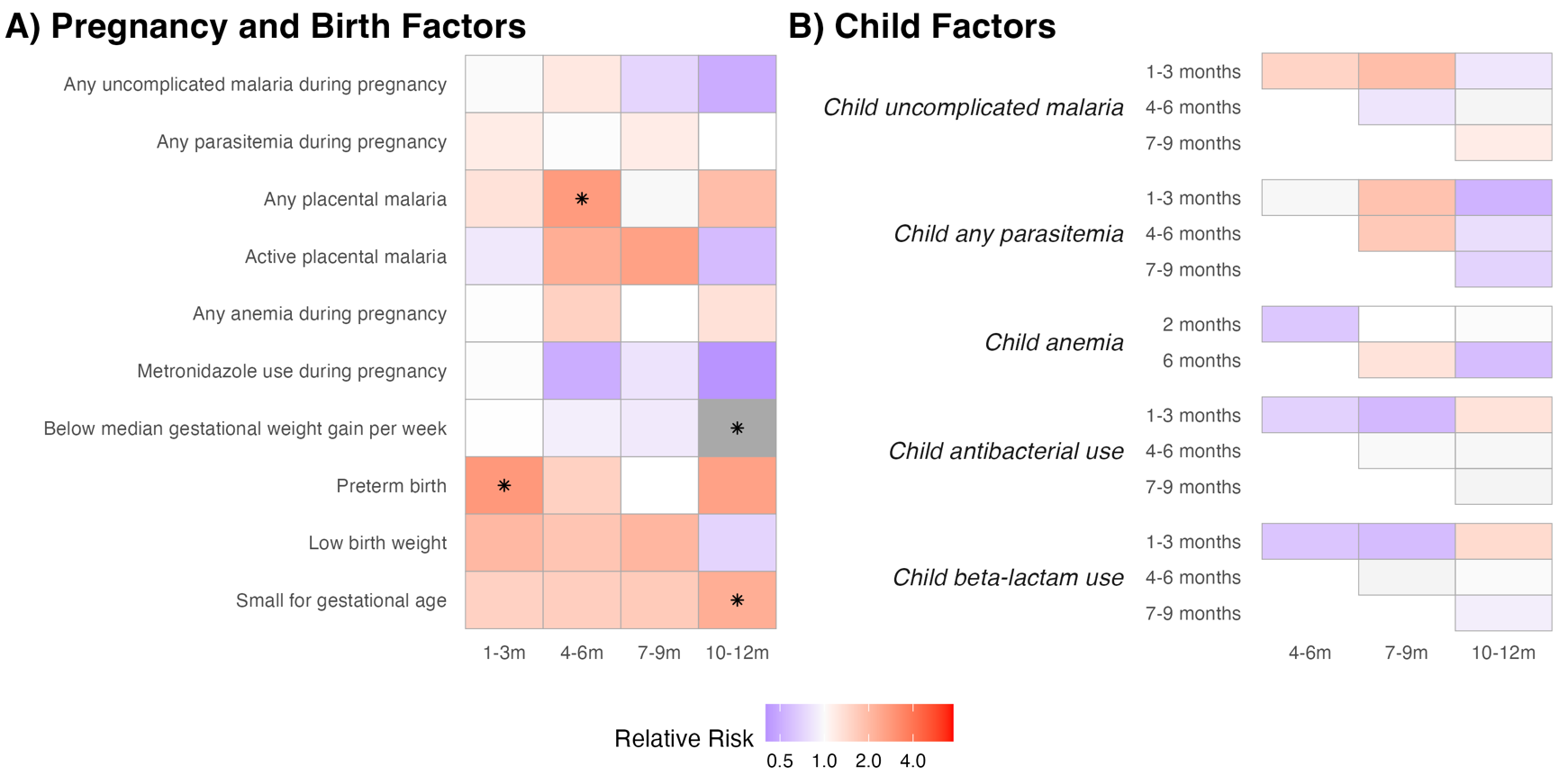

Heatmap of estimated incidence rate ratios for (A) pregnancy and birth factors and (b) infant factors measured at different ages on wasting incidence. Children were considered at risk of incident wasting if they had not been wasted for the prior 60 days. Incidence rate ratios were estimated with log-binomial models with robust standard errors, adjusting for baseline covariates. If log-binomial models failed to converge, we used modified Poisson regression models. Tiles are shaded in color if the incidence rate ratio is large (IRR > 1.1, IRR < 0.9) or had p-values < 0.05. Estimates where confidence intervals did not cross the null are marked by asterisks. Effects were not estimated if there were less than 20 observed cases, or if the mediator did not precede the outcome – these cases are indicated by the absence of a tile.

### Table S8: Effects of Mediators on Incidence of Wasting

| **Mediator** | **Mediator Age** | **Risk Ratio for Wasting Onset (95% CI)** | | | | |
| --- | --- | --- | --- | --- | --- | --- |
|  |  | Birth | 1-3 months | 4-6 months | 7-9 months | 10-12 months |
| Any uncomplicated malaria during pregnancy | | 1.62 (0.67, 3.95) | 1.08 (0.45, 2.59) | 1.22 (0.41, 3.67) | 0.72 (0.22, 2.42) | 0.50 (0.12, 2.03) |
| Any parasitemia during pregnancy | | 0.71 (0.34, 1.48) | 1.17 (0.52, 2.63) | 0.95 (0.39, 2.32) | 1.16 (0.41, 3.32) | --- |
| Any placental malaria | | 1.18 (0.62, 2.26) | 1.30 (0.78, 2.17) | *2.75 (1.20, 6.30) | 0.91 (0.49, 1.68) | 1.93 (0.75, 4.92) |
| Past placental malaria | | 1.21 (0.63, 2.32) | 1.32 (0.79, 2.21) | *2.86 (1.25, 6.51) | 0.92 (0.50, 1.71) | 2.02 (0.78, 5.20) |
| Active placental malaria | | 0.49 (0.07, 3.46) | 0.85 (0.33, 2.19) | 2.26 (0.66, 7.75) | 2.59 (0.80, 8.39) | 0.57 (0.04, 9.16) |
| High-grade past placental malaria | | 3.34 (0.81, 13.71) | 1.68 (0.50, 5.63) | 1.46 (0.43, 5.01) | 2.29 (0.44, 11.90) | 1.83 (0.37, 8.98) |
| Any anemia during pregnancy | | 0.99 (0.59, 1.68) | 1.04 (0.67, 1.62) | 1.57 (0.76, 3.25) | 0.99 (0.54, 1.81) | 1.31 (0.56, 3.07) |
| Metronidazole use during pregnancy | | 0.94 (0.56, 1.59) | 0.95 (0.59, 1.54) | 0.51 (0.23, 1.14) | 0.82 (0.44, 1.51) | 0.40 (0.16, 1.05) |
| Below median gestational weight gain per week | | 1.14 (0.65, 1.99) | 0.98 (0.62, 1.54) | 0.90 (0.44, 1.81) | 0.86 (0.46, 1.63) | *0.36 (0.16, 0.80) |
| Preterm birth | | *3.45 (1.12, 10.60) | *2.83 (1.64, 4.89) | 1.56 (0.39, 6.26) | --- | 2.58 (0.57, 11.74) |
| Low birth weight | | *5.50 (2.79, 10.85) | 2.03 (0.91, 4.51) | 1.77 (0.56, 5.55) | 2.07 (0.68, 6.29) | 0.71 (0.07, 7.30) |
| Small for gestational age | | *2.77 (1.64, 4.68) | 1.56 (0.96, 2.52) | 1.60 (0.83, 3.09) | 1.67 (0.86, 3.23) | *2.27 (1.09, 4.77) |
| Child uncomplicated malaria | 1-3 months | --- | --- | 1.51 (0.64, 3.58) | 1.91 (0.91, 4.03) | 0.84 (0.20, 3.48) |
|  | 4-6 months | --- | --- | --- | 0.83 (0.37, 1.87) | 1.08 (0.41, 2.83) |
|  | 7-9 months | --- | --- | --- | --- | 1.16 (0.37, 3.68) |
| Child any parasitemia | 1-3 months | --- | --- | 1.06 (0.53, 2.14) | 1.83 (0.96, 3.49) | 0.53 (0.16, 1.73) |
|  | 4-6 months | --- | --- | --- | 1.70 (0.88, 3.26) | 0.78 (0.34, 1.78) |
|  | 7-9 months | --- | --- | --- | --- | 0.71 (0.26, 1.90) |
| Child anemia | 2 months | --- | --- | 0.64 (0.29, 1.38) | 1.00 (0.47, 2.11) | 0.97 (0.30, 3.13) |
|  | 6 months | --- | --- | --- | 1.29 (0.57, 2.94) | 0.58 (0.20, 1.69) |
| Child antimalarial use | 1-3 months | --- | --- | 1.74 (0.76, 3.96) | 2.00 (0.96, 4.15) | 0.98 (0.23, 4.16) |
|  | 4-6 months | --- | --- | --- | 1.16 (0.54, 2.51) | 1.14 (0.45, 2.90) |
|  | 7-9 months | --- | --- | --- | --- | 1.20 (0.35, 4.09) |
| Child antibacterial use | 1-3 months | --- | --- | 0.70 (0.36, 1.36) | 0.56 (0.31, 1.03) | 1.29 (0.52, 3.19) |
|  | 4-6 months | --- | --- | --- | 1.05 (0.47, 2.34) | 1.06 (0.37, 3.04) |
|  | 7-9 months | --- | --- | --- | --- | 0.91 (0.36, 2.34) |
| Child beta-lactam use | 1-3 months | --- | --- | 0.63 (0.32, 1.23) | 0.58 (0.32, 1.05) | 1.41 (0.57, 3.46) |
|  | 4-6 months | --- | --- | --- | 1.09 (0.52, 2.31) | 1.04 (0.40, 2.71) |
|  | 7-9 months | --- | --- | --- | --- | 0.90 (0.36, 2.23) |

Asterisks indicate effect estimates in which the confidence interval does not contain the null. Effects were not estimated if there were less than 20 observed cases, or if the mediator preceded the outcome, as indicated with “---”

### Table S9: Effects of Mediators on LAZs

|  | **Mediator** | **Mediator Age** | **Mean difference in LAZ (95% CI)** | | | |
| --- | --- | --- | --- | --- | --- | --- |
|  |  |  | 1-3 months | 4-6 months | 7-9 months | 10-12 months |
| Overall | Any uncomplicated malaria during pregnancy | | -0.04 (-0.32, 0.25) | 0.04 (-0.25, 0.33) | 0.01 (-0.28, 0.30) | 0.03 (-0.26, 0.32) |
|  | Any parasitemia during pregnancy | | -0.12 (-0.35, 0.11) | 0.03 (-0.20, 0.26) | 0.05 (-0.19, 0.28) | -0.00 (-0.24, 0.23) |
|  | Any placental malaria | | -0.06 (-0.24, 0.12) | 0.03 (-0.15, 0.21) | -0.06 (-0.24, 0.12) | -0.14 (-0.32, 0.05) |
|  | Past placental malaria | | -0.07 (-0.25, 0.11) | 0.01 (-0.18, 0.19) | -0.07 (-0.25, 0.12) | -0.15 (-0.33, 0.03) |
|  | Active placental malaria | | 0.06 (-0.35, 0.46) | 0.02 (-0.40, 0.43) | 0.08 (-0.33, 0.50) | 0.06 (-0.36, 0.48) |
|  | High-grade past placental malaria | | *-0.52 (-0.99, -0.05) | -0.37 (-0.84, 0.10) | -0.33 (-0.83, 0.18) | -0.49 (-1.00, 0.01) |
|  | Any anemia during pregnancy | | 0.09 (-0.07, 0.25) | 0.12 (-0.04, 0.28) | -0.10 (-0.26, 0.07) | -0.08 (-0.24, 0.08) |
|  | Metronidazole use during pregnancy | | *0.23 (0.07, 0.39) | 0.16 (-0.01, 0.32) | *0.19 (0.02, 0.36) | *0.17 (0.01, 0.34) |
|  | Below median gestational weight gain per week | | *-0.22 (-0.38, -0.06) | *-0.21 (-0.37, -0.05) | -0.13 (-0.29, 0.04) | -0.12 (-0.28, 0.05) |
|  | Preterm birth | | *-1.69 (-2.08, -1.29) | *-1.38 (-1.79, -0.97) | *-1.05 (-1.48, -0.63) | *-0.64 (-1.06, -0.22) |
|  | Low birth weight | | *-1.54 (-1.87, -1.22) | *-1.22 (-1.56, -0.89) | *-0.90 (-1.25, -0.55) | *-0.57 (-0.92, -0.22) |
|  | Small for gestational age | | *-0.49 (-0.68, -0.31) | *-0.42 (-0.61, -0.22) | *-0.31 (-0.51, -0.12) | *-0.35 (-0.55, -0.16) |
|  | Child uncomplicated malaria | 1-3 months | --- | -0.11 (-0.39, 0.16) | -0.15 (-0.43, 0.13) | -0.19 (-0.46, 0.09) |
|  |  | 4-6 months | --- | --- | 0.03 (-0.19, 0.25) | 0.04 (-0.18, 0.26) |
|  |  | 7-9 months | --- | --- | --- | 0.11 (-0.10, 0.31) |
|  | Child any parasitemia | 1-3 months | --- | -0.04 (-0.23, 0.16) | 0.02 (-0.18, 0.22) | 0.05 (-0.15, 0.25) |
|  |  | 4-6 months | --- | --- | -0.02 (-0.20, 0.15) | 0.04 (-0.13, 0.21) |
|  |  | 7-9 months | --- | --- | --- | -0.05 (-0.23, 0.13) |
|  | Child anemia | 2 months | --- | -0.13 (-0.34, 0.07) | -0.11 (-0.32, 0.10) | -0.11 (-0.32, 0.09) |
|  |  | 6 months | --- | --- | -0.11 (-0.33, 0.11) | -0.06 (-0.28, 0.16) |
|  | Child antimalarial use | 1-3 months | --- | -0.14 (-0.42, 0.13) | -0.19 (-0.47, 0.10) | -0.21 (-0.49, 0.07) |
|  |  | 4-6 months | --- | --- | 0.09 (-0.12, 0.31) | 0.08 (-0.14, 0.29) |
|  |  | 7-9 months | --- | --- | --- | 0.11 (-0.09, 0.32) |
|  | Child antibacterial use | 1-3 months | --- | 0.06 (-0.10, 0.22) | -0.08 (-0.25, 0.08) | -0.12 (-0.28, 0.05) |
|  |  | 4-6 months | --- | --- | 0.04 (-0.15, 0.22) | 0.04 (-0.15, 0.23) |
|  |  | 7-9 months | --- | --- | --- | 0.15 (-0.02, 0.32) |
|  | Child beta-lactam use | 1-3 months | --- | 0.08 (-0.08, 0.24) | -0.04 (-0.20, 0.12) | -0.09 (-0.25, 0.07) |
|  |  | 4-6 months | --- | --- | -0.00 (-0.18, 0.18) | 0.01 (-0.17, 0.19) |
|  |  | 7-9 months | --- | --- | --- | 0.16 (-0.01, 0.33) |
| *Stratrified by Gravidity* | | | | | | |
| Primigravida | Any uncomplicated malaria during pregnancy | | -0.09 (-0.57, 0.39) | -0.19 (-0.69, 0.31) | 0.01 (-0.51, 0.52) | 0.31 (-0.19, 0.81) |
|  | Any parasitemia during pregnancy | | 0.41 (-0.19, 1.01) | 0.37 (-0.26, 0.99) | 0.50 (-0.14, 1.15) | 0.22 (-0.41, 0.86) |
|  | Any placental malaria | | 0.38 (-0.11, 0.87) | 0.17 (-0.34, 0.69) | -0.00 (-0.54, 0.53) | 0.03 (-0.50, 0.57) |
|  | Past placental malaria | | 0.41 (-0.08, 0.89) | 0.19 (-0.32, 0.70) | 0.04 (-0.49, 0.57) | 0.05 (-0.48, 0.58) |
|  | Any anemia during pregnancy | | -0.09 (-0.48, 0.30) | 0.01 (-0.39, 0.42) | -0.15 (-0.57, 0.27) | -0.06 (-0.47, 0.35) |
|  | Metronidazole use during pregnancy | | 0.30 (-0.04, 0.64) | 0.28 (-0.07, 0.64) | *0.44 (0.08, 0.80) | *0.39 (0.03, 0.74) |
|  | Below median gestational weight gain per week | | -0.15 (-0.52, 0.22) | *-0.44 (-0.82, -0.05) | -0.22 (-0.63, 0.18) | -0.31 (-0.70, 0.09) |
|  | Small for gestational age | | *-0.52 (-0.87, -0.16) | *-0.46 (-0.84, -0.09) | -0.37 (-0.76, 0.01) | *-0.54 (-0.91, -0.17) |
|  | Child uncomplicated malaria | 4-6 months | --- | --- | 0.06 (-0.40, 0.52) | -0.21 (-0.66, 0.24) |
|  |  | 7-9 months | --- | --- | --- | -0.02 (-0.48, 0.44) |
|  | Child any parasitemia | 1-3 months | --- | -0.14 (-0.60, 0.33) | -0.14 (-0.62, 0.34) | 0.04 (-0.43, 0.51) |
|  |  | 4-6 months | --- | --- | 0.17 (-0.22, 0.55) | 0.17 (-0.20, 0.54) |
|  |  | 7-9 months | --- | --- | --- | -0.04 (-0.45, 0.38) |
|  | Child anemia | 2 months | --- | 0.11 (-0.38, 0.61) | 0.04 (-0.50, 0.57) | 0.11 (-0.40, 0.63) |
|  |  | 6 months | --- | --- | 0.05 (-0.51, 0.61) | 0.27 (-0.26, 0.81) |
|  | Child antimalarial use | 4-6 months | --- | --- | 0.13 (-0.33, 0.59) | -0.24 (-0.68, 0.20) |
|  |  | 7-9 months | --- | --- | --- | -0.01 (-0.45, 0.43) |
|  | Child antibacterial use | 1-3 months | --- | 0.28 (-0.08, 0.65) | -0.15 (-0.53, 0.22) | -0.23 (-0.59, 0.13) |
|  |  | 4-6 months | --- | --- | *0.44 (0.03, 0.86) | 0.23 (-0.17, 0.64) |
|  |  | 7-9 months | --- | --- | --- | 0.34 (-0.08, 0.75) |
|  | Child beta-lactam use | 1-3 months | --- | *0.41 (0.05, 0.76) | -0.04 (-0.42, 0.33) | -0.16 (-0.52, 0.20) |
|  |  | 4-6 months | --- | --- | 0.36 (-0.06, 0.77) | 0.13 (-0.27, 0.54) |
|  |  | 7-9 months | --- | --- | --- | 0.25 (-0.16, 0.66) |
| Multigravida | Any uncomplicated malaria during pregnancy | | 0.12 (-0.26, 0.50) | 0.23 (-0.15, 0.61) | 0.09 (-0.30, 0.47) | -0.11 (-0.51, 0.28) |
|  | Any parasitemia during pregnancy | | -0.18 (-0.43, 0.07) | 0.01 (-0.24, 0.26) | -0.01 (-0.27, 0.25) | -0.03 (-0.29, 0.23) |
|  | Any placental malaria | | -0.15 (-0.34, 0.05) | -0.02 (-0.22, 0.18) | -0.09 (-0.29, 0.10) | -0.18 (-0.38, 0.02) |
|  | Past placental malaria | | -0.16 (-0.35, 0.04) | -0.04 (-0.24, 0.15) | -0.11 (-0.30, 0.09) | -0.20 (-0.40, 0.00) |
|  | Any anemia during pregnancy | | 0.16 (-0.01, 0.34) | 0.15 (-0.03, 0.32) | -0.07 (-0.26, 0.11) | -0.09 (-0.27, 0.09) |
|  | Metronidazole use during pregnancy | | *0.21 (0.03, 0.40) | 0.13 (-0.06, 0.32) | 0.13 (-0.06, 0.32) | 0.10 (-0.09, 0.29) |
|  | Below median gestational weight gain per week | | *-0.24 (-0.42, -0.06) | -0.17 (-0.36, 0.01) | -0.09 (-0.27, 0.10) | -0.07 (-0.26, 0.12) |
|  | Low birth weight | | *-1.65 (-2.09, -1.22) | *-1.12 (-1.57, -0.67) | *-0.93 (-1.39, -0.46) | *-0.68 (-1.15, -0.21) |
|  | Small for gestational age | | *-0.47 (-0.70, -0.25) | *-0.38 (-0.60, -0.15) | *-0.26 (-0.50, -0.03) | *-0.27 (-0.50, -0.03) |
|  | Child uncomplicated malaria | 1-3 months | --- | -0.14 (-0.44, 0.16) | -0.10 (-0.41, 0.20) | -0.21 (-0.51, 0.10) |
|  |  | 4-6 months | --- | --- | 0.03 (-0.23, 0.29) | 0.12 (-0.13, 0.38) |
|  |  | 7-9 months | --- | --- | --- | 0.13 (-0.11, 0.37) |
|  | Child any parasitemia | 1-3 months | --- | -0.00 (-0.22, 0.22) | 0.06 (-0.17, 0.28) | 0.06 (-0.16, 0.29) |
|  |  | 4-6 months | --- | --- | -0.05 (-0.25, 0.15) | 0.00 (-0.20, 0.20) |
|  |  | 7-9 months | --- | --- | --- | -0.08 (-0.28, 0.13) |
|  | Child anemia | 2 months | --- | -0.15 (-0.38, 0.09) | -0.13 (-0.36, 0.11) | -0.13 (-0.37, 0.10) |
|  |  | 6 months | --- | --- | -0.14 (-0.39, 0.11) | -0.12 (-0.37, 0.12) |
|  | Child antimalarial use | 1-3 months | --- | -0.17 (-0.47, 0.14) | -0.15 (-0.46, 0.16) | -0.23 (-0.54, 0.08) |
|  |  | 4-6 months | --- | --- | 0.08 (-0.17, 0.34) | 0.17 (-0.09, 0.42) |
|  |  | 7-9 months | --- | --- | --- | 0.14 (-0.10, 0.38) |
|  | Child antibacterial use | 1-3 months | --- | -0.02 (-0.20, 0.16) | -0.08 (-0.27, 0.11) | -0.12 (-0.31, 0.07) |
|  |  | 4-6 months | --- | --- | -0.12 (-0.33, 0.10) | -0.03 (-0.24, 0.19) |
|  |  | 7-9 months | --- | --- | --- | 0.12 (-0.08, 0.31) |
|  | Child beta-lactam use | 1-3 months | --- | -0.03 (-0.21, 0.15) | -0.06 (-0.24, 0.12) | -0.10 (-0.29, 0.08) |
|  |  | 4-6 months | --- | --- | -0.13 (-0.34, 0.07) | -0.03 (-0.24, 0.17) |
|  |  | 7-9 months | --- | --- | --- | 0.16 (-0.03, 0.35) |
| *Stratified by Gender* | | | | | | |
| Female | Any uncomplicated malaria during pregnancy | | -0.23 (-0.68, 0.22) | -0.30 (-0.74, 0.14) | -0.29 (-0.71, 0.13) | -0.29 (-0.72, 0.14) |
|  | Any parasitemia during pregnancy | | -0.21 (-0.55, 0.12) | 0.01 (-0.31, 0.34) | -0.09 (-0.40, 0.22) | -0.05 (-0.37, 0.26) |
|  | Any placental malaria | | 0.02 (-0.23, 0.28) | 0.10 (-0.16, 0.35) | 0.02 (-0.22, 0.25) | -0.05 (-0.30, 0.19) |
|  | Past placental malaria | | 0.03 (-0.23, 0.28) | 0.09 (-0.16, 0.35) | 0.02 (-0.21, 0.26) | -0.05 (-0.30, 0.19) |
|  | Any anemia during pregnancy | | 0.03 (-0.19, 0.25) | 0.10 (-0.12, 0.31) | -0.09 (-0.29, 0.12) | -0.03 (-0.24, 0.18) |
|  | Metronidazole use during pregnancy | | 0.09 (-0.14, 0.32) | 0.13 (-0.09, 0.36) | 0.16 (-0.05, 0.37) | 0.16 (-0.06, 0.38) |
|  | Below median gestational weight gain per week | | -0.20 (-0.44, 0.03) | *-0.28 (-0.50, -0.05) | -0.11 (-0.33, 0.10) | -0.11 (-0.33, 0.11) |
|  | Small for gestational age | | *-0.56 (-0.84, -0.28) | *-0.48 (-0.75, -0.20) | *-0.41 (-0.67, -0.15) | *-0.38 (-0.64, -0.11) |
|  | Child uncomplicated malaria | 1-3 months | --- | -0.13 (-0.54, 0.28) | 0.01 (-0.38, 0.40) | -0.07 (-0.47, 0.33) |
|  |  | 4-6 months | --- | --- | *0.30 (0.02, 0.59) | 0.24 (-0.06, 0.53) |
|  |  | 7-9 months | --- | --- | --- | 0.12 (-0.15, 0.40) |
|  | Child any parasitemia | 1-3 months | --- | 0.02 (-0.27, 0.30) | 0.09 (-0.18, 0.37) | 0.14 (-0.13, 0.42) |
|  |  | 4-6 months | --- | --- | 0.21 (-0.01, 0.43) | *0.23 (0.00, 0.46) |
|  |  | 7-9 months | --- | --- | --- | 0.02 (-0.21, 0.25) |
|  | Child anemia | 2 months | --- | -0.16 (-0.45, 0.14) | -0.18 (-0.45, 0.09) | *-0.31 (-0.59, -0.04) |
|  |  | 6 months | --- | --- | -0.21 (-0.49, 0.07) | -0.18 (-0.47, 0.11) |
|  | Child antimalarial use | 4-6 months | --- | --- | *0.30 (0.02, 0.57) | 0.24 (-0.05, 0.52) |
|  |  | 7-9 months | --- | --- | --- | 0.21 (-0.06, 0.47) |
|  | Child antibacterial use | 1-3 months | --- | 0.01 (-0.21, 0.23) | -0.05 (-0.27, 0.16) | -0.13 (-0.34, 0.09) |
|  |  | 4-6 months | --- | --- | 0.03 (-0.21, 0.27) | -0.00 (-0.25, 0.24) |
|  |  | 7-9 months | --- | --- | --- | 0.05 (-0.18, 0.28) |
|  | Child beta-lactam use | 1-3 months | --- | -0.00 (-0.22, 0.21) | -0.05 (-0.25, 0.16) | -0.12 (-0.33, 0.10) |
|  |  | 4-6 months | --- | --- | -0.03 (-0.26, 0.20) | -0.05 (-0.28, 0.19) |
|  |  | 7-9 months | --- | --- | --- | 0.07 (-0.15, 0.30) |
| Male | Any uncomplicated malaria during pregnancy | | 0.01 (-0.36, 0.38) | 0.18 (-0.21, 0.57) | 0.13 (-0.28, 0.55) | 0.18 (-0.23, 0.59) |
|  | Any parasitemia during pregnancy | | -0.07 (-0.40, 0.26) | 0.04 (-0.30, 0.39) | 0.15 (-0.22, 0.52) | 0.02 (-0.34, 0.39) |
|  | Any placental malaria | | -0.18 (-0.44, 0.07) | -0.09 (-0.36, 0.18) | -0.20 (-0.48, 0.08) | -0.25 (-0.53, 0.03) |
|  | Past placental malaria | | -0.21 (-0.47, 0.04) | -0.13 (-0.40, 0.13) | -0.22 (-0.50, 0.06) | -0.28 (-0.56, 0.00) |
|  | Any anemia during pregnancy | | 0.14 (-0.09, 0.37) | 0.15 (-0.09, 0.38) | -0.10 (-0.35, 0.16) | -0.11 (-0.36, 0.14) |
|  | Metronidazole use during pregnancy | | *0.42 (0.19, 0.65) | 0.23 (-0.01, 0.47) | 0.25 (-0.01, 0.51) | 0.21 (-0.05, 0.47) |
|  | Below median gestational weight gain per week | | *-0.23 (-0.45, -0.00) | -0.15 (-0.39, 0.08) | -0.14 (-0.39, 0.11) | -0.12 (-0.37, 0.12) |
|  | Small for gestational age | | *-0.43 (-0.69, -0.17) | *-0.37 (-0.64, -0.10) | -0.23 (-0.52, 0.07) | *-0.33 (-0.62, -0.05) |
|  | Child uncomplicated malaria | 1-3 months | --- | -0.11 (-0.50, 0.27) | -0.24 (-0.65, 0.16) | -0.31 (-0.71, 0.09) |
|  |  | 4-6 months | --- | --- | -0.23 (-0.56, 0.11) | -0.17 (-0.51, 0.16) |
|  |  | 7-9 months | --- | --- | --- | 0.12 (-0.21, 0.44) |
|  | Child any parasitemia | 1-3 months | --- | -0.08 (-0.36, 0.20) | -0.05 (-0.35, 0.24) | -0.05 (-0.34, 0.25) |
|  |  | 4-6 months | --- | --- | -0.24 (-0.50, 0.03) | -0.15 (-0.42, 0.11) |
|  |  | 7-9 months | --- | --- | --- | -0.09 (-0.37, 0.19) |
|  | Child anemia | 2 months | --- | -0.10 (-0.41, 0.21) | -0.05 (-0.38, 0.28) | 0.05 (-0.28, 0.37) |
|  |  | 6 months | --- | --- | -0.04 (-0.40, 0.32) | 0.02 (-0.34, 0.37) |
|  | Child antimalarial use | 1-3 months | --- | -0.14 (-0.52, 0.24) | -0.24 (-0.64, 0.16) | -0.32 (-0.72, 0.07) |
|  |  | 4-6 months | --- | --- | -0.12 (-0.46, 0.21) | -0.11 (-0.44, 0.22) |
|  |  | 7-9 months | --- | --- | --- | 0.07 (-0.25, 0.40) |
|  | Child antibacterial use | 1-3 months | --- | 0.12 (-0.12, 0.36) | -0.08 (-0.33, 0.18) | -0.10 (-0.35, 0.15) |
|  |  | 4-6 months | --- | --- | 0.02 (-0.27, 0.31) | 0.08 (-0.21, 0.36) |
|  |  | 7-9 months | --- | --- | --- | 0.23 (-0.03, 0.50) |
|  | Child beta-lactam use | 1-3 months | --- | 0.19 (-0.05, 0.43) | -0.00 (-0.25, 0.25) | -0.05 (-0.30, 0.20) |
|  |  | 4-6 months | --- | --- | 0.01 (-0.27, 0.29) | 0.06 (-0.22, 0.34) |
|  |  | 7-9 months | --- | --- | --- | 0.23 (-0.03, 0.48) |

Asterisks indicate effect estimates in which the confidence interval does not contain the null. Effects were not estimated if there were less than 20 observed cases, or if the mediator preceded the outcome, as indicated with “---”

### Table S10: Effects of Mediators on Incidence of First-Time Stunting

| **Mediator** | **Mediator Age** | **Risk Ratio for First-Time Stunting (95% CI)** | | |
| --- | --- | --- | --- | --- |
|  |  | Birth | 1-3 months | 4-6 months |
| Any uncomplicated malaria during pregnancy | | 0.84 (0.59, 1.21) | 0.81 (0.50, 1.33) | 0.61 (0.19, 1.94) |
| Any parasitemia during pregnancy | | 1.03 (0.76, 1.41) | 1.21 (0.76, 1.93) | 0.79 (0.39, 1.61) |
| Any placental malaria | | 0.98 (0.77, 1.24) | 0.94 (0.67, 1.34) | 1.69 (0.94, 3.02) |
| Past placental malaria | | 0.98 (0.77, 1.24) | 0.96 (0.67, 1.36) | 1.74 (0.97, 3.13) |
| Active placental malaria | | 1.06 (0.66, 1.71) | 0.64 (0.26, 1.57) | 0.83 (0.20, 3.43) |
| High-grade past placental malaria | | 1.02 (0.60, 1.76) | 1.33 (0.70, 2.50) | --- |
| Any anemia during pregnancy | | 0.84 (0.68, 1.03) | 0.91 (0.67, 1.25) | 0.81 (0.44, 1.49) |
| Metronidazole use during pregnancy | | 0.83 (0.67, 1.03) | 0.78 (0.55, 1.09) | 0.69 (0.37, 1.30) |
| Below median gestational weight gain per week | | 1.16 (0.94, 1.42) | 1.05 (0.75, 1.46) | 1.62 (0.86, 3.04) |
| Preterm birth | | *1.83 (1.31, 2.56) | *2.69 (1.82, 3.99) | --- |
| Low birth weight | | *2.05 (1.58, 2.68) | *2.56 (1.72, 3.81) | --- |
| Small for gestational age | | *1.59 (1.29, 1.95) | *1.78 (1.27, 2.51) | 1.54 (0.74, 3.22) |
| Child uncomplicated malaria | 1-3 months | --- | --- | 1.08 (0.38, 3.07) |
| Child any parasitemia | 1-3 months | --- | --- | 1.23 (0.61, 2.46) |
| Child anemia | 2 months | --- | --- | 1.33 (0.65, 2.73) |
| Child antimalarial use | 1-3 months | --- | --- | 1.33 (0.52, 3.41) |
| Child antibacterial use | 1-3 months | --- | --- | 0.92 (0.51, 1.65) |
| Child beta-lactam use | 1-3 months | --- | --- | 0.99 (0.55, 1.79) |

Asterisks indicate effect estimates in which the confidence interval does not contain the null. Effects were not estimated if there were less than 20 observed cases, or if the mediator preceded the outcome, as indicated with “---”

### Figure S10: Mediator-Outcome Effects, Incidence of First-Time Stunting

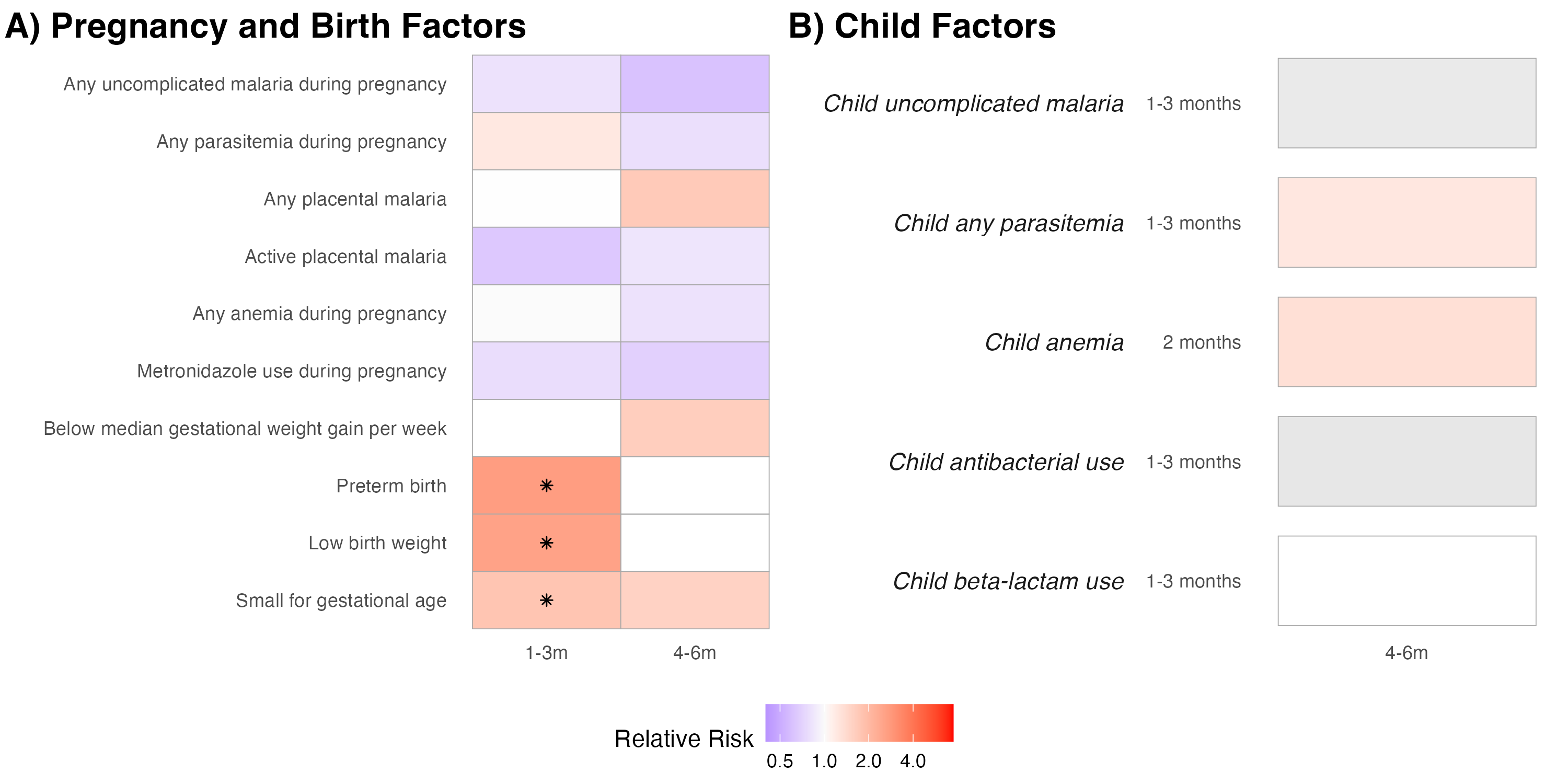

Heatmap of estimated incidence rate ratios for (A) pregnancy and birth factors and (b) infant factors measured at different ages on stunting incidence. Children were considered at risk of incident stunting if they had never been stunted at a prior time point. Incidence rate ratios were estimated with log-binomial models with robust standard errors, adjusting for baseline covariates. If log-binomial models failed to converge, we used modified Poisson regression models. Tiles are shaded in color if the incidence rate ratio is large (IRR > 1.1, IRR < 0.9) or had p-values < 0.05. Estimates where confidence intervals did not cross the null are marked by asterisks. Effects were not estimated if there were less than 20 observed cases, or if the mediator did not precede the outcome – these cases are indicated by the absence of a tile.

### Figure S11: Interventional Indirect Effects for WLZ

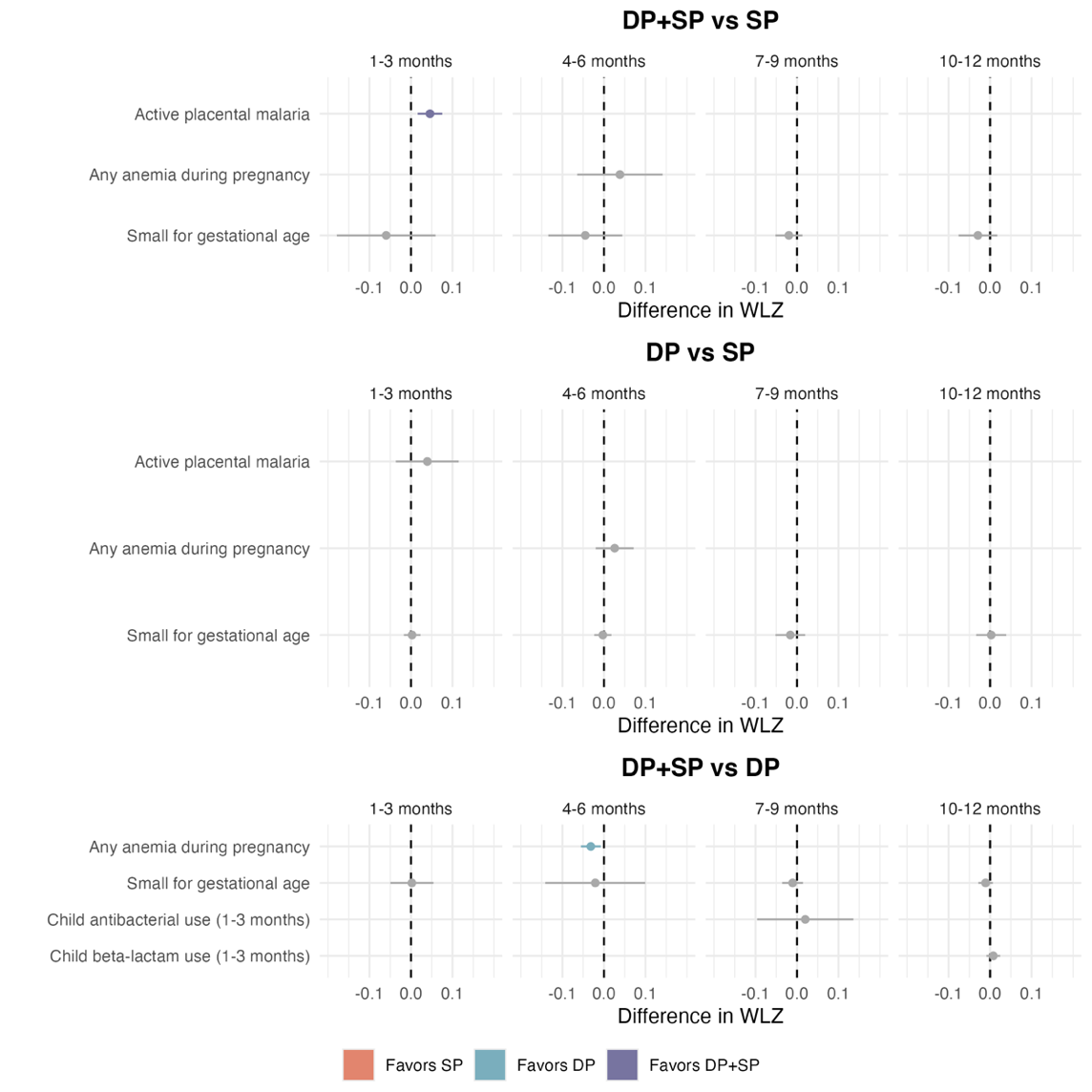

Interventional indirect effects and 95% confidence intervals of IPTp on weight-for-length z-scores (WLZs). Models were adjusted for baseline covariates, as well as intermediate confounders. Statistically significant effect estimates are displayed in color, non-significant estimates are displayed in gray. Color is determined by the comparison arm with higher WLZs (the arm “favored” to provide an increase in WLZ). Interventional indirect effects were only estimated if both the treatment-mediator and mediator-outcome relationships were large (absolute mean difference > 0.5) or had p-values below 0.05.

### Table S11: Interventional Indirect Effects of IPTp on WLZs

|  | **Mediator** | | **Outcome Measurement Age** | **Interventional Indirect Effect (95% CI)** | | |
| --- | --- | --- | --- | --- | --- | --- |
|  |  |  |  | DP+SP vs SP | DP+SP vs DP | DP vs SP |
| Overall | Pooled by Anticipated Negative Indirect Effect | Any anemia during pregnancy, Small for gestational age | 4-6 months | --- | *-0.05 (-0.09, -0.01) | --- |
|  | Active placental malaria | | 1-3 months | *0.05 (0.02, 0.08) | --- | 0.04 (-0.04, 0.12) |
|  | Any anemia during pregnancy | | 4-6 months | 0.04 (-0.06, 0.14) | *-0.03 (-0.06, -0.01) | 0.03 (-0.02, 0.07) |
|  | Small for gestational age | | 1-3 months | -0.06 (-0.18, 0.06) | 0.00 (-0.05, 0.05) | 0.00 (-0.02, 0.02) |
|  |  |  | 4-6 months | -0.05 (-0.13, 0.04) | -0.02 (-0.14, 0.10) | -0.00 (-0.02, 0.02) |
|  |  |  | 7-9 months | -0.02 (-0.05, 0.01) | -0.01 (-0.04, 0.01) | -0.02 (-0.05, 0.02) |
|  |  |  | 10-12 months | -0.03 (-0.08, 0.02) | -0.01 (-0.03, 0.01) | 0.00 (-0.03, 0.04) |
|  | Child antibacterial use (1-3 months) | | 7-9 months | --- | 0.02 (-0.10, 0.14) | --- |
|  | Child beta-lactam use (1-3 months) | | 10-12 months | --- | 0.01 (-0.01, 0.02) | --- |
| *Stratified by Gravidity* | | | | | | |
| Primigravida | Pooled by Anticipated Positive Indirect Effect | Any placental malaria, Past placental malaria | 1-3 months | *0.07 (0.03, 0.12) | --- | *0.06 (0.02, 0.09) |
|  |  | Child beta-lactam use (1-3 months), Child antibacterial use (1-3 months) | 4-6 months | --- | --- | *0.06 (0.02, 0.10) |
|  |  | Child antibacterial use (7-9 months), Any uncomplicated malaria during pregnancy | 10-12 months | *0.34 (0.20, 0.47) | --- | --- |
|  | Pooled by Anticipated Negative Indirect Effect | Child beta-lactam use (1-3 months), Child antibacterial use (1-3 months) | 4-6 months | *-0.06 (-0.12, -0.00) | -0.04 (-0.12, 0.03) | --- |
|  |  | Child beta-lactam use (7-9 months), Below median gestational weight gain per week | 10-12 months | --- | --- | 0.07 (-0.04, 0.19) |
|  | Any placental malaria | | 1-3 months | *0.12 (0.00, 0.23) | --- | 0.02 (-0.02, 0.05) |
|  | Past placental malaria | | 1-3 months | *0.10 (0.05, 0.15) | --- | 0.07 (-0.05, 0.19) |
|  | Below median gestational weight gain per week | | 7-9 months | --- | 0.03 (-0.12, 0.18) | --- |
|  |  |  | 10-12 months | --- | 0.03 (-0.07, 0.13) | 0.04 (-0.09, 0.18) |
|  | Child antibacterial use (1-3 months) | | 4-6 months | -0.06 (-0.15, 0.02) | --- | -0.00 (-0.04, 0.03) |
|  | Child antibacterial use (7-9 months) | | 10-12 months | 0.07 (-0.02, 0.16) | --- | --- |
|  | Child beta-lactam use (1-3 months) | | 4-6 months | -0.04 (-0.12, 0.04) | --- | -0.04 (-0.11, 0.03) |
|  | Child beta-lactam use (7-9 months) | | 10-12 months | --- | --- | 0.05 (-0.03, 0.12) |
| Multigravida | Any anemia during pregnancy | | 4-6 months | --- | -0.02 (-0.07, 0.04) | 0.02 (-0.14, 0.17) |
|  | Small for gestational age | | 7-9 months | -0.03 (-0.06, 0.00) | -0.01 (-0.02, 0.01) | -0.00 (-0.03, 0.02) |
|  |  |  | 10-12 months | -0.04 (-0.15, 0.06) | 0.01 (-0.02, 0.04) | 0.01 (-0.03, 0.04) |
| *Stratified by Child Sex* | | | | | | |
| Female | Pooled by Anticipated Negative Indirect Effect | Child antimalarial use (7-9 months), Small for gestational age | 10-12 months | --- | -0.03 (-0.08, 0.02) | --- |
|  | Small for gestational age | | 1-3 months | --- | -0.01 (-0.03, 0.02) | --- |
|  |  |  | 7-9 months | -0.05 (-0.11, 0.01) | -0.00 (-0.11, 0.11) | --- |
|  |  |  | 10-12 months | 0.01 (-0.02, 0.03) | -0.01 (-0.04, 0.02) | --- |
|  | Child antimalarial use (7-9 months) | | 10-12 months | --- | *-0.06 (-0.10, -0.01) | -0.02 (-0.12, 0.07) |
| Male | Pooled by Anticipated Positive Indirect Effect | Any placental malaria, Past placental malaria | 4-6 months | 0.06 (-0.01, 0.12) | --- | *0.14 (0.09, 0.20) |
|  | Pooled by Anticipated Negative Indirect Effect | Any placental malaria, Past placental malaria | 4-6 months | --- | -0.05 (-0.16, 0.05) | --- |
|  | Any placental malaria | | 4-6 months | *0.06 (0.00, 0.11) | -0.04 (-0.08, 0.01) | *0.08 (0.03, 0.13) |
|  | Past placental malaria | | 4-6 months | *0.04 (0.01, 0.08) | -0.01 (-0.07, 0.05) | 0.06 (-0.04, 0.16) |
|  | Metronidazole use during pregnancy | | 1-3 months | *0.05 (0.01, 0.10) | 0.02 (-0.05, 0.09) | --- |
|  |  |  | 10-12 months | *-0.08 (-0.12, -0.03) | -0.02 (-0.08, 0.03) | --- |
|  | Small for gestational age | | 7-9 months | -0.02 (-0.07, 0.03) | -0.05 (-0.13, 0.02) | -0.02 (-0.16, 0.11) |
|  | Child antimalarial use (1-3 months) | | 4-6 months | -0.01 (-0.05, 0.03) | --- | -0.01 (-0.05, 0.03) |

Asterisks indicate effect estimates in which the confidence interval does not contain the null. Interventional indirect effects were only estimated if both the treatment-mediator and mediator-outcome relationships were large (absolute mean difference > 0.5) or had p-values below 0.05. Mediators are grouped based on the anticipated direction of their indirect effect estimate. We expected a positive indirect effect if the treatment increased a mediator that is positively associated with growth or decreased a mediator that is negatively associated with growth. We expected a negative indirect effect if the treatment decreased a mediator that is positively associated with growth or increased a mediator that is negatively associated with growth.

### Figure S12: Interventional Indirect Effects for WLZ, Stratified by Gravidity

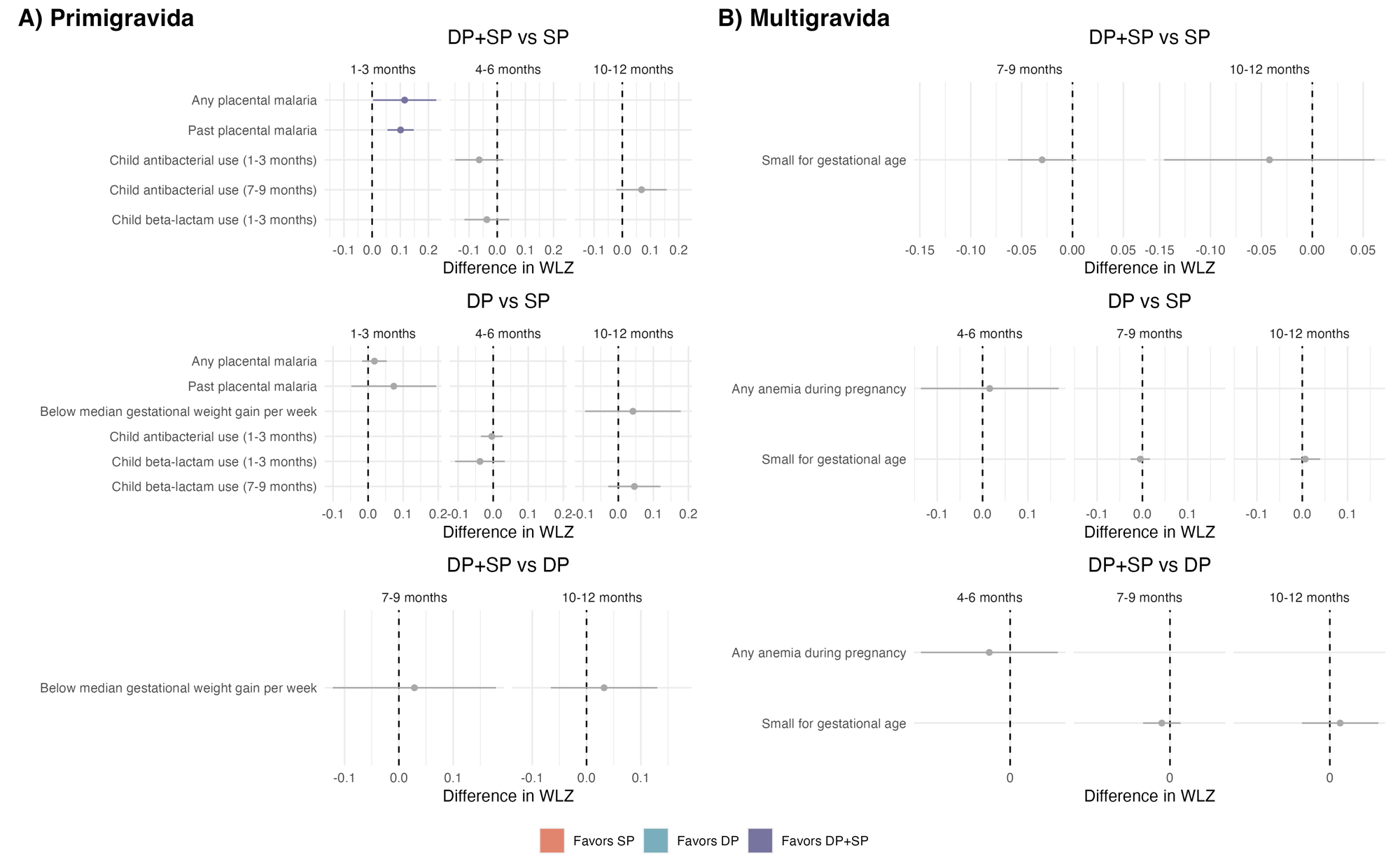

Interventional indirect effects and 95% confidence intervals of IPTp on weight-for-length z-scores (WLZs), by gravidity. Models were adjusted for baseline covariates, as well as intermediate confounders. Statistically significant effect estimates are displayed in color, non-significant estimates are displayed in gray. Color is determined by the comparison arm with higher WLZs (the arm “favored” to provide an increase in WLZ). Interventional indirect effects were only estimated if both the treatment-mediator and mediator-outcome relationships were large (absolute mean difference > 0.5) or had p-values below 0.05.

### Figure S13: Interventional Indirect Effects for WLZ, Stratified by Child Sex

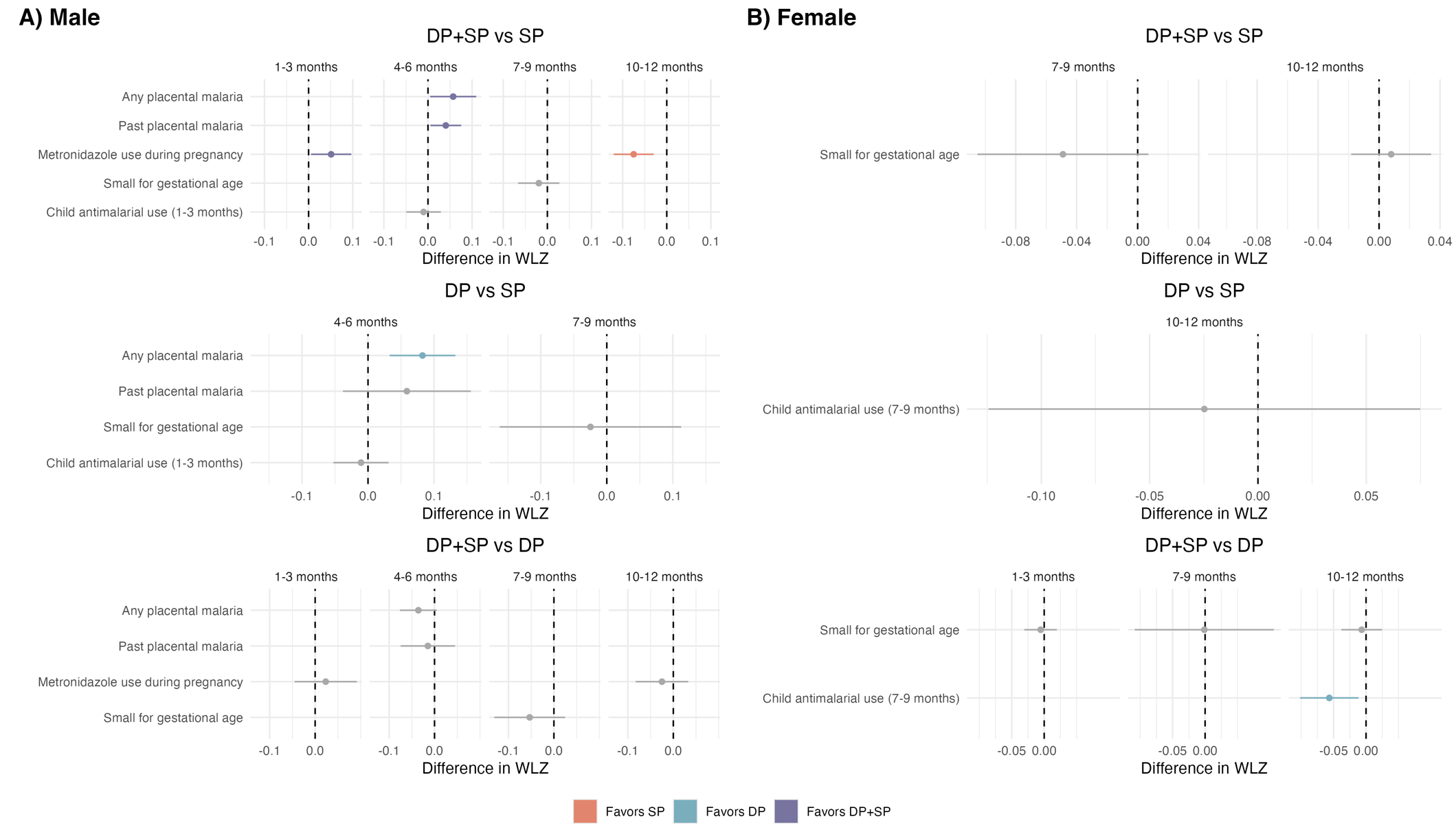

Interventional indirect effects and 95% confidence intervals of IPTp on weight-for-length z-scores (WLZs), by child sex. Models were adjusted for baseline covariates, as well as intermediate confounders. Statistically significant effect estimates are displayed in color, non-significant estimates are displayed in gray. Color is determined by the comparison arm with higher WLZs (the arm “favored” to provide an increase in WLZ). Interventional indirect effects were only estimated if both the treatment-mediator and mediator-outcome relationships were large (absolute mean difference > 0.5) or had p-values below 0.05.

### Table S12: Interventional Indirect Effects of IPTp on Incidence of Wasting

| **Mediator** | **Outcome Measurement Age** | **Intervention Indirect Effect, Difference in Incidence Percentage Points (95% CI)** | | |
| --- | --- | --- | --- | --- |
|  |  | DP+SP vs SP | DP+SP vs DP | DP vs SP |
| Any uncomplicated malaria during pregnancy | 7-9 months | -0.00 (-0.90, 0.90) | --- | --- |
|  | 10-12 months | --- | --- | *0.21 (0.00, 0.42) |
| Any parasitemia during pregnancy | 1-3 months | -0.16 (-2.63, 2.30) | --- | -0.04 (-1.93, 1.84) |
|  | 7-9 months | 0.15 (-1.76, 2.07) | --- | --- |
| Any placental malaria | 1-3 months | --- | *0.58 (0.08, 1.08) | -0.50 (-1.15, 0.16) |
|  | 4-6 months | --- | 0.09 (-0.94, 1.13) | --- |
|  | 10-12 months | --- | --- | 0.01 (-0.79, 0.80) |
| Past placental malaria | 1-3 months | --- | 0.62 (-0.01, 1.24) | *-0.84 (-1.32, -0.36) |
|  | 4-6 months | --- | 0.57 (-0.07, 1.22) | --- |
|  | 10-12 months | --- | --- | 0.11 (-0.11, 0.33) |
| Active placental malaria | 1-3 months | --- | --- | 0.52 (-0.02, 1.05) |
|  | 10-12 months | --- | --- | *-0.46 (-0.87, -0.05) |
| High-grade past placental malaria | 4-6 months | 1.65 (-0.57, 3.88) | --- | --- |
|  | 7-9 months | 1.04 (-2.13, 4.20) | --- | -0.91 (-5.28, 3.45) |
|  | 10-12 months | 1.49 (-1.72, 4.70) | --- | 0.13 (-3.32, 3.57) |
| Any anemia during pregnancy | 4-6 months | --- | 0.20 (-0.17, 0.56) | 0.05 (-0.39, 0.48) |
|  | 10-12 months | --- | -0.01 (-0.30, 0.28) | -0.39 (-1.87, 1.08) |
| Metronidazole use during pregnancy | 4-6 months | --- | --- | 0.19 (-1.19, 1.58) |
|  | 7-9 months | 0.64 (-0.34, 1.61) | --- | --- |
|  | 10-12 months | --- | --- | -0.09 (-1.49, 1.31) |
| Below median gestational weight gain per week | 4-6 months | --- | 0.08 (-0.26, 0.43) | --- |
|  | 7-9 months | *0.13 (0.03, 0.24) | --- | -0.47 (-1.96, 1.02) |
|  | 10-12 months | --- | 0.34 (-0.08, 0.76) | -0.16 (-0.59, 0.27) |
| Preterm birth | 1-3 months | --- | 0.23 (-0.62, 1.08) | --- |
|  | 4-6 months | --- | 0.63 (-0.46, 1.73) | --- |
|  | 10-12 months | --- | -0.07 (-0.51, 0.36) | --- |
| Low birth weight | 1-3 months | --- | 0.48 (-1.76, 2.72) | --- |
|  | 4-6 months | --- | *0.15 (0.04, 0.26) | --- |
|  | 7-9 months | -0.30 (-1.03, 0.44) | --- | -0.04 (-3.13, 3.04) |
|  | 10-12 months | --- | -0.00 (-0.32, 0.31) | --- |
| Small for gestational age | 1-3 months | --- | 0.00 (-2.71, 2.71) | 1.73 (-1.36, 4.82) |
|  | 4-6 months | 0.24 (-0.08, 0.55) | -0.13 (-1.22, 0.96) | --- |
|  | 7-9 months | --- | --- | 0.37 (-1.25, 1.98) |
|  | 10-12 months | --- | 0.03 (-1.21, 1.27) | --- |
| Child uncomplicated malaria (1-3 months) | 4-6 months | --- | 0.16 (-0.31, 0.63) | --- |
|  | 7-9 months | --- | -0.17 (-2.53, 2.19) | *0.31 (0.03, 0.60) |
|  | 10-12 months | --- | 0.15 (-0.38, 0.68) | 0.25 (-0.16, 0.66) |
| Child uncomplicated malaria (7-9 months) | 10-12 months | --- | --- | -0.13 (-0.49, 0.22) |
| Child any parasitemia (1-3 months) | 7-9 months | --- | --- | 0.12 (-0.18, 0.42) |
|  | 10-12 months | --- | --- | 0.10 (-0.26, 0.45) |
| Child any parasitemia (4-6 months) | 7-9 months | --- | 0.20 (-2.24, 2.63) | --- |
|  | 10-12 months | 0.06 (-1.77, 1.90) | 0.10 (-1.37, 1.56) | --- |
| Child any parasitemia (7-9 months) | 10-12 months | --- | --- | -0.14 (-0.45, 0.18) |
| Child anemia (6 months) | 7-9 months | -0.13 (-0.29, 0.04) | --- | --- |
|  | 10-12 months | 0.24 (-2.02, 2.51) | --- | --- |
| Child antimalarial use (4-6 months) | 7-9 months | 0.00 (-2.04, 2.05) | --- | --- |
| Child antimalarial use (7-9 months) | 10-12 months | --- | 0.05 (-1.01, 1.11) | -0.20 (-0.97, 0.57) |

Asterisks indicate effect estimates in which the confidence interval does not contain the null. Interventional indirect effects were only estimated if both the treatment-mediator and mediator-outcome relationships were large (risk ratio > 1.1 or risk ratio < 0.9) or had p-values below 0.05. Effects were not estimated if there were less than 20 observed cases, or if the mediator preceded the outcome. Effects that were not estimated are indicated with “---”.

### Figure S15: Interventional Indirect Effects for LAZ

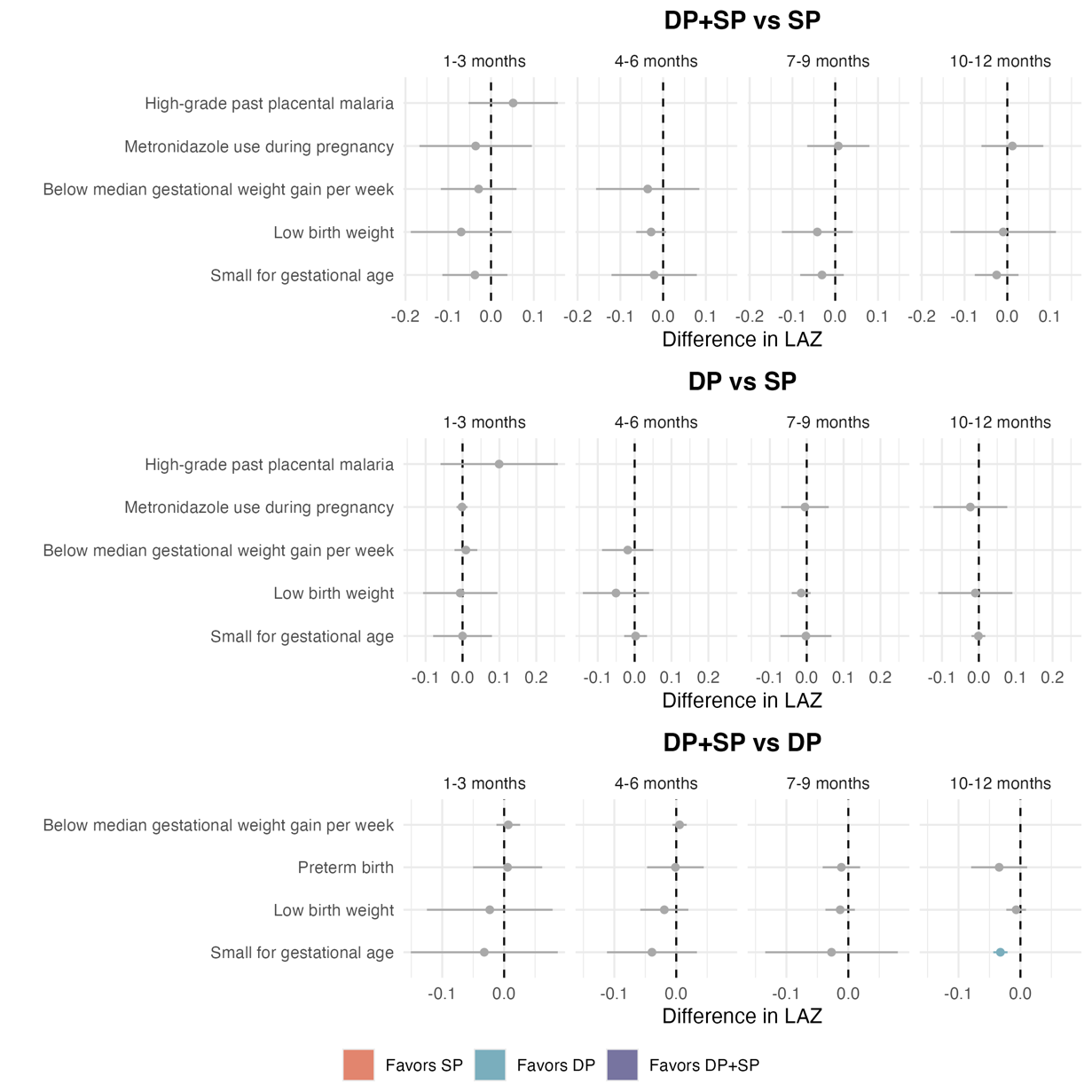

Interventional indirect effects and 95% confidence intervals of IPTp on length-for-age z-scores (LAZs). Models were adjusted for baseline covariates, as well as intermediate confounders. Statistically significant effect estimates are displayed in color, non-significant estimates are displayed in gray. Color is determined by the comparison arm with higher LAZs (the arm “favored” to provide an increase in LAZ). Interventional indirect effects were only estimated if both the treatment-mediator and mediator-outcome relationships were large (absolute mean difference > 0.5) or had p-values below 0.05.

### Table S13: Interventional Indirect Effects of IPTp on LAZs

|  | **Mediator** | | **Outcome Measurement Age** | **Interventional Indirect Effect (95% CI)** | | |
| --- | --- | --- | --- | --- | --- | --- |
|  |  |  |  | DP+SP vs SP | DP+SP vs DP | DP vs SP |
| Overall | Pooled by Anticipated Negative Indirect Effect | Below median gestational weight gain per week, Low birth weight, Small for gestational age, Metronidazole use during pregnancy | 1-3 months | *-0.15 (-0.23, -0.08) | --- | *-0.08 (-0.13, -0.03) |
|  |  | Preterm birth, Low birth weight, Small for gestational age | 1-3 months | --- | -0.04 (-0.08, 0.00) | --- |
|  |  | Below median gestational weight gain per week, Low birth weight, Small for gestational age | 4-6 months | -0.08 (-0.19, 0.03) | --- | -0.04 (-0.11, 0.03) |
|  |  | Preterm birth, Low birth weight, Small for gestational age | 4-6 months | --- | -0.04 (-0.10, 0.03) | --- |
|  |  | Low birth weight, Small for gestational age, Metronidazole use during pregnancy | 7-9 months | -0.04 (-0.13, 0.04) | --- | -0.03 (-0.14, 0.09) |
|  |  | Preterm birth, Low birth weight, Small for gestational age | 7-9 months | --- | *-0.03 (-0.05, -0.00) | --- |
|  |  | Low birth weight, Small for gestational age, Metronidazole use during pregnancy | 10-12 months | -0.06 (-0.17, 0.05) | --- | 0.01 (-0.01, 0.03) |
|  |  | Preterm birth, Low birth weight, Small for gestational age | 10-12 months | --- | *-0.03 (-0.05, -0.01) | --- |
|  | High-grade past placental malaria | | 1-3 months | *0.06 (0.02, 0.10) | --- | *0.08 (0.04, 0.11) |
|  | Metronidazole use during pregnancy | | 1-3 months | -0.04 (-0.17, 0.10) | --- | -0.00 (-0.02, 0.01) |
|  |  |  | 7-9 months | 0.01 (-0.07, 0.08) | --- | -0.00 (-0.07, 0.06) |
|  |  |  | 10-12 months | 0.01 (-0.06, 0.08) | --- | -0.02 (-0.12, 0.08) |
|  | Below median gestational weight gain per week | | 1-3 months | -0.03 (-0.12, 0.06) | 0.01 (-0.01, 0.03) | 0.01 (-0.02, 0.04) |
|  |  |  | 4-6 months | -0.04 (-0.16, 0.08) | 0.01 (-0.01, 0.02) | -0.02 (-0.09, 0.05) |
|  | Preterm birth | | 1-3 months | --- | 0.01 (-0.05, 0.06) | --- |
|  |  |  | 4-6 months | --- | -0.00 (-0.05, 0.04) | --- |
|  |  |  | 7-9 months | --- | -0.01 (-0.04, 0.02) | --- |
|  |  |  | 10-12 months | --- | -0.03 (-0.08, 0.01) | --- |
|  | Low birth weight | | 1-3 months | -0.07 (-0.19, 0.05) | -0.02 (-0.12, 0.08) | -0.01 (-0.11, 0.09) |
|  |  |  | 4-6 months | -0.03 (-0.06, 0.01) | -0.02 (-0.06, 0.02) | -0.05 (-0.14, 0.04) |
|  |  |  | 7-9 months | -0.04 (-0.12, 0.04) | -0.01 (-0.04, 0.01) | -0.01 (-0.04, 0.01) |
|  |  |  | 10-12 months | -0.01 (-0.13, 0.11) | -0.01 (-0.02, 0.01) | -0.01 (-0.11, 0.09) |
|  | Small for gestational age | | 1-3 months | -0.04 (-0.11, 0.04) | -0.03 (-0.15, 0.09) | 0.00 (-0.08, 0.08) |
|  |  |  | 4-6 months | -0.02 (-0.12, 0.08) | -0.04 (-0.11, 0.03) | 0.00 (-0.03, 0.03) |
|  |  |  | 7-9 months | -0.03 (-0.08, 0.02) | -0.03 (-0.13, 0.08) | -0.00 (-0.07, 0.07) |
|  |  |  | 10-12 months | -0.02 (-0.08, 0.03) | *-0.03 (-0.04, -0.02) | -0.00 (-0.02, 0.02) |
| *Stratified by Gravidity* | | | | | | |
| Primigravida | Pooled by Anticipated Positive Indirect Effect | Child beta-lactam use (1-3 months), Below median gestational weight gain per week | 4-6 months | --- | 0.05 (-0.02, 0.12) | --- |
|  | Pooled by Anticipated Negative Indirect Effect | Child beta-lactam use (1-3 months), Below median gestational weight gain per week, Small for gestational age | 4-6 months | --- | --- | *-0.15 (-0.25, -0.05) |
|  |  | Any parasitemia during pregnancy, Metronidazole use during pregnancy | 7-9 months | --- | --- | -0.07 (-0.23, 0.09) |
|  |  | Small for gestational age, Metronidazole use during pregnancy | 10-12 months | --- | --- | -0.00 (-0.07, 0.06) |
|  | Any parasitemia during pregnancy | | 7-9 months | --- | --- | -0.03 (-0.15, 0.10) |
|  | Metronidazole use during pregnancy | | 7-9 months | --- | --- | *-0.02 (-0.05, -0.00) |
|  |  |  | 10-12 months | --- | --- | -0.02 (-0.09, 0.06) |
|  | Below median gestational weight gain per week | | 4-6 months | --- | --- | -0.10 (-0.24, 0.04) |
|  | Small for gestational age | | 1-3 months | -0.01 (-0.11, 0.10) | --- | -0.01 (-0.04, 0.02) |
|  |  |  | 4-6 months | 0.03 (-0.10, 0.16) | --- | -0.01 (-0.12, 0.10) |
|  |  |  | 10-12 months | -0.06 (-0.16, 0.04) | --- | 0.01 (-0.03, 0.06) |
|  | Child antibacterial use (4-6 months) | | 7-9 months | --- | 0.07 (-0.05, 0.20) | --- |
|  | Child beta-lactam use (1-3 months) | | 4-6 months | -0.04 (-0.12, 0.04) | 0.04 (-0.05, 0.14) | -0.04 (-0.13, 0.04) |
| Multigravida | Pooled by Anticipated Negative Indirect Effect | Below median gestational weight gain per week, Small for gestational age, Metronidazole use during pregnancy | 1-3 months | *-0.11 (-0.15, -0.08) | --- | -0.04 (-0.08, 0.00) |
|  |  | Small for gestational age, Metronidazole use during pregnancy | 1-3 months | --- | -0.05 (-0.14, 0.04) | --- |
|  | Metronidazole use during pregnancy | | 1-3 months | -0.02 (-0.06, 0.02) | -0.01 (-0.09, 0.07) | -0.01 (-0.03, 0.01) |
|  | Below median gestational weight gain per week | | 1-3 months | -0.03 (-0.07, 0.01) | -0.00 (-0.09, 0.09) | *-0.03 (-0.05, -0.00) |
|  | Small for gestational age | | 1-3 months | -0.05 (-0.16, 0.06) | -0.05 (-0.10, 0.00) | -0.01 (-0.08, 0.06) |
|  |  |  | 4-6 months | -0.04 (-0.15, 0.07) | -0.04 (-0.09, 0.01) | -0.00 (-0.13, 0.13) |
|  |  |  | 7-9 months | *-0.01 (-0.03, -0.00) | *-0.04 (-0.08, -0.00) | -0.02 (-0.05, 0.00) |
|  |  |  | 10-12 months | -0.05 (-0.13, 0.03) | -0.03 (-0.07, 0.01) | -0.01 (-0.04, 0.02) |
| *Stratified by Child Gender* | | | | | | |
| Female | Pooled by Anticipated Positive Indirect Effect | Child antimalarial use (4-6 months), Child uncomplicated malaria (4-6 months) | 7-9 months | 0.02 (-0.09, 0.12) | 0.02 (-0.05, 0.09) | --- |
|  |  | Child anemia (2 months), Child any parasitemia (4-6 months) | 10-12 months | --- | --- | *0.06 (0.03, 0.10) |
|  | Pooled by Anticipated Negative Indirect Effect | Below median gestational weight gain per week, Small for gestational age | 4-6 months | -0.06 (-0.12, 0.01) | --- | --- |
|  | Below median gestational weight gain per week | | 4-6 months | -0.03 (-0.14, 0.07) | -0.01 (-0.02, 0.01) | -0.00 (-0.02, 0.02) |
|  | Small for gestational age | | 1-3 months | -0.05 (-0.14, 0.03) | *-0.07 (-0.13, -0.00) | --- |
|  |  |  | 4-6 months | -0.02 (-0.13, 0.08) | -0.02 (-0.06, 0.01) | --- |
|  |  |  | 7-9 months | 0.01 (-0.10, 0.11) | 0.02 (-0.02, 0.07) | --- |
|  |  |  | 10-12 months | *-0.02 (-0.04, -0.01) | -0.06 (-0.16, 0.04) | --- |
|  | Child uncomplicated malaria (4-6 months) | | 7-9 months | 0.01 (-0.06, 0.07) | -0.00 (-0.08, 0.08) | --- |
|  | Child any parasitemia (4-6 months) | | 10-12 months | 0.01 (-0.04, 0.05) | --- | 0.02 (-0.05, 0.08) |
|  | Child anemia (2 months) | | 10-12 months | --- | --- | 0.03 (-0.02, 0.09) |
|  | Child antimalarial use (4-6 months) | | 7-9 months | 0.02 (-0.03, 0.06) | *-0.02 (-0.05, -0.00) | -0.02 (-0.09, 0.05) |
| Male | Pooled by Anticipated Negative Indirect Effect | Below median gestational weight gain per week, Small for gestational age | 1-3 months | --- | --- | 0.02 (-0.14, 0.18) |
|  |  | Small for gestational age, Metronidazole use during pregnancy | 1-3 months | --- | -0.05 (-0.09, 0.00) | --- |
|  |  | Below median gestational weight gain per week, Small for gestational age, Metronidazole use during pregnancy | 1-3 months | -0.07 (-0.20, 0.07) | --- | --- |
|  | Metronidazole use during pregnancy | | 1-3 months | -0.07 (-0.15, 0.01) | -0.03 (-0.06, 0.01) | --- |
|  | Below median gestational weight gain per week | | 1-3 months | 0.01 (-0.03, 0.05) | --- | -0.01 (-0.04, 0.03) |
|  | Small for gestational age | | 1-3 months | -0.00 (-0.12, 0.11) | -0.04 (-0.15, 0.06) | -0.00 (-0.09, 0.09) |
|  |  |  | 4-6 months | -0.01 (-0.06, 0.05) | -0.02 (-0.05, 0.01) | 0.00 (-0.05, 0.05) |
|  |  |  | 10-12 months | -0.00 (-0.11, 0.11) | -0.01 (-0.18, 0.15) | -0.01 (-0.05, 0.02) |

Asterisks indicate effect estimates in which the confidence interval does not contain the null. Interventional indirect effects were only estimated if both the treatment-mediator and mediator-outcome relationships were large (absolute mean difference > 0.5) or had p-values below 0.05. Mediators are grouped based on the anticipated direction of their indirect effect estimate. We expected a positive indirect effect if the treatment increased a mediator that is positively associated with growth or decreased a mediator that is negatively associated with growth. We expected a negative indirect effect if the treatment decreased a mediator that is positively associated with growth or increased a mediator that is negatively associated with growth.

### Figure S16: Interventional Indirect Effects for LAZ, Stratified by Gravidity

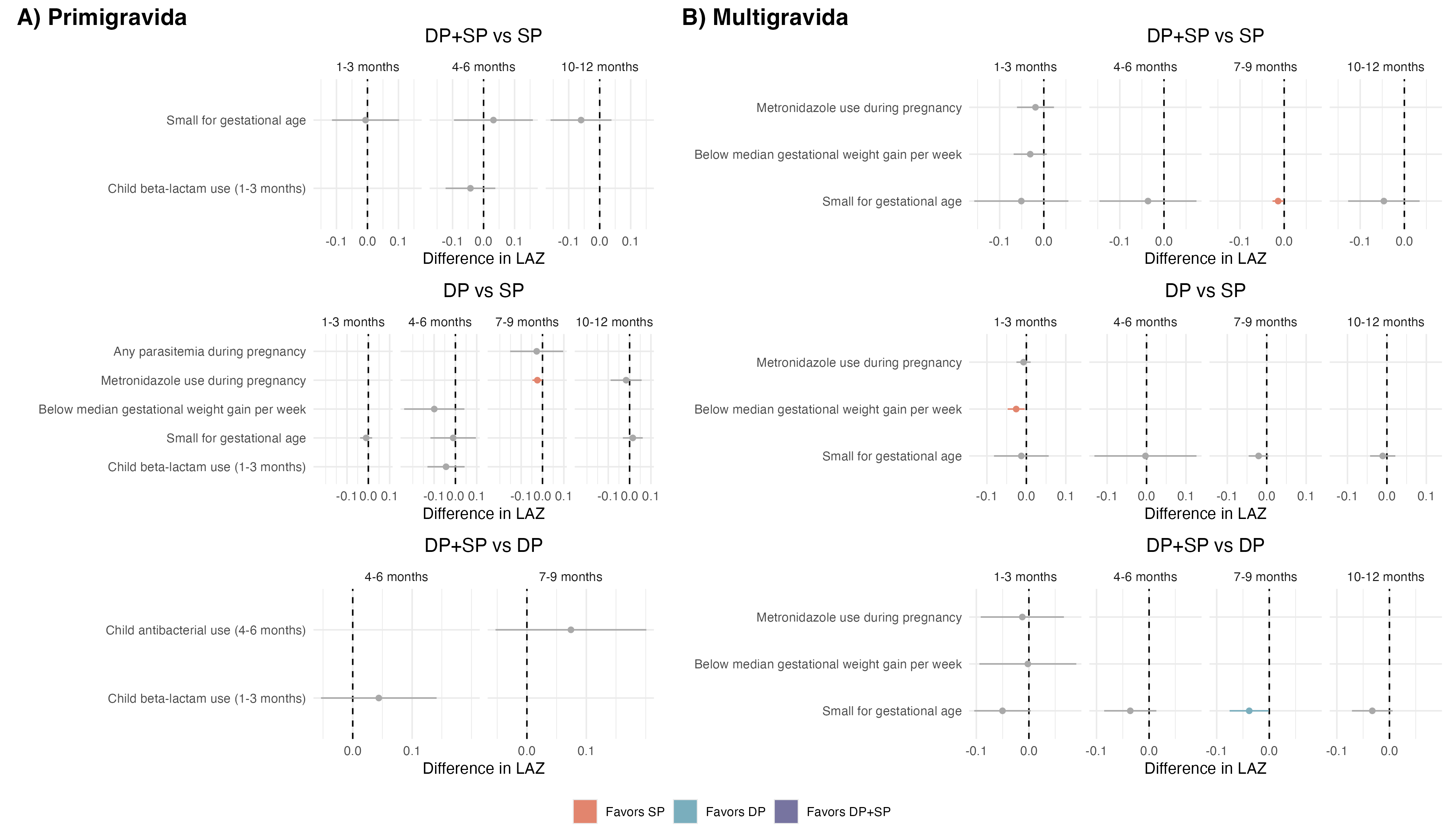

Interventional indirect effects and 95% confidence intervals of IPTp on length-for-age z-scores (LAZs), by gravidity. Models were adjusted for baseline covariates, as well as intermediate confounders. Statistically significant effect estimates are displayed in color, non-significant estimates are displayed in gray. Color is determined by the comparison arm with higher LAZs (the arm “favored” to provide an increase in LAZ). Interventional indirect effects were only estimated if both the treatment-mediator and mediator-outcome relationships were large (absolute mean difference > 0.5) or had p-values below 0.05.

### Figure S17: Interventional Indirect Effects for LAZ, Stratified by Child Sex

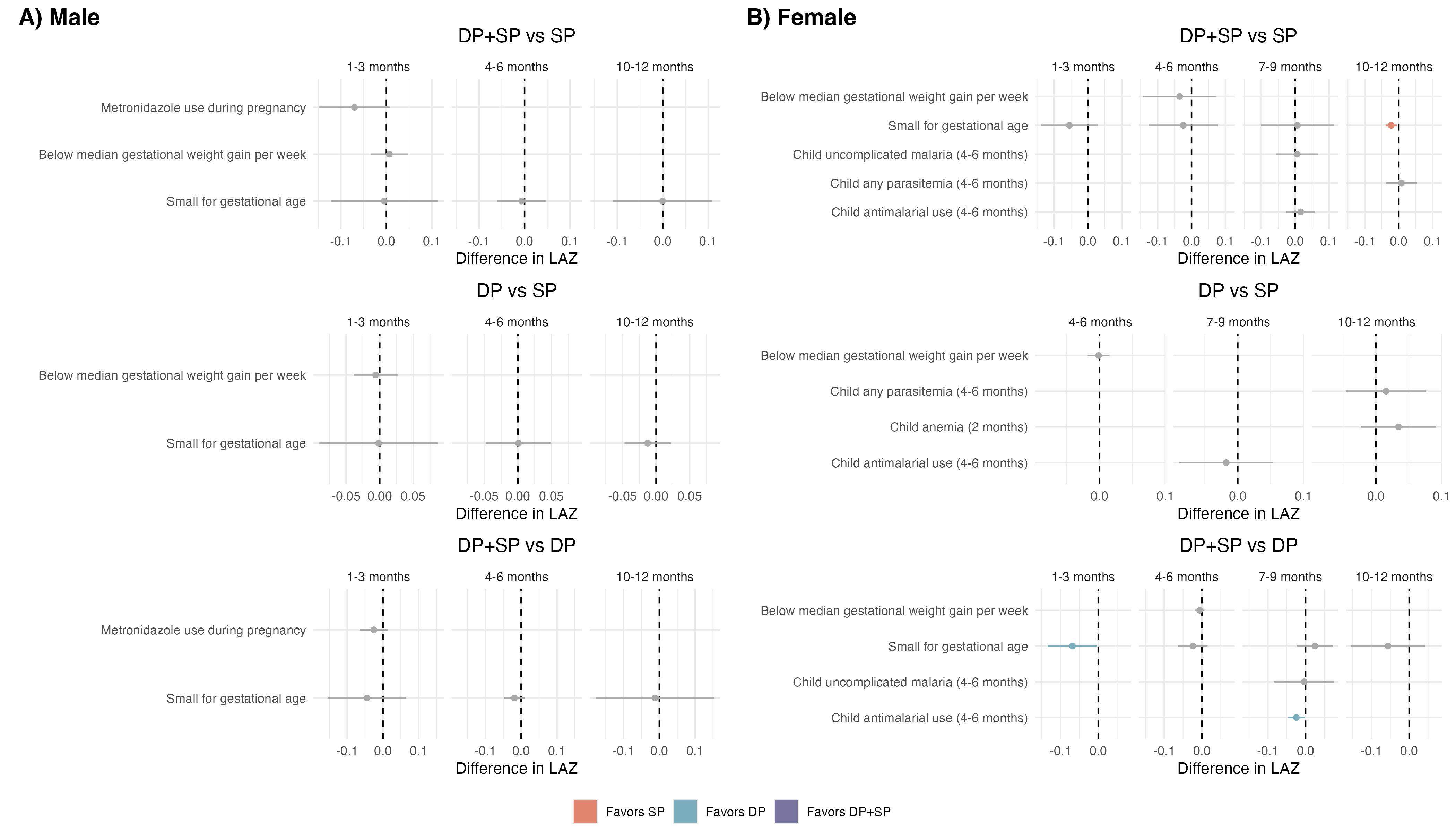

Interventional indirect effects and 95% confidence intervals of IPTp on length-for-age z-scores (LAZs), by child sex. Models were adjusted for baseline covariates, as well as intermediate confounders. Statistically significant effect estimates are displayed in color, non-significant estimates are displayed in gray. Color is determined by the comparison arm with higher LAZs (the arm “favored” to provide an increase in LAZ). Interventional indirect effects were only estimated if both the treatment-mediator and mediator-outcome relationships were large (absolute mean difference > 0.5) or had p-values below 0.05.

### Table S14: Interventional Indirect Effects of IPTp on Incidence of First-Time Stunting

| **Mediator** | **Outcome Measurement Age** | **Intervention Indirect Effect, Difference in Incidence Percentage Points (95% CI)** | | |
| --- | --- | --- | --- | --- |
|  |  | DP+SP vs SP | DP+SP vs DP | DP vs SP |
| Any uncomplicated malaria during pregnancy | 1-3 months | 0.02 (-0.79, 0.82) | --- | 1.35 (-0.31, 3.02) |
|  | 4-6 months | -0.81 (-3.10, 1.48) | --- | 0.39 (-3.14, 3.91) |
| Any parasitemia during pregnancy | 1-3 months | 0.13 (-1.08, 1.34) | --- | -0.19 (-1.11, 0.73) |
|  | 4-6 months | -0.78 (-2.61, 1.05) | --- | -0.12 (-0.55, 0.30) |
| Any placental malaria | 4-6 months | -0.04 (-0.67, 0.58) | --- | --- |
| Past placental malaria | 4-6 months | 0.43 (-0.24, 1.10) | --- | --- |
| Active placental malaria | 1-3 months | 0.07 (-0.57, 0.71) | --- | 0.55 (-0.99, 2.10) |
|  | 4-6 months | -0.10 (-0.56, 0.36) | --- | 0.14 (-0.51, 0.79) |
| High-grade past placental malaria | 1-3 months | -0.58 (-3.58, 2.43) | --- | -1.20 (-3.38, 0.97) |
| Any anemia during pregnancy | 4-6 months | -0.04 (-0.91, 0.82) | --- | -2.00 (-5.60, 1.59) |
| Metronidazole use during pregnancy | 1-3 months | -0.05 (-0.86, 0.76) | --- | --- |
|  | 4-6 months | -0.16 (-1.02, 0.71) | --- | --- |
| Preterm birth | 1-3 months | --- | 0.80 (-0.06, 1.67) | --- |
| Low birth weight | 1-3 months | 0.85 (-3.89, 5.58) | -1.13 (-4.41, 2.16) | 0.06 (-1.15, 1.27) |
| Small for gestational age | 1-3 months | 0.55 (-3.43, 4.54) | -0.02 (-1.24, 1.20) | 0.24 (-3.76, 4.25) |
|  | 4-6 months | *1.44 (0.28, 2.60) | --- | --- |
| Child antimalarial use (1-3 months) | 4-6 months | --- | --- | 0.12 (-0.70, 0.94) |

Asterisks indicate effect estimates in which the confidence interval does not contain the null. Interventional indirect effects were only estimated if both the treatment-mediator and mediator-outcome relationships were large (risk ratio > 1.1 or risk ratio < 0.9) or had p-values below 0.05. Effects were not estimated if there were less than 20 observed cases, or if the mediator preceded the outcome. Effects that were not estimated are indicated with “---”.
